# Insights on Telemedicine Use by Physiatrists Before, During, and Beyond the COVID-19 Pandemic

**DOI:** 10.1101/2022.02.19.22270722

**Authors:** M. J. Sobrepera, A. Weseloh, D. Chaudhri, M. J. Johnson

## Abstract

**Purpose:** To determine if physiatrists increased their use of telemedicine during the first 11 months of the COVID-19 pandemic and if changes are expected to persist post-pandemic. And to identify key tasks which require video for telemedicine.

**Materials and Methods:** A survey containing questions about the telemedicine tools used before, during, and after (planned) the COVID-19 pandemic was distributed to physiatrists. Analysis was conducted to evaluate the change in usage of telemedicine due to the pandemic and predict whether the pandemic will lead to a durable change in usage. Tasks which physiatrists have/believe they can complete with different modes of interaction were explored.

**Results:** Responses from 56 physiatrists showed a 105% increase in video-based telemedicine use during the pandemic. The use of phone and video communications for care delivery significantly increased. 79% of respondents planned to use video-based telemedicine post-pandemic, a significant increase from pre-pandemic use. Motor assessments, cognitive assessments, stretching, strength building, and orthotics assessment and prescription were identified as key tasks that require video for telemedicine.

**Conclusion:** This study confirms increased use of telemedicine by physiatrists during the pandemic and suggests this shift will be durable. Key tasks where video is necessary for telemedicine were identified.

## 1. Introduction

Physiatry is a specialty that leverages a unique interdisciplinary approach to provide coordinated care to patients with congenital or acquired disabilities involving, but not limited to, pediatric limb deficiency, prosthetics and orthotics, spinal cord injury, brain injury, stroke, musculoskeletal injury, burn injury, as well as cardiopulmonary and transplant rehabilitation. Physiatrists often lead teams of allied medical professionals including physical therapists, occupational therapists, speech language pathologists, and neuropsychologists to help deliver care focused on optimizing function and community reintegration. Given the relatively low number of physiatrists nationally, it may be difficult for patients to have access to physiatric care. According to the Association of Academic Physiatrists, there are about 10,000 practicing physiatrists to serve the needs of 61 million Americans with disabilities [1,2]. Mitsunaga et al., reported that in 2009 there was 1 physiatrist per 90,712 people in Wyoming and 1 physiatrist per 16,207 people in Washington D.C, representing the lowest and highest per capita density of physiatrists within the United States [3]. Telemedicine offers a solution that could potentially allow physiatrists to reach a larger patient population and in a more time efficient manner, thus decreasing this discrepancy long term and improving their practice during the ever-evolving challenges the COVID-19 pandemic has created regarding in-person visitation and availability.

The purpose of our research was to explore the use of telecommunication mediums by physiatrists in relation to the COVID-19 pandemic. We sought to explore if there was an increase in telehealth visits specifically among physiatrists during the COVID-19 pandemic as well as explore the potential for continued use beyond COVID-19. In doing so, we hoped to garner insights into which telecommunication mediums (phone calls, video calls, text-based communication, etc) may have expected use in the future. In addition, we sought to explore which methods of communication were preferred by physiatrists to 1) complete critical rehabilitation tasks and 2) accurately assess and manage patients in clinical practice.

## 2. Background

According to the National Institutes of Health, telemedicine is defined as “the use of electronic information and communications technologies to provide and support health care when distance separates the participants” [4]. In this study, telemedicine is the term used to cover a variety of clinical encounters that may be termed telehealth visits, teleconsultations, and telerehabilitation, delivered using 1 or more communication modes such as text, phone, and/or video. Telemedicine has the potential to improve physiatric care in many ways, including decreasing travel between rural communities and specialized urban health centers, better continuity of care to aid family support or social integration into the community, more options for patients who prefer local care, and improved access to care when extraneous events occur such as inclement weather which can prove particularly arduous for someone ambulating with prosthetics or using adaptive equipment. Studies specifically referencing physiatrist use of telemedicine indicate potential use in a variety of patient populations and visit types. A study by Savard et al. referenced the use of telemedicine via the Polycom ™ Viewstation MP VC platform in 117 patient encounters of which 38 were neurological cases. The study also presents a particular case where a physiatrist from the National Rehabilitation Hospital in Washington DC performed remote assessment and management of gait, tone, balance, cognition, dysarthria and dysphagia in a 9-year-old boy who was recovering from Dengue Fever in the US territory of American Samoa in conjunction with local therapist functioning as the primary caregiver [5]. Physiatrists have gone as far as to develop their own telemedicine platforms such as TelAbility, a video conferencing telehealth program for children with disabilities that links pediatric physiatrists with providers at early intervention centers, day cares, and private practices in rural North Carolina with favorable results. Patient family and physiatrist satisfaction and comfort, using the TelAbility system, as measured by 5-point Likert scales, were 4.91 and 4.98 respectively in 100 patient visits [6]. Similarly, a meta-analysis conducted by Cottrell using 13 studies with 1520 patients to evaluate the effectiveness of telemedicine in rehabilitation for musculoskeletal conditions found telerehab provided similar outcomes to face-to-face care with respect to care directed at physical function and disability [7].

SARS-CoV2 causing Coronavirus Infectious Disease 2019 or COVID-19 is profoundly affecting the delivery of health care globally. LM Koonin, et al [8] found a 154% increase in telemedicine visits during the last week of March 2020 compared to the same period in 2019. Notably, during the period from January to March 2020, 93% of virtually treated patients sought care for conditions other than COVID-19.

Overall, the growing adoption of technologies for telecommunication as well as pressures from the COVID-19 pandemic, continue to support the use of telemedicine and one can anticipate its use will continue to grow.

## 3. Methods

This study was conducted by analyzing survey responses from physiatrists as part of a broader effort assessing rehab care delivery, telemedicine, telepresence, and the potential of robotic interface use by various disciplines within physical medicine and rehabilitation. The primary survey was distributed to rehabilitation care providers (physiatrists, physical therapists, occupational therapists, speech language pathologists, neurologists, neurosurgeons, and hospitalists in rehab settings) through state therapy associations in the United States, Facebook groups, and direct mailing lists. Specific to the physiatrist participant cohort, physiatrists from various academic institutions within the United States as well as some of their alumni databases were invited to complete the survey. The survey, distribution plan, and general analysis plan was approved by the University of Pennsylvania Institutional Review Board. Broad data collection via REDCap [9] survey began on June 30th, 2020, and physiatrist data collection concluded on January 10th, 2021.

The survey asked several demographic questions about the responding clinician and their patients. It asked about impacts of COVID-19 on their practice of rehabilitation and their patients and included multiple questions about telemedicine and the use of telecommunication mediums such as text-based communication, phone calls, video calls, or other methods to facilitate it. The full survey can be viewed in the supplemental material. All participants were optionally invited to opt into a drawing for $20 Amazon gift cards, awarded to 1 in every 20 respondents. All survey responses where the respondent self-identified as any of the following: Physiatrist, PM&R resident, Physical Medicine and Rehabilitation, PM&R, Peds PM&R, Pediatric Rehabilitation Medicine Physician, Pediatric Physiatrist, and MD Specialty PMR were selected for analysis and non-physiatrist physician responses were disregarded (i.e. Neurosurgeon, Neurologist, Hospitalist, med consult, n= 11). Ultimately, 56 physiatrist responses were extracted and analyzed.

We hypothesized that physiatrists’ use of telecommunication would increase due to the SARS-CoV2 pandemic similarly to data previously published by Koonin et al. [8]. This was evaluated using 4 specific sub-hypotheses centered on different types of communication mediums. **H1a**: use of phone calls to patients would increase, **H1b**: use of video calls to patients at home would increase, **H1c**: use of text-based communication would increase, an **H1d**: use of video calls to patients in other clinical settings would increase. We then partitioned survey responses into telemedicine encounters that used video-based remote telecommunication (VC) and those that used non-video remote telecommunication (NVC), which included phone calls, text messages, email, instant messages, and other types of communication where physiatrists interacted with patients from afar, without using video. Hypothesis 2 (**H2**), was that COVID-19 would cause a durable change in physiatrists’ plan to use video-based telemedicine past the end of the pandemic. We also described the rehabilitation tasks that physiatrists had completed and believed could be completed during telemedicine visits using either VC or NVC methods.

Statistical analysis was performed using R [10] with the tidyverse family of packages [11] for data manipulation and for data visualization. The complete data analysis and de-identified data can be found in the supplemental material. Data for each of the **H1** sub-hypothesis was formed into contingency tables with axes: usage previous to COVID-19 and usage during COVID-19 for each mode of interaction (**H1a**: phone calls prior to vs during the pandemic, **H1b**: video calls to patients at home prior to vs during the pandemic, **H1c**: text based communication prior to vs during the pandemic, **H1d**: video calls to patients in another clinical settings prior to vs during the pandemic). To test **H1a, H1b, H1c**, and **H1d**, the paired proportion test described by Liddell [12], an exact alternative to McNemar’s test, was used, which tests the significance of the ratio of respondents who started using the medium to those who stopped using it as well as generating the confidence interval of the ratio of those who starting using the medium vs those who stopped. All tests were run with alpha set to 0.05, generating 95% confidence intervals. Cohen’s G was calculated to determine the effect size of the changes.

To evaluate **H2** (the COVID-19 pandemic led to a change in physiatrists long term plans to use telemedicine), the data was formed into a contingency table with axes: usage of video-based telemedicine prior to the onset of the COVID-19 pandemic and future (planned) usage of video-based telemedicine. Here, video-based telemedicine includes both video to patients at home and other clinical settings. The response, no future usage, included no plans for future usage and not sure about future usage, generating a conservative estimate of future usage. The same method used for testing **H1** was then applied to **H2**.

We further describe what rehabilitation tasks were amenable to or not amendable to telemedicine using either VC or NVC communication modes.

## 4. Results

### 4.1. Survey Demographics

The responses analyzed came from 56 physiatrists of whom 50% are female. The youngest physiatrist was 26 and eldest was 60 with a mean age of 35.29 (SD=7.9 years). 89% percent (50/56) of respondents reported they are from the United States, with the remaining 6 coming from Mexico (1), Brazil (1), Costa Rica (2), Honduras (1), and Turkey (1). Of those practicing in the United States, 29 are from Pennsylvania, 8 from New York, 3 from Illinois, 2 from California, 2 from New Jersey, 2 from North Carolina, 2 from Ohio, 1 from Oregon, and 1 from Massachusetts. Respondents had, on average, 6.19 years (SD=6.97) of experience with a minimum of 1 years and maximum of 35 years. Respondents practice in a variety of settings including 42 in a “rehab center”, 33 in an “outpatient facility”, 26 in an “inpatient facility”, 20 in a “general hospital”, 6 in a “hospital for children”, and 4 in a “private practice”. They practice across areas with varying population densities with 49 practicing in an urban setting, 12 in a suburban setting, and 2 in a rural setting. The type of pathologies treated by the physiatrists (labeled “patient types” in the survey) include traumatic brain injury (35), stroke (42), dementia (13), sports injuries (30), arthritis (35), general traumatic injuries (34), amputations (35), cardiovascular disease (19), mental health issues (11), burns (6), autism (5), cerebral palsy (27), speech disorders (19), and others. The respondents treat patients across the spectrum from not cognitively impaired to severely cognitively impaired and from not motor impaired to severely motor impaired. Respondents treat patients of all ages: adults 65 and older (49), middle aged adults 40–65 years old (50), adults aged 21–40 (48), young adults aged 18–21 (29), teens aged 12–18 (12), grade schoolers aged 5–12 (6), preschoolers aged 3–5 (6), toddlers aged 1–3 (6), and infants aged 0–1 years (5).

### 4.2. Use of Telemedicine for Care by Physiatrists Before, During, and After COVID-19

A summary of the change in usage of telemedicine from prior to the pandemic to during the pandemic is shown in fig. 1. There was a statistically significant increase in the use of phone calls with patients for care delivery among physiatrists during COVID-19 (**H1a**) (p=0.012; CI 1.42–433.98) with a large effect (Cohen’s g= 0.41). Ten (10) respondents who did not previously use phone calls with patients began using them during COVID, while 1 respondent who has previously used phone calls stopped using them. Forty-one (41) respondents who were previously using phone calls continued to use them. Four (4) who were not previously using phone calls with patients continued to not use them. There was also a statistically significant change in the utilization of video calls to patients at home during the COVID-19 pandemic (**H1b**) (p= 0.0001; CI 2.98–105) with a large effect (Cohen’s g=0.423). Twenty-four (24) physiatrists who were not previously making video calls to patients at home started making them. Two (2) physiatrists who were previously using video calls to home stopped using them. Eleven (11) physiatrists who were not and 19 who were making video calls to patients at home did not change their usage of video calls to home. The use of text-based communication did not increase significantly (**H1c**) (p=1.000). Three (3) physiatrists who had not previously used text-based communication to communicate with patients started using it and 2 who had previously used it stopped using it. Thirty-six (36) physiatrists who were not using text-based communication and 15 who were did not change their usage. There was a significant increase in video calls to another clinical setting (**H1d**) (p=0.016; CI 1.44+) with a large effect (Cohen’s g=0.5). An upper bound for the confidence interval could not be found because no respondents who were using video calls to another setting stopped doing so during the pandemic. Seven (7) physiatrists who had not previously used video to other clinical settings began doing so during the pandemic. Forty-five (45) physiatrists who had not been and 4 who had been using video to other clinical settings did not change their usage. Hypothesis 2 (**H2**) assessed whether physiatrists planned usage of VC for telemedicine post-pandemic exceeded their pre-pandemic usage (table 1). Prior to COVID-19, 37.5% (21/56) of respondents were using video to connect with patients at either their homes or another clinical setting and 78.6% (44/56) planned to use it after the pandemic has ended. Making, for the purpose of analysis, the conservative assumption that respondents who were unsure about future usage of VC (9) would in fact not use it: 7 had not and did not plan to use video-based remote communication, 5 had used VC prior to the pandemic but did not plan to post pandemic, 28 had not used VC prior to the pandemic but planned to post-pandemic, and 16 had used VC prior to the pandemic and intended to continue to do so. This change in video communication usage from prior to the pandemic to post pandemic (anticipated), is statistically significant (p= 0.000066; CI 2.13–18.6) with large effect size (Cohen’s g=0.348).

**Table 1.** Usage of Video Based Telemedicine to Patients at Home and Other Clinical Settings, Before Pandemic, During Pandemic, and Planned Usage After Pandemic

| Usage Pre-COVID | Usage During COVID | Future Usage |  |  |
| --- | --- | --- | --- | --- |
|  |  | Yes | No | Not Sure |
| No | No | 7 | 1 | 3 |
|  | Yes | 21 | 0 | 3 |
| Yes | No | 1 | 0 | 1 |
|  | Yes | 15 | 2 | 2 |

**Figure 1.**
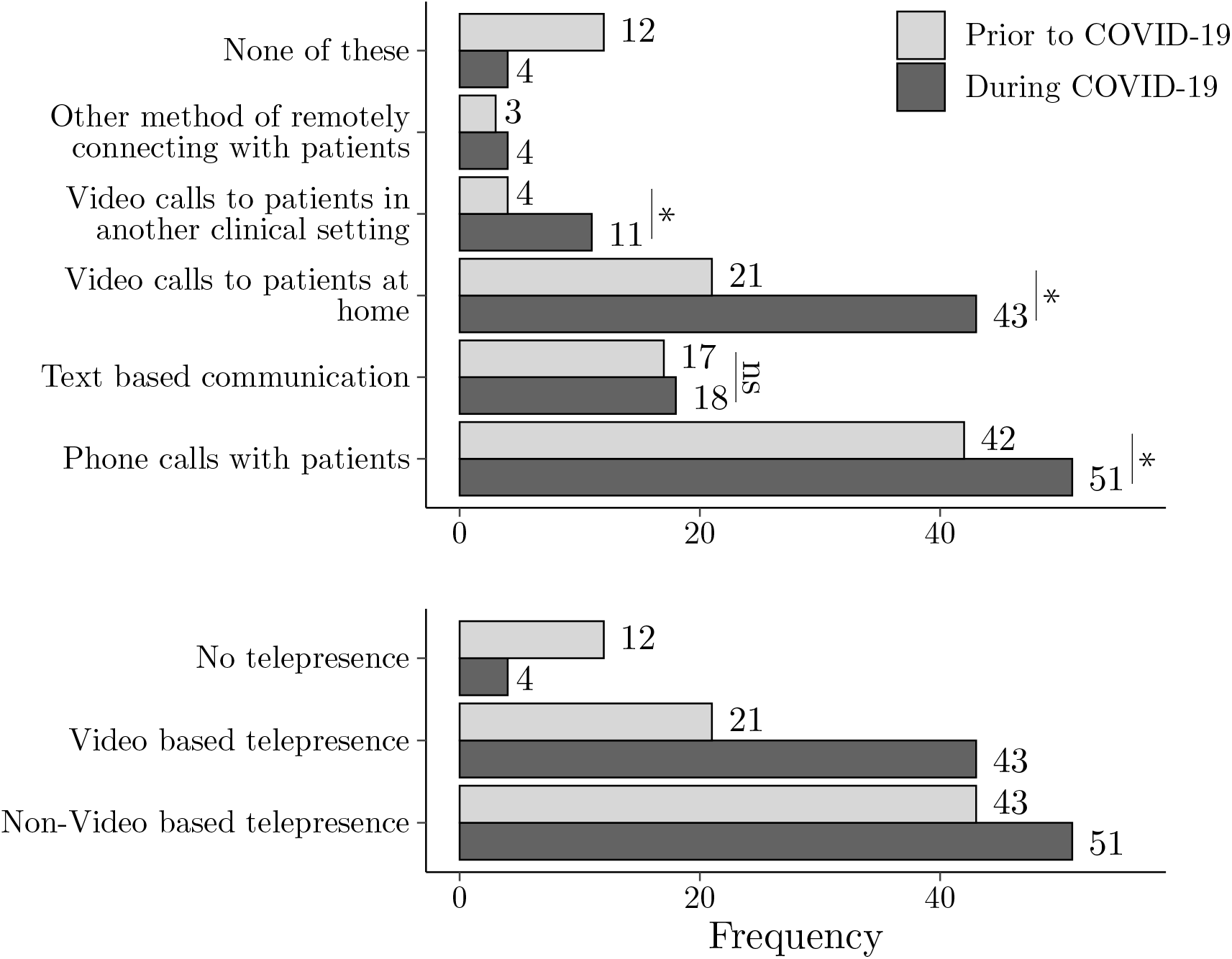
On top: Usage of various modalities for remotely connecting with patients which physiatrists in the study were using prior to and during the COVID-19 pandemic. Significant changes marked with “*”, tested but non-significant changes marked with “ns”. On bottom: The same data compressed into no telepresence, videobased telepresence, and non-video-based telepresence. None of these/No telepresence are exclusive categories, the other categories are not.

### 4.3. Activities Which Physiatrists Believe Can Be Accomplished Via Telemedicine

The activities which physiatrists have done/believe can be done using different modalities of communication are shown in table 2, along with which activities they have done in person. There were notable differences in responses to what physiatrists “have done” compared with what they thought they “could do” regarding orthotic assessments, motor assessments, activities of daily living (ADL) practice, cognitive assessment, cognitive exercises, stretching, strength building, and environmental adaptation. Motor assessments and cognitive assessments stand out as more than 70% of the physiatrists have done these activities in person, indicating their importance to practice.

**Table 2.** Rehab Activities Which Respondents Have Done (HD) In Person, Could Do (CD) and Have Done with Non-Video Remote Communication (NVC), Could Do and Have Done with Video Based Remote Communication (VC), Have Not Done in Any Form (HND) and Could Not Be Done Remotely (CNBDR). Rows ordered by number of physiatrists who have done the activity in person.

|  | In Person<br>HD | NVC |  | VC |  | HND | CNBDR |
| --- | --- | --- | --- | --- | --- | --- | --- |
|  |  | CD | HD | CD | HD |  |  |
| Motor Assessments | 49 | 4 | 3 | 41 | 36 | 2 | 10 |
| Medical Prescriptions | 45 | 40 | 38 | 51 | 39 | 1 | 2 |
| Discussions about Radiology Results | 42 | 37 | 30 | 52 | 35 | 5 | 1 |
| Cognitive Assessments | 41 | 24 | 10 | 53 | 31 | 9 | 1 |
| Discussions about Surgery | 39 | 37 | 21 | 52 | 30 | 11 | 1 |
| Stretching | 37 | 7 | 4 | 41 | 20 | 16 | 11 |
| Orthotics Assessment/Prescription | 33 | 8 | 5 | 36 | 16 | 20 | 20 |
| Strength Building | 32 | 9 | 4 | 44 | 13 | 21 | 9 |
| ADL Practice | 21 | 3 | 3 | 46 | 8 | 32 | 6 |
| Cognitive Exercises | 16 | 19 | 3 | 52 | 9 | 36 | 2 |
| Environmental Adaptation | 16 | 6 | 4 | 41 | 11 | 36 | 10 |

There are also several activities which the physiatrists both believe can be done remotely without video and have experience doing remotely. Of note, more than 65% of respondents believe they could do medical prescriptions, discussions about radiology results, and discussions about surgery.

The results for motor assessments via VC with 36 responses for “have done” and 41 for “could do” compared to (NVC) with only 3 responses for “have done” and 4 for “could do” suggest that video is preferable for this activity among physiatrists. Similarly, ADL practice with VC had 8 responses for “have done” and 46 responses for “could do”, compared to NVC with 3 for “have done” and 3 for “could do”.

## 5. Discussion

The pressures of care delivery during the pandemic have necessitated forms of patient interaction beyond in-person visits and thus, in line with results from other studies, our results corroborate the increased use of telecommunication by physiatrists during the COVID-19 pandemic. This was consistent with the findings in a study performed by Escalon in April 2020 which found an increase of telehealth usage from 16.6% prior to the pandemic to 86.5% out of 178 attending PM&R physicians [13].

Our results confirm that telemedicine usage, both by video and phone, was significantly increased during the early portion of the COVID-19 pandemic, compared to the pre-pandemic period, and that most physiatrists plan to continue to use it after the pandemic, representing a significant increase in video-based telepresence usage post-pandemic, compared to pre-pandemic. From a patient’s perspective, in a study conducted by Brennan et al., 70% of those who used telerehabilitation amid the pandemic reported to be likely to use it after the pandemic has ended [14] which suggests that continued implementation by physiatrists will be welcomed by patients.

The data further highlights activities which provide particular opportunities for the use of video-based telemedicine. Notably motor assessments, cognitive assessments, stretching, strength building, and orthotics assessment and prescription have been done in person by the majority of the study cohort and 2–10 times more respondents feel that they can be done using video than with non-video telemedicine.

Establishing which types of clinician activities require video features versus which can be accomplished without video may help promote efficiency and effectiveness of assessments and interventions going forward. Resource utilization could be stratified or triaged in a manner that could promote better allocation of resources and videoconferencing platforms. This has potential to optimize patient evaluation in both the inpatient and outpatient settings. For example, if a short pre-consultation survey or simple data gathering is needed to determine chief complaint or issue, it may be possible to determine if video-enabled technology is needed to address or further triage the patient issue at hand. If the need for assessment of motor function is established, then protocols could be set forth that this type of evaluation requires video capability for effective assessment. Similarly, if prosthetic or orthotic needs are determined, video technology would likely be required.

The emphasis on direct visualization for certain rehabilitation tasks is due to inherent complexity of physical assessment. While hands-on evaluation remains superior, being able to visualize extremity movement can help distinguish nuances in assessment. For example, if a patient or caregiver verbally reports that the patient cannot raise their arms above their head, there are many questions as to why this is not possible. For example, is this due to weakness, joint contracture, muscle spasticity, or motor control or is it just poor activation, participation, or a proprioceptive deficit? With video telecommunication, these types of questions can be more easily answered. On the other hand, VC or NVC would likely be acceptable for the activities of discussion of surgery, discussion of radiology, and medical prescriptions where there were high response rates for both VC and NVC for “have done” and “could do”. In addition, these activities had low “could not be done remotely” responses further supporting the utility of telehealth mediums for these activities.

### 5.1. Study Limitations

This study has several limitations. The study has a relatively small sample size of 56 with most responses from the United States, specifically providers practicing in urban settings in Pennsylvania which limits its generalizability. As it was part of a broader focused study also assessing therapist considerations as well as use of the socially interactive robotic platform Flo, survey questions were not targeting the physiatrist cohort directly and more targeted questions could have been used. The chosen statistical analysis, using the method provided by Liddell, is a test of marginal homogeneity, which ignores the subjects who fall on the diagonal of a contingency table (did not change their value over the condition boundary; ex: used telepresence before and during the pandemic). As such, some portion of the data is not considered when testing significance. Despite these limitations, our research provides insight on how physiatrists responded to the pandemic with greater use of telemedicine, of the perceived capabilities of currently used telemedicine methods, and physiatrists disposition to implementing telemedicine post-pandemic.

## 6. Conclusion

Telemedicine use has increased as technology has improved. The COVID-19 pandemic has added external pressures which have contributed to increased use of technology to allow medical care delivery. Physiatrists have adopted this technology and the continued use of telemedicine is expected by physiatrists. Certain clinical activities such as motor assessments, cognitive assessments, and stretching may require video based remote telecommunication whereas others such as discussions on surgery, radiology results and medication prescriptions may not. Further research is needed to formalize guidelines for how, when, and for what telemedicine should be used, this may help pave the way for better allocation of resources and better delivery of telerehabilitation.

## Data Availability

All data produced in the present work are contained in the supplemental material

## Abbreviations

PM&R: Physical medicine and rehabilitation
ADL: Activities of daily living

## Acknowledgement(s)

We want to thank all the physiatrists who completed our survey. We also want to thank the professional societies, Facebook community groups, and others who were instrumental in recruiting subjects for the survey.

## Data Availability Statement

The authors confirm that the data supporting the findings of this study are available within the article [and/or] its supplemental material.

