## Supplementary material for "Insights on Telemedicine Use by Physiatrists Before, During, and Beyond the COVID-19 Pandemic": physiatrists-covid-data_export.R

```
library(dplyr)

### LOAD DATA ###

# Clear existing data and graphics
# rm(list=ls())
# graphics.off()

# Load Hmisc library
library(Hmisc)

# Read Data ----
data <- read.csv("./data/FloClinicianSurveys_DATA.csv")

# Setting Labels ----

label(data$record_id) <- "Record ID"
label(data$clinician_survey_timestamp) <- "Survey Timestamp"
label(data$occupation) <- "Are you:"
label(data$occupation_therapist_type) <- "What type of therapist are you?"
label(data$occupation_nurse_type) <- "What type of nurse are you?"
label(data$occupation_doctor_type) <- "What type of doctor are you?"
label(data$occupation_medtech_type) <- "What type of medical technologist are you?"
label(data$occupation_other_type) <- "What type of clinician are you?"
label(data$age) <- "What is your age (years)"
label(data$gender) <- "What is your gender?"
label(data$location___1) <- "School"
label(data$location___2) <- "Hospital for Children"
label(data$location___3) <- "General Hospital"
label(data$location___4) <- "Elder Care Hospital"
label(data$location___5) <- "Rehab Center"
label(data$location___6) <- "Elder Care Home"
label(data$location___7) <- "Community Center"
label(data$location___8) <- "Private Practice"
label(data$location___9) <- "Patient Home"
label(data$location___10) <- "Inpatient Facility"
label(data$location___11) <- "Outpatient Facility"
label(data$location___12) <- "Other"
label(data$location_other) <- "Please specify your work location:"
label(data$location_type___1) <- "Urban"
label(data$location_type___2) <- "Suburban"
label(data$location_type___3) <- "Rural"
label(data$location_country) <- "What country do you primarily work in? "
label(data$location_state) <- "What state do you primarily work in?"
label(data$years_experience) <- "How many years have you been practicing?"
label(data$patient_pop___1) <- "CP"
label(data$patient_pop___2) <- "Stroke"
label(data$patient_pop___3) <- "Brachial Plexus Injuries"
label(data$patient_pop___4) <- "Autism"
label(data$patient_pop___5) <- "TBI"
label(data$patient_pop___6) <- "Cardiovascular Disease"
label(data$patient_pop___7) <- "Speech Disorders"
label(data$patient_pop___8) <- "Sports Injuries"
label(data$patient_pop___9) <- "Amputations"
label(data$patient_pop___10) <- "Arthritis"
label(data$patient_pop___11) <- "Dementia"
label(data$patient_pop___12) <- "Burns"
label(data$patient_pop___13) <- "General Traumatic Injuries"
label(data$patient_pop___15) <- "Mental Health Issues"
label(data$patient_pop___14) <- "Other"
label(data$patient_pop_other) <- "Please describe the other patient types who you treat:"
label(data$patient_pop_cog___1) <- "Severely Impaired"
label(data$patient_pop_cog___2) <- "Moderately Impaired"
label(data$patient_pop_cog___3) <- "Mildly Impaired"
label(data$patient_pop_cog___4) <- "Not Impaired"
label(data$patient_pop_motor___1) <- "Severely Impaired"
label(data$patient_pop_motor___2) <- "Moderately Impaired"
label(data$patient_pop_motor___3) <- "Mildly Impaired"
label(data$patient_pop_motor___4) <- "Not Impaired"
label(data$patient_pop_age___1) <- "Infants (0-1)"
label(data$patient_pop_age___2) <- "Toddlers (1-3)"
label(data$patient_pop_age___3) <- "Preschoolers (3-5)"
label(data$patient_pop_age___4) <- "Gradeschoolers (5-12)"
label(data$patient_pop_age___5) <- "Teens (12-18)"
label(data$patient_pop_age___6) <- "Young Adults (18-21)"
label(data$patient_pop_age___7) <- "Adults (21-40)"
label(data$patient_pop_age___8) <- "Middle Aged Adults (40-65)"
label(data$patient_pop_age___9) <- "Older Adults (65+)"
label(data$patient_pop_phase___1) <- "Acute"
label(data$patient_pop_phase___2) <- "Sub-Acute"
label(data$patient_pop_phase___3) <- "Chronic"
label(data$past_telepresence___1) <- "Phone calls with patients"
label(data$past_telepresence___2) <- "Text based communication"
label(data$past_telepresence___3) <- "Video calls to patients at home"
label(data$past_telepresence___4) <- "Video calls to patients in another clinical setting"
label(data$past_telepresence___5) <- "Other method of remotely connecting with patients"
label(data$past_telepresence___6) <- "None of these"
label(data$past_telepresence_other) <- "What is the other method you have used?"
label(data$covid_inperson) <- "Have you been able to see your patients in-person during the pandemic?"
label(data$covid_treatment_difficulty) <- "How has the pandemic changed your level of interaction with your patients?"
label(data$covid_outcomes) <- "How do you expect the pandemic to affect your patients long term health outcomes?"
label(data$covid_patient_selfcare) <- "How do you believe the pandemic is affecting your patients ability to care for themselves and meet their rehabilitation goals? "
label(data$covid_telepresence___1) <- "Phone calls with patients"
label(data$covid_telepresence___2) <- "Text based communication"
label(data$covid_telepresence___3) <- "Video calls to patients at home"
label(data$covid_telepresence___4) <- "Video calls to patients in another clinical setting"
label(data$covid_telepresence___5) <- "Other method of remotely connecting with patients"
label(data$covid_telepresence___6) <- "None of these"
label(data$covid_telepresence_other) <- "What is the other method you are using?"
label(data$video_plan) <- "Do you plan to use video calls to talk to your patients going forward during the COVID-19 pandemic?"
label(data$video_satisfaction) <- "Please rate your overall SATISFACTION using video calls for rehab"
label(data$video_communication) <- "How well are you able to COMMUNICATE with your patients over video calls compared to in-person?"
label(data$pvideo_communication) <- "How well do you believe you could COMMUNICATE with your patients over video calls compared to in-person?"
label(data$video_assessment) <- "How well are you able to ASSESS your patients level of function over video calls compared to in-person?"
label(data$pvideo_assessment) <- "How well do you believe you could ASSESS your patients level of function over video calls compared to in-person?"
label(data$video_motivation) <- "How MOTIVATED are your patients DURING a video call compared to in-person?"
label(data$pvideo_motivation) <- "How MOTIVATED do you believe your patients would be DURING a video call compared to in-person?"
label(data$video_compliance_during) <- "How well do your patients COMPLY with instructions DURING a video call vs in-person visit?"
label(data$pvideo_compliance_during) <- "How well do you believe your patients would COMPLY with instructions DURING a video call vs in-person visit?"
label(data$video_adherence_after) <- "How well do your patients ADHERE to the treatment plan AFTER a video call vs in-person visit?"
label(data$pvideo_adherence_after) <- "How well do you believe your patients would ADHERE to the treatment plan AFTER a video call vs in-person visit?"
label(data$video_challenges___1) <- "No challenges"
label(data$video_challenges___2) <- "connection failures"
label(data$video_challenges___3) <- "poor audio quality"
label(data$video_challenges___4) <- "poor video quality"
label(data$video_challenges___5) <- "Patients do not have a device for video"
label(data$video_challenges___6) <- "Patients do not have internet"
label(data$video_challenges___7) <- "Patients cannot understand what I want"
label(data$video_challenges___8) <- "I cannot understand what patients want"
label(data$video_challenges___9) <- "Patients cannot complete activities without my physical intervention"
label(data$video_challenges___10) <- "Assessments require the use of a tool which the patient does not have"
label(data$video_challenges___11) <- "It is not safe for patients to do the desired activities without a professional present"
label(data$video_challenges___12) <- "Other challenges"
label(data$pvideo_challenges___1) <- "No challenges"
label(data$pvideo_challenges___2) <- "Connection failures"
label(data$pvideo_challenges___3) <- "Poor audio quality"
label(data$pvideo_challenges___4) <- "Poor video quality"
label(data$pvideo_challenges___5) <- "Patients would not have a device for video"
label(data$pvideo_challenges___6) <- "Patients would not have internet"
label(data$pvideo_challenges___7) <- "Patients would not understand what I want"
label(data$pvideo_challenges___8) <- "I would not understand what patients want"
label(data$pvideo_challenges___9) <- "Patients could not complete activities without my physical intervention"
label(data$pvideo_challenges___10) <- "Assessments require the use of a tool which the patient would not have"
label(data$pvideo_challenges___11) <- "It would not be safe for patients to do the desired activities without a professional present"
label(data$pvideo_challenges___12) <- "Other challenges"
label(data$video_challenges_other) <- "What other challenges?"
label(data$video_future) <- "Do you plan to use video calls with your patients for your practice after the COVID-19 pandemic has ended?"
label(data$flo_prior_experience___1) <- "No prior knowledge"
label(data$flo_prior_experience___2) <- "I have read a paper on the system"
label(data$flo_prior_experience___3) <- "I have seen the system in person"
label(data$flo_prior_experience___4) <- "I have used the system"
label(data$flo_prior_experience___5) <- "I have some other experience with system"
label(data$flo_interest) <- "How interested would you be in using the LilFlo system?"
label(data$flo_location___1) <- "Rural outpatient clinics"
label(data$flo_location___2) <- "Rural inpatient clinics"
label(data$flo_location___3) <- "Elder care facilities"
label(data$flo_location___4) <- "Schools"
label(data$flo_location___5) <- "Patient homes"
label(data$flo_location___6) <- "Community centers"
label(data$flo_location___7) <- "Urban inpatient clinics"
label(data$flo_location___8) <- "Urban outpatient clinics"
label(data$flo_location___9) <- "None"
label(data$flo_location___10) <- "Other"
label(data$flo_location_other) <- "What other locations?"
label(data$flo_communication) <- "COMMUNICATION during the interaction"
label(data$flo_motivation) <- "Patient MOTIVATION during the interaction"
label(data$flo_assessment) <- "Your ability to ASSESS your patients from telepresence interactions"
label(data$flo_compliance) <- "How well your patients COMPLY with instructions DURING the telepresence interaction"
label(data$flo_adherence) <- "How well your patients ADHERE to the treatment plan AFTER a telepresence interaction"
label(data$telemed_sys_feat_mobile) <- "A mobile system which can be driven remotely"
label(data$telemed_sys_feat_autodrive) <- "A mobile system which can drive on its own"
label(data$telemed_sys_feat_arms) <- "A social robot with arms to augment standard video calls"
label(data$telemed_sys_feat_face) <- "A social robot with an expressive face to augment standard video calls"
label(data$telemed_sys_feat_autointer) <- "A social robot that can interact with patients without needing operator input"
label(data$telemed_sys_feat_web) <- "A web interface for controlling remote telerehabilitation systems"
label(data$telemed_sys_feat_games) <- "A social robot which can play games with subjects during telerehabilitation calls to augment standard video"
label(data$telemed_sys_feat_asses) <- "A system which collects data and performs automated assessments of patient function"
label(data$telemed_sys_feat_screen) <- "A clear screen to see the clinician"
label(data$telemed_sys_feat_vid) <- "High quality video to see the patient"
label(data$flo_features_other) <- "Please list any other features which you believe would be useful."
label(data$activities_d_motor_ass___1) <- "Non-Video Remote Interaction"
label(data$activities_d_motor_ass___3) <- "Video Call Based Remote Interaction"
label(data$activities_d_motor_ass___4) <- "LilFlo Based Remote Interaction"
label(data$activities_d_motor_ass___5) <- "In Person"
label(data$activities_d_motor_ass___6) <- "Have Not Done"
label(data$activities_d_stretching___1) <- "Non-Video Remote Interaction"
label(data$activities_d_stretching___3) <- "Video Call Based Remote Interaction"
label(data$activities_d_stretching___4) <- "LilFlo Based Remote Interaction"
label(data$activities_d_stretching___5) <- "In Person"
label(data$activities_d_stretching___6) <- "Have Not Done"
label(data$activities_d_strength___1) <- "Non-Video Remote Interaction"
label(data$activities_d_strength___3) <- "Video Call Based Remote Interaction"
label(data$activities_d_strength___4) <- "LilFlo Based Remote Interaction"
label(data$activities_d_strength___5) <- "In Person"
label(data$activities_d_strength___6) <- "Have Not Done"
label(data$activities_d_adl___1) <- "Non-Video Remote Interaction"
label(data$activities_d_adl___3) <- "Video Call Based Remote Interaction"
label(data$activities_d_adl___4) <- "LilFlo Based Remote Interaction"
label(data$activities_d_adl___5) <- "In Person"
label(data$activities_d_adl___6) <- "Have Not Done"
label(data$activities_d_cog_ass___1) <- "Non-Video Remote Interaction"
label(data$activities_d_cog_ass___3) <- "Video Call Based Remote Interaction"
label(data$activities_d_cog_ass___4) <- "LilFlo Based Remote Interaction"
label(data$activities_d_cog_ass___5) <- "In Person"
label(data$activities_d_cog_ass___6) <- "Have Not Done"
label(data$activites_d_cog_ex___1) <- "Non-Video Remote Interaction"
label(data$activites_d_cog_ex___3) <- "Video Call Based Remote Interaction"
label(data$activites_d_cog_ex___4) <- "LilFlo Based Remote Interaction"
label(data$activites_d_cog_ex___5) <- "In Person"
label(data$activites_d_cog_ex___6) <- "Have Not Done"
label(data$activities_d_env_adap___1) <- "Non-Video Remote Interaction"
label(data$activities_d_env_adap___3) <- "Video Call Based Remote Interaction"
label(data$activities_d_env_adap___4) <- "LilFlo Based Remote Interaction"
label(data$activities_d_env_adap___5) <- "In Person"
label(data$activities_d_env_adap___6) <- "Have Not Done"
label(data$activities_d_orthotics___1) <- "Non-Video Remote Interaction"
label(data$activities_d_orthotics___3) <- "Video Call Based Remote Interaction"
label(data$activities_d_orthotics___4) <- "LilFlo Based Remote Interaction"
label(data$activities_d_orthotics___5) <- "In Person"
label(data$activities_d_orthotics___6) <- "Have Not Done"
label(data$activities_d_surg___1) <- "Non-Video Remote Interaction"
label(data$activities_d_surg___3) <- "Video Call Based Remote Interaction"
label(data$activities_d_surg___4) <- "LilFlo Based Remote Interaction"
label(data$activities_d_surg___5) <- "In Person"
label(data$activities_d_surg___6) <- "Have Not Done"
label(data$activities_d_rads___1) <- "Non-Video Remote Interaction"
label(data$activities_d_rads___3) <- "Video Call Based Remote Interaction"
label(data$activities_d_rads___4) <- "LilFlo Based Remote Interaction"
label(data$activities_d_rads___5) <- "In Person"
label(data$activities_d_rads___6) <- "Have Not Done"
label(data$activities_d_med___1) <- "Non-Video Remote Interaction"
label(data$activities_d_med___3) <- "Video Call Based Remote Interaction"
label(data$activities_d_med___4) <- "LilFlo Based Remote Interaction"
label(data$activities_d_med___5) <- "In Person"
label(data$activities_d_med___6) <- "Have Not Done"
label(data$activities_b_motor_ass___1) <- "Non-Video Remote Interaction"
label(data$activities_b_motor_ass___3) <- "Video Call Based Remote Interaction"
label(data$activities_b_motor_ass___4) <- "LilFlo Based Remote Interaction"
label(data$activities_b_motor_ass___5) <- "Could Not Be Done Remotely"
label(data$activities_b_stretching___1) <- "Non-Video Remote Interaction"
label(data$activities_b_stretching___3) <- "Video Call Based Remote Interaction"
label(data$activities_b_stretching___4) <- "LilFlo Based Remote Interaction"
label(data$activities_b_stretching___5) <- "Could Not Be Done Remotely"
label(data$activities_b_strength___1) <- "Non-Video Remote Interaction"
label(data$activities_b_strength___3) <- "Video Call Based Remote Interaction"
label(data$activities_b_strength___4) <- "LilFlo Based Remote Interaction"
label(data$activities_b_strength___5) <- "Could Not Be Done Remotely"
label(data$activities_b_adl___1) <- "Non-Video Remote Interaction"
label(data$activities_b_adl___3) <- "Video Call Based Remote Interaction"
label(data$activities_b_adl___4) <- "LilFlo Based Remote Interaction"
label(data$activities_b_adl___5) <- "Could Not Be Done Remotely"
label(data$activities_b_cog_ass___1) <- "Non-Video Remote Interaction"
label(data$activities_b_cog_ass___3) <- "Video Call Based Remote Interaction"
label(data$activities_b_cog_ass___4) <- "LilFlo Based Remote Interaction"
label(data$activities_b_cog_ass___5) <- "Could Not Be Done Remotely"
label(data$activites_b_cog_ex___1) <- "Non-Video Remote Interaction"
label(data$activites_b_cog_ex___3) <- "Video Call Based Remote Interaction"
label(data$activites_b_cog_ex___4) <- "LilFlo Based Remote Interaction"
label(data$activites_b_cog_ex___5) <- "Could Not Be Done Remotely"
label(data$activities_b_env_adap___1) <- "Non-Video Remote Interaction"
label(data$activities_b_env_adap___3) <- "Video Call Based Remote Interaction"
label(data$activities_b_env_adap___4) <- "LilFlo Based Remote Interaction"
label(data$activities_b_env_adap___5) <- "Could Not Be Done Remotely"
label(data$activities_b_orthotics___1) <- "Non-Video Remote Interaction"
label(data$activities_b_orthotics___3) <- "Video Call Based Remote Interaction"
label(data$activities_b_orthotics___4) <- "LilFlo Based Remote Interaction"
label(data$activities_b_orthotics___5) <- "Could Not Be Done Remotely"
label(data$activities_b_surg___1) <- "Non-Video Remote Interaction"
label(data$activities_b_surg___3) <- "Video Call Based Remote Interaction"
label(data$activities_b_surg___4) <- "LilFlo Based Remote Interaction"
label(data$activities_b_surg___5) <- "Could Not Be Done Remotely"
label(data$activities_b_rads___1) <- "Non-Video Remote Interaction"
label(data$activities_b_rads___3) <- "Video Call Based Remote Interaction"
label(data$activities_b_rads___4) <- "LilFlo Based Remote Interaction"
label(data$activities_b_rads___5) <- "Could Not Be Done Remotely"
label(data$activities_b_med___1) <- "Non-Video Remote Interaction"
label(data$activities_b_med___3) <- "Video Call Based Remote Interaction"
label(data$activities_b_med___4) <- "LilFlo Based Remote Interaction"
label(data$activities_b_med___5) <- "Could Not Be Done Remotely"
label(data$robot_feelings_positive) <- "I have general positive feelings towards robots"
label(data$robot_feelings_same) <- "Robots are the same as any other medical/assistive devices"
label(data$robot_feelings_indep) <- "Robots will improve patient independence"
label(data$robot_feelings_focus) <- "Robots allow other caretakers to focus on higher level work"
label(data$robot_feelings_accurate) <- "Robots are consistent and accurate"
label(data$robot_feelings_expensive) <- "Robots are too expensive"
label(data$robot_feelings_time) <- "Robots take too much caretaker time to set up"
label(data$robot_feelings_unethical) <- "The use of robots for care and treatment is unethical"
label(data$robot_feelings_patwant) <- "Patients dont want robots to take care of them"
label(data$robot_feelings_control) <- "I want robots that I can control"
label(data$robot_feelings_jobs) <- "Robots are going to take jobs"
label(data$robot_feelings_fail) <- "Robots are too likely to break or fail"
label(data$robot_feelings_clinfeel) <- "Robots will improve clinician well-being"
label(data$robot_feelings_neg) <- "I have general negative feelings towards robots"
label(data$robot_feelings_failethic) <- "Failing to use robots for care and treatment is unethical"
label(data$robot_feelings_interact) <- "Using robots will damage peoples ability to interact with other people"
label(data$robot_feelings_laws) <- "We need more laws and regulations about robots"
label(data$robot_feelings_study) <- "We need to study more about how robots affect function"
label(data$robot_feelings_autonomy) <- "I want robots that act on their own"
label(data$robot_feelings_bad) <- "Robots are bad for society"
label(data$robot_feelings_other) <- "What other feelings do you have towards robots?"
label(data$final_other) <- "If you have any other general thoughts you would like to share with the community about COVID-19s impacts on your practice and patients, telemedicine/telepresence, LilFlo, etc. Please share them here. This is the final question of the survey. "
label(data$giftcard) <- "Would you like to be entered into the drawing for a $20 Amazon gift card?"
label(data$results) <- "Would you like to be made aware of the results of this survey when they are published?"
label(data$clinician_survey_complete) <- "Complete?"

# Setting Factors(will create new variable for factors) ----
data$occupation.factor <- factor(data$occupation, levels = c("1", "2", "3", "4", "5"))
data$gender.factor <- factor(data$gender, levels = c("1", "2", "3"))
data$location___1.factor <- factor(data$location___1, levels = c("0", "1"))
data$location___2.factor <- factor(data$location___2, levels = c("0", "1"))
data$location___3.factor <- factor(data$location___3, levels = c("0", "1"))
data$location___4.factor <- factor(data$location___4, levels = c("0", "1"))
data$location___5.factor <- factor(data$location___5, levels = c("0", "1"))
data$location___6.factor <- factor(data$location___6, levels = c("0", "1"))
data$location___7.factor <- factor(data$location___7, levels = c("0", "1"))
data$location___8.factor <- factor(data$location___8, levels = c("0", "1"))
data$location___9.factor <- factor(data$location___9, levels = c("0", "1"))
data$location___10.factor <- factor(data$location___10, levels = c("0", "1"))
data$location___11.factor <- factor(data$location___11, levels = c("0", "1"))
data$location___12.factor <- factor(data$location___12, levels = c("0", "1"))
data$location_type___1.factor <- factor(data$location_type___1, levels = c("0", "1"))
data$location_type___2.factor <- factor(data$location_type___2, levels = c("0", "1"))
data$location_type___3.factor <- factor(data$location_type___3, levels = c("0", "1"))
data$location_country.factor <- factor(data$location_country, levels = c("1", "2", "3", "4", "5", "6", "7", "8", "9", "10", "11", "12", "13", "14", "15", "16", "17", "18", "19", "20", "21", "22", "23", "24", "25", "26", "27", "28", "29", "30", "31", "32", "33", "34", "35", "36", "37", "38", "39", "40", "41", "42", "43", "44", "45", "46", "47", "48", "49", "50", "51", "52", "53", "54", "55", "56", "57", "58", "59", "60", "61", "62", "63", "64", "65", "66", "67", "68", "69", "70", "71", "72", "73", "74", "75", "76", "77", "78", "79", "80", "81", "82", "83", "84", "85", "86", "87", "88", "89", "90", "91", "92", "93", "94", "95", "96", "97", "98", "99", "100", "101", "102", "103", "104", "105", "106", "107", "108", "109", "110", "111", "112", "113", "114", "115", "116", "117", "118", "119", "120", "121", "122", "123", "124", "125", "126", "127", "128", "129", "130", "131", "132", "133", "134", "135", "136", "137", "138", "139", "140", "141", "142", "143", "144", "145", "146", "147", "148", "149", "150", "151", "152", "153", "154", "155", "156", "157", "158", "159", "160", "161", "162", "163", "164", "165", "166", "167", "168", "169", "170", "171", "172", "173", "174", "175", "176", "177", "178", "179", "180", "181", "182", "183", "184", "185", "186", "187", "188", "Venezuela", "189", "190", "191", "192"))
data$location_state.factor <- factor(data$location_state, levels = c("1", "2", "3", "4", "5", "6", "7", "8", "9", "10", "11", "12", "13", "14", "15", "16", "17", "18", "19", "20", "21", "22", "23", "24", "25", "26", "27", "28", "29", "30", "31", "32", "33", "34", "35", "36", "37", "38", "39", "40", "41", "42", "43", "44", "45", "46", "47", "48", "49", "50", "51"))
data$patient_pop___1.factor <- factor(data$patient_pop___1, levels = c("0", "1"))
data$patient_pop___2.factor <- factor(data$patient_pop___2, levels = c("0", "1"))
data$patient_pop___3.factor <- factor(data$patient_pop___3, levels = c("0", "1"))
data$patient_pop___4.factor <- factor(data$patient_pop___4, levels = c("0", "1"))
data$patient_pop___5.factor <- factor(data$patient_pop___5, levels = c("0", "1"))
data$patient_pop___6.factor <- factor(data$patient_pop___6, levels = c("0", "1"))
data$patient_pop___7.factor <- factor(data$patient_pop___7, levels = c("0", "1"))
data$patient_pop___8.factor <- factor(data$patient_pop___8, levels = c("0", "1"))
data$patient_pop___9.factor <- factor(data$patient_pop___9, levels = c("0", "1"))
data$patient_pop___10.factor <- factor(data$patient_pop___10, levels = c("0", "1"))
data$patient_pop___11.factor <- factor(data$patient_pop___11, levels = c("0", "1"))
data$patient_pop___12.factor <- factor(data$patient_pop___12, levels = c("0", "1"))
data$patient_pop___13.factor <- factor(data$patient_pop___13, levels = c("0", "1"))
data$patient_pop___15.factor <- factor(data$patient_pop___15, levels = c("0", "1"))
data$patient_pop___14.factor <- factor(data$patient_pop___14, levels = c("0", "1"))
data$patient_pop_cog___1.factor <- factor(data$patient_pop_cog___1, levels = c("0", "1"))
data$patient_pop_cog___2.factor <- factor(data$patient_pop_cog___2, levels = c("0", "1"))
data$patient_pop_cog___3.factor <- factor(data$patient_pop_cog___3, levels = c("0", "1"))
data$patient_pop_cog___4.factor <- factor(data$patient_pop_cog___4, levels = c("0", "1"))
data$patient_pop_motor___1.factor <- factor(data$patient_pop_motor___1, levels = c("0", "1"))
data$patient_pop_motor___2.factor <- factor(data$patient_pop_motor___2, levels = c("0", "1"))
data$patient_pop_motor___3.factor <- factor(data$patient_pop_motor___3, levels = c("0", "1"))
data$patient_pop_motor___4.factor <- factor(data$patient_pop_motor___4, levels = c("0", "1"))
data$patient_pop_age___1.factor <- factor(data$patient_pop_age___1, levels = c("0", "1"))
data$patient_pop_age___2.factor <- factor(data$patient_pop_age___2, levels = c("0", "1"))
data$patient_pop_age___3.factor <- factor(data$patient_pop_age___3, levels = c("0", "1"))
data$patient_pop_age___4.factor <- factor(data$patient_pop_age___4, levels = c("0", "1"))
data$patient_pop_age___5.factor <- factor(data$patient_pop_age___5, levels = c("0", "1"))
data$patient_pop_age___6.factor <- factor(data$patient_pop_age___6, levels = c("0", "1"))
data$patient_pop_age___7.factor <- factor(data$patient_pop_age___7, levels = c("0", "1"))
data$patient_pop_age___8.factor <- factor(data$patient_pop_age___8, levels = c("0", "1"))
data$patient_pop_age___9.factor <- factor(data$patient_pop_age___9, levels = c("0", "1"))
data$patient_pop_phase___1.factor <- factor(data$patient_pop_phase___1, levels = c("0", "1"))
data$patient_pop_phase___2.factor <- factor(data$patient_pop_phase___2, levels = c("0", "1"))
data$patient_pop_phase___3.factor <- factor(data$patient_pop_phase___3, levels = c("0", "1"))
data$past_telepresence___1.factor <- factor(data$past_telepresence___1, levels = c("0", "1"))
data$past_telepresence___2.factor <- factor(data$past_telepresence___2, levels = c("0", "1"))
data$past_telepresence___3.factor <- factor(data$past_telepresence___3, levels = c("0", "1"))
data$past_telepresence___4.factor <- factor(data$past_telepresence___4, levels = c("0", "1"))
data$past_telepresence___5.factor <- factor(data$past_telepresence___5, levels = c("0", "1"))
data$past_telepresence___6.factor <- factor(data$past_telepresence___6, levels = c("0", "1"))
data$covid_inperson.factor <- factor(data$covid_inperson, levels = c("1", "0"))
data$covid_telepresence___1.factor <- factor(data$covid_telepresence___1, levels = c("0", "1"))
data$covid_telepresence___2.factor <- factor(data$covid_telepresence___2, levels = c("0", "1"))
data$covid_telepresence___3.factor <- factor(data$covid_telepresence___3, levels = c("0", "1"))
data$covid_telepresence___4.factor <- factor(data$covid_telepresence___4, levels = c("0", "1"))
data$covid_telepresence___5.factor <- factor(data$covid_telepresence___5, levels = c("0", "1"))
data$covid_telepresence___6.factor <- factor(data$covid_telepresence___6, levels = c("0", "1"))
data$video_plan.factor <- factor(data$video_plan, levels = c("1", "0"))
data$video_challenges___1.factor <- factor(data$video_challenges___1, levels = c("0", "1"))
data$video_challenges___2.factor <- factor(data$video_challenges___2, levels = c("0", "1"))
data$video_challenges___3.factor <- factor(data$video_challenges___3, levels = c("0", "1"))
data$video_challenges___4.factor <- factor(data$video_challenges___4, levels = c("0", "1"))
data$video_challenges___5.factor <- factor(data$video_challenges___5, levels = c("0", "1"))
data$video_challenges___6.factor <- factor(data$video_challenges___6, levels = c("0", "1"))
data$video_challenges___7.factor <- factor(data$video_challenges___7, levels = c("0", "1"))
data$video_challenges___8.factor <- factor(data$video_challenges___8, levels = c("0", "1"))
data$video_challenges___9.factor <- factor(data$video_challenges___9, levels = c("0", "1"))
data$video_challenges___10.factor <- factor(data$video_challenges___10, levels = c("0", "1"))
data$video_challenges___11.factor <- factor(data$video_challenges___11, levels = c("0", "1"))
data$video_challenges___12.factor <- factor(data$video_challenges___12, levels = c("0", "1"))
data$pvideo_challenges___1.factor <- factor(data$pvideo_challenges___1, levels = c("0", "1"))
data$pvideo_challenges___2.factor <- factor(data$pvideo_challenges___2, levels = c("0", "1"))
data$pvideo_challenges___3.factor <- factor(data$pvideo_challenges___3, levels = c("0", "1"))
data$pvideo_challenges___4.factor <- factor(data$pvideo_challenges___4, levels = c("0", "1"))
data$pvideo_challenges___5.factor <- factor(data$pvideo_challenges___5, levels = c("0", "1"))
data$pvideo_challenges___6.factor <- factor(data$pvideo_challenges___6, levels = c("0", "1"))
data$pvideo_challenges___7.factor <- factor(data$pvideo_challenges___7, levels = c("0", "1"))
data$pvideo_challenges___8.factor <- factor(data$pvideo_challenges___8, levels = c("0", "1"))
data$pvideo_challenges___9.factor <- factor(data$pvideo_challenges___9, levels = c("0", "1"))
data$pvideo_challenges___10.factor <- factor(data$pvideo_challenges___10, levels = c("0", "1"))
data$pvideo_challenges___11.factor <- factor(data$pvideo_challenges___11, levels = c("0", "1"))
data$pvideo_challenges___12.factor <- factor(data$pvideo_challenges___12, levels = c("0", "1"))
data$video_future.factor <- factor(data$video_future, levels = c("1", "2", "3"))
data$flo_prior_experience___1.factor <- factor(data$flo_prior_experience___1, levels = c("0", "1"))
data$flo_prior_experience___2.factor <- factor(data$flo_prior_experience___2, levels = c("0", "1"))
data$flo_prior_experience___3.factor <- factor(data$flo_prior_experience___3, levels = c("0", "1"))
data$flo_prior_experience___4.factor <- factor(data$flo_prior_experience___4, levels = c("0", "1"))
data$flo_prior_experience___5.factor <- factor(data$flo_prior_experience___5, levels = c("0", "1"))
data$flo_location___1.factor <- factor(data$flo_location___1, levels = c("0", "1"))
data$flo_location___2.factor <- factor(data$flo_location___2, levels = c("0", "1"))
data$flo_location___3.factor <- factor(data$flo_location___3, levels = c("0", "1"))
data$flo_location___4.factor <- factor(data$flo_location___4, levels = c("0", "1"))
data$flo_location___5.factor <- factor(data$flo_location___5, levels = c("0", "1"))
data$flo_location___6.factor <- factor(data$flo_location___6, levels = c("0", "1"))
data$flo_location___7.factor <- factor(data$flo_location___7, levels = c("0", "1"))
data$flo_location___8.factor <- factor(data$flo_location___8, levels = c("0", "1"))
data$flo_location___9.factor <- factor(data$flo_location___9, levels = c("0", "1"))
data$flo_location___10.factor <- factor(data$flo_location___10, levels = c("0", "1"))
data$telemed_sys_feat_mobile.factor <- factor(data$telemed_sys_feat_mobile, levels = c("1", "2", "3", "4", "5"))
data$telemed_sys_feat_autodrive.factor <- factor(data$telemed_sys_feat_autodrive, levels = c("1", "2", "3", "4", "5"))
data$telemed_sys_feat_arms.factor <- factor(data$telemed_sys_feat_arms, levels = c("1", "2", "3", "4", "5"))
data$telemed_sys_feat_face.factor <- factor(data$telemed_sys_feat_face, levels = c("1", "2", "3", "4", "5"))
data$telemed_sys_feat_autointer.factor <- factor(data$telemed_sys_feat_autointer, levels = c("1", "2", "3", "4", "5"))
data$telemed_sys_feat_web.factor <- factor(data$telemed_sys_feat_web, levels = c("1", "2", "3", "4", "5"))
data$telemed_sys_feat_games.factor <- factor(data$telemed_sys_feat_games, levels = c("1", "2", "3", "4", "5"))
data$telemed_sys_feat_asses.factor <- factor(data$telemed_sys_feat_asses, levels = c("1", "2", "3", "4", "5"))
data$telemed_sys_feat_screen.factor <- factor(data$telemed_sys_feat_screen, levels = c("1", "2", "3", "4", "5"))
data$telemed_sys_feat_vid.factor <- factor(data$telemed_sys_feat_vid, levels = c("1", "2", "3", "4", "5"))
data$activities_d_motor_ass___1.factor <- factor(data$activities_d_motor_ass___1, levels = c("0", "1"))
data$activities_d_motor_ass___3.factor <- factor(data$activities_d_motor_ass___3, levels = c("0", "1"))
data$activities_d_motor_ass___4.factor <- factor(data$activities_d_motor_ass___4, levels = c("0", "1"))
data$activities_d_motor_ass___5.factor <- factor(data$activities_d_motor_ass___5, levels = c("0", "1"))
data$activities_d_motor_ass___6.factor <- factor(data$activities_d_motor_ass___6, levels = c("0", "1"))
data$activities_d_stretching___1.factor <- factor(data$activities_d_stretching___1, levels = c("0", "1"))
data$activities_d_stretching___3.factor <- factor(data$activities_d_stretching___3, levels = c("0", "1"))
data$activities_d_stretching___4.factor <- factor(data$activities_d_stretching___4, levels = c("0", "1"))
data$activities_d_stretching___5.factor <- factor(data$activities_d_stretching___5, levels = c("0", "1"))
data$activities_d_stretching___6.factor <- factor(data$activities_d_stretching___6, levels = c("0", "1"))
data$activities_d_strength___1.factor <- factor(data$activities_d_strength___1, levels = c("0", "1"))
data$activities_d_strength___3.factor <- factor(data$activities_d_strength___3, levels = c("0", "1"))
data$activities_d_strength___4.factor <- factor(data$activities_d_strength___4, levels = c("0", "1"))
data$activities_d_strength___5.factor <- factor(data$activities_d_strength___5, levels = c("0", "1"))
data$activities_d_strength___6.factor <- factor(data$activities_d_strength___6, levels = c("0", "1"))
data$activities_d_adl___1.factor <- factor(data$activities_d_adl___1, levels = c("0", "1"))
data$activities_d_adl___3.factor <- factor(data$activities_d_adl___3, levels = c("0", "1"))
data$activities_d_adl___4.factor <- factor(data$activities_d_adl___4, levels = c("0", "1"))
data$activities_d_adl___5.factor <- factor(data$activities_d_adl___5, levels = c("0", "1"))
data$activities_d_adl___6.factor <- factor(data$activities_d_adl___6, levels = c("0", "1"))
data$activities_d_cog_ass___1.factor <- factor(data$activities_d_cog_ass___1, levels = c("0", "1"))
data$activities_d_cog_ass___3.factor <- factor(data$activities_d_cog_ass___3, levels = c("0", "1"))
data$activities_d_cog_ass___4.factor <- factor(data$activities_d_cog_ass___4, levels = c("0", "1"))
data$activities_d_cog_ass___5.factor <- factor(data$activities_d_cog_ass___5, levels = c("0", "1"))
data$activities_d_cog_ass___6.factor <- factor(data$activities_d_cog_ass___6, levels = c("0", "1"))
data$activites_d_cog_ex___1.factor <- factor(data$activites_d_cog_ex___1, levels = c("0", "1"))
data$activites_d_cog_ex___3.factor <- factor(data$activites_d_cog_ex___3, levels = c("0", "1"))
data$activites_d_cog_ex___4.factor <- factor(data$activites_d_cog_ex___4, levels = c("0", "1"))
data$activites_d_cog_ex___5.factor <- factor(data$activites_d_cog_ex___5, levels = c("0", "1"))
data$activites_d_cog_ex___6.factor <- factor(data$activites_d_cog_ex___6, levels = c("0", "1"))
data$activities_d_env_adap___1.factor <- factor(data$activities_d_env_adap___1, levels = c("0", "1"))
data$activities_d_env_adap___3.factor <- factor(data$activities_d_env_adap___3, levels = c("0", "1"))
data$activities_d_env_adap___4.factor <- factor(data$activities_d_env_adap___4, levels = c("0", "1"))
data$activities_d_env_adap___5.factor <- factor(data$activities_d_env_adap___5, levels = c("0", "1"))
data$activities_d_env_adap___6.factor <- factor(data$activities_d_env_adap___6, levels = c("0", "1"))
data$activities_d_orthotics___1.factor <- factor(data$activities_d_orthotics___1, levels = c("0", "1"))
data$activities_d_orthotics___3.factor <- factor(data$activities_d_orthotics___3, levels = c("0", "1"))
data$activities_d_orthotics___4.factor <- factor(data$activities_d_orthotics___4, levels = c("0", "1"))
data$activities_d_orthotics___5.factor <- factor(data$activities_d_orthotics___5, levels = c("0", "1"))
data$activities_d_orthotics___6.factor <- factor(data$activities_d_orthotics___6, levels = c("0", "1"))
data$activities_d_surg___1.factor <- factor(data$activities_d_surg___1, levels = c("0", "1"))
data$activities_d_surg___3.factor <- factor(data$activities_d_surg___3, levels = c("0", "1"))
data$activities_d_surg___4.factor <- factor(data$activities_d_surg___4, levels = c("0", "1"))
data$activities_d_surg___5.factor <- factor(data$activities_d_surg___5, levels = c("0", "1"))
data$activities_d_surg___6.factor <- factor(data$activities_d_surg___6, levels = c("0", "1"))
data$activities_d_rads___1.factor <- factor(data$activities_d_rads___1, levels = c("0", "1"))
data$activities_d_rads___3.factor <- factor(data$activities_d_rads___3, levels = c("0", "1"))
data$activities_d_rads___4.factor <- factor(data$activities_d_rads___4, levels = c("0", "1"))
data$activities_d_rads___5.factor <- factor(data$activities_d_rads___5, levels = c("0", "1"))
data$activities_d_rads___6.factor <- factor(data$activities_d_rads___6, levels = c("0", "1"))
data$activities_d_med___1.factor <- factor(data$activities_d_med___1, levels = c("0", "1"))
data$activities_d_med___3.factor <- factor(data$activities_d_med___3, levels = c("0", "1"))
data$activities_d_med___4.factor <- factor(data$activities_d_med___4, levels = c("0", "1"))
data$activities_d_med___5.factor <- factor(data$activities_d_med___5, levels = c("0", "1"))
data$activities_d_med___6.factor <- factor(data$activities_d_med___6, levels = c("0", "1"))
data$activities_b_motor_ass___1.factor <- factor(data$activities_b_motor_ass___1, levels = c("0", "1"))
data$activities_b_motor_ass___3.factor <- factor(data$activities_b_motor_ass___3, levels = c("0", "1"))
data$activities_b_motor_ass___4.factor <- factor(data$activities_b_motor_ass___4, levels = c("0", "1"))
data$activities_b_motor_ass___5.factor <- factor(data$activities_b_motor_ass___5, levels = c("0", "1"))
data$activities_b_stretching___1.factor <- factor(data$activities_b_stretching___1, levels = c("0", "1"))
data$activities_b_stretching___3.factor <- factor(data$activities_b_stretching___3, levels = c("0", "1"))
data$activities_b_stretching___4.factor <- factor(data$activities_b_stretching___4, levels = c("0", "1"))
data$activities_b_stretching___5.factor <- factor(data$activities_b_stretching___5, levels = c("0", "1"))
data$activities_b_strength___1.factor <- factor(data$activities_b_strength___1, levels = c("0", "1"))
data$activities_b_strength___3.factor <- factor(data$activities_b_strength___3, levels = c("0", "1"))
data$activities_b_strength___4.factor <- factor(data$activities_b_strength___4, levels = c("0", "1"))
data$activities_b_strength___5.factor <- factor(data$activities_b_strength___5, levels = c("0", "1"))
data$activities_b_adl___1.factor <- factor(data$activities_b_adl___1, levels = c("0", "1"))
data$activities_b_adl___3.factor <- factor(data$activities_b_adl___3, levels = c("0", "1"))
data$activities_b_adl___4.factor <- factor(data$activities_b_adl___4, levels = c("0", "1"))
data$activities_b_adl___5.factor <- factor(data$activities_b_adl___5, levels = c("0", "1"))
data$activities_b_cog_ass___1.factor <- factor(data$activities_b_cog_ass___1, levels = c("0", "1"))
data$activities_b_cog_ass___3.factor <- factor(data$activities_b_cog_ass___3, levels = c("0", "1"))
data$activities_b_cog_ass___4.factor <- factor(data$activities_b_cog_ass___4, levels = c("0", "1"))
data$activities_b_cog_ass___5.factor <- factor(data$activities_b_cog_ass___5, levels = c("0", "1"))
data$activites_b_cog_ex___1.factor <- factor(data$activites_b_cog_ex___1, levels = c("0", "1"))
data$activites_b_cog_ex___3.factor <- factor(data$activites_b_cog_ex___3, levels = c("0", "1"))
data$activites_b_cog_ex___4.factor <- factor(data$activites_b_cog_ex___4, levels = c("0", "1"))
data$activites_b_cog_ex___5.factor <- factor(data$activites_b_cog_ex___5, levels = c("0", "1"))
data$activities_b_env_adap___1.factor <- factor(data$activities_b_env_adap___1, levels = c("0", "1"))
data$activities_b_env_adap___3.factor <- factor(data$activities_b_env_adap___3, levels = c("0", "1"))
data$activities_b_env_adap___4.factor <- factor(data$activities_b_env_adap___4, levels = c("0", "1"))
data$activities_b_env_adap___5.factor <- factor(data$activities_b_env_adap___5, levels = c("0", "1"))
data$activities_b_orthotics___1.factor <- factor(data$activities_b_orthotics___1, levels = c("0", "1"))
data$activities_b_orthotics___3.factor <- factor(data$activities_b_orthotics___3, levels = c("0", "1"))
data$activities_b_orthotics___4.factor <- factor(data$activities_b_orthotics___4, levels = c("0", "1"))
data$activities_b_orthotics___5.factor <- factor(data$activities_b_orthotics___5, levels = c("0", "1"))
data$activities_b_surg___1.factor <- factor(data$activities_b_surg___1, levels = c("0", "1"))
data$activities_b_surg___3.factor <- factor(data$activities_b_surg___3, levels = c("0", "1"))
data$activities_b_surg___4.factor <- factor(data$activities_b_surg___4, levels = c("0", "1"))
data$activities_b_surg___5.factor <- factor(data$activities_b_surg___5, levels = c("0", "1"))
data$activities_b_rads___1.factor <- factor(data$activities_b_rads___1, levels = c("0", "1"))
data$activities_b_rads___3.factor <- factor(data$activities_b_rads___3, levels = c("0", "1"))
data$activities_b_rads___4.factor <- factor(data$activities_b_rads___4, levels = c("0", "1"))
data$activities_b_rads___5.factor <- factor(data$activities_b_rads___5, levels = c("0", "1"))
data$activities_b_med___1.factor <- factor(data$activities_b_med___1, levels = c("0", "1"))
data$activities_b_med___3.factor <- factor(data$activities_b_med___3, levels = c("0", "1"))
data$activities_b_med___4.factor <- factor(data$activities_b_med___4, levels = c("0", "1"))
data$activities_b_med___5.factor <- factor(data$activities_b_med___5, levels = c("0", "1"))
data$robot_feelings_positive.factor <- factor(data$robot_feelings_positive, levels = c("1", "2", "3", "4", "5"))
data$robot_feelings_same.factor <- factor(data$robot_feelings_same, levels = c("1", "2", "3", "4", "5"))
data$robot_feelings_indep.factor <- factor(data$robot_feelings_indep, levels = c("1", "2", "3", "4", "5"))
data$robot_feelings_focus.factor <- factor(data$robot_feelings_focus, levels = c("1", "2", "3", "4", "5"))
data$robot_feelings_accurate.factor <- factor(data$robot_feelings_accurate, levels = c("1", "2", "3", "4", "5"))
data$robot_feelings_expensive.factor <- factor(data$robot_feelings_expensive, levels = c("1", "2", "3", "4", "5"))
data$robot_feelings_time.factor <- factor(data$robot_feelings_time, levels = c("1", "2", "3", "4", "5"))
data$robot_feelings_unethical.factor <- factor(data$robot_feelings_unethical, levels = c("1", "2", "3", "4", "5"))
data$robot_feelings_patwant.factor <- factor(data$robot_feelings_patwant, levels = c("1", "2", "3", "4", "5"))
data$robot_feelings_control.factor <- factor(data$robot_feelings_control, levels = c("1", "2", "3", "4", "5"))
data$robot_feelings_jobs.factor <- factor(data$robot_feelings_jobs, levels = c("1", "2", "3", "4", "5"))
data$robot_feelings_fail.factor <- factor(data$robot_feelings_fail, levels = c("1", "2", "3", "4", "5"))
data$robot_feelings_clinfeel.factor <- factor(data$robot_feelings_clinfeel, levels = c("1", "2", "3", "4", "5"))
data$robot_feelings_neg.factor <- factor(data$robot_feelings_neg, levels = c("1", "2", "3", "4", "5"))
data$robot_feelings_failethic.factor <- factor(data$robot_feelings_failethic, levels = c("1", "2", "3", "4", "5"))
data$robot_feelings_interact.factor <- factor(data$robot_feelings_interact, levels = c("1", "2", "3", "4", "5"))
data$robot_feelings_laws.factor <- factor(data$robot_feelings_laws, levels = c("1", "2", "3", "4", "5"))
data$robot_feelings_study.factor <- factor(data$robot_feelings_study, levels = c("1", "2", "3", "4", "5"))
data$robot_feelings_autonomy.factor <- factor(data$robot_feelings_autonomy, levels = c("1", "2", "3", "4", "5"))
data$robot_feelings_bad.factor <- factor(data$robot_feelings_bad, levels = c("1", "2", "3", "4", "5"))
data$giftcard.factor <- factor(data$giftcard, levels = c("1", "0"))
data$results.factor <- factor(data$results, levels = c("1", "0"))
data$clinician_survey_complete.factor <- factor(data$clinician_survey_complete, levels = c("0", "1", "2"))

levels(data$occupation.factor) <- c("Therapist", "Nurse", "Doctor", "Medical Technologist", "Other")
levels(data$gender.factor) <- c("Male", "Female", "Other")
levels(data$location___1.factor) <- c("Unchecked", "Checked")
levels(data$location___2.factor) <- c("Unchecked", "Checked")
levels(data$location___3.factor) <- c("Unchecked", "Checked")
levels(data$location___4.factor) <- c("Unchecked", "Checked")
levels(data$location___5.factor) <- c("Unchecked", "Checked")
levels(data$location___6.factor) <- c("Unchecked", "Checked")
levels(data$location___7.factor) <- c("Unchecked", "Checked")
levels(data$location___8.factor) <- c("Unchecked", "Checked")
levels(data$location___9.factor) <- c("Unchecked", "Checked")
levels(data$location___10.factor) <- c("Unchecked", "Checked")
levels(data$location___11.factor) <- c("Unchecked", "Checked")
levels(data$location___12.factor) <- c("Unchecked", "Checked")
levels(data$location_type___1.factor) <- c("Unchecked", "Checked")
levels(data$location_type___2.factor) <- c("Unchecked", "Checked")
levels(data$location_type___3.factor) <- c("Unchecked", "Checked")
levels(data$location_country.factor) <- c("United States of America", "Canada", "Mexico", "Afghanistan", "Albania", "Algeria", "Andorra", "Angola", "Antigua and Barbuda", "Argentina", "Armenia", "Australia", "Austria", "Azerbaijan", "Bahamas", "Bahrain", "Bangladesh", "Barbados", "Belarus", "Belgium", "Belize", "Benin", "Bhutan", "Bolivia (Plurinational State of)", "Bosnia and Herzegovina", "Botswana", "Brazil", "Brunei Darussalam", "Bulgaria", "Burkina Faso", "Burundi", "Cabo Verde", "Cambodia", "Cameroon", "Central African Republic", "Chad", "Chile", "China", "Colombia", "Comoros", "Congo", "Costa Rica", "Côte DIvoire", "Croatia", "Cuba", "Cyprus", "Czech Republic", "Democratic Peoples Republic of Korea", "Democratic Republic of the Congo", "Denmark", "Djibouti", "Dominica", "Dominican Republic", "Ecuador", "Egypt", "El Salvador", "Equatorial Guinea", "Eritrea", "Estonia", "Eswatini (the Kingdom of)", "Ethiopia", "Fiji", "Finland", "France", "Gabon", "Gambia (Republic of The)", "Georgia", "Germany", "Ghana", "Greece", "Grenada", "Guatemala", "Guinea", "Guinea Bissau", "Guyana", "Haiti", "Honduras", "Hungary", "Iceland", "India", "Indonesia", "Iran (Islamic Republic of)", "Iraq", "Ireland", "Israel", "Italy", "Jamaica", "Japan", "Jordan", "Kazakhstan", "Kenya", "Kiribati", "Kuwait", "Kyrgyzstan", "Lao Peoples Democratic Republic", "Latvia", "Lebanon", "Lesotho", "Liberia", "Libya", "Liechtenstein", "Lithuania", "Luxembourg", "Madagascar", "Malawi", "Malaysia", "Maldives", "Mali", "Malta", "Marshall Islands", "Mauritania", "Mauritius", "Micronesia (Federated States of)", "Monaco", "Mongolia", "Montenegro", "Morocco", "Mozambique", "Myanmar", "Namibia", "Nauru", "Nepal", "Netherlands", "New Zealand", "Nicaragua", "Niger", "Nigeria", "Norway", "Oman", "Pakistan", "Palau", "Panama", "Papua New Guinea", "Paraguay", "Peru", "Philippines", "Poland", "Portugal", "Qatar", "Republic of Korea", "Republic of Moldova", "Romania", "Russian Federation", "Rwanda", "Saint Kitts and Nevis", "Saint Lucia", "Saint Vincent and the Grenadines", "Samoa", "San Marino", "Sao Tome and Principe", "Saudi Arabia", "Senegal", "Serbia", "Seychelles", "Sierra Leone", "Singapore", "Slovakia", "Slovenia", "Solomon Islands", "Somalia", "South Africa", "South Sudan", "Spain", "Sri Lanka", "Sudan", "Suriname", "Sweden", "Switzerland", "Syrian Arab Republic", "Tajikistan", "Thailand", "The former Yugoslav Republic of Macedonia", "Timor-Leste", "Togo", "Tonga", "Trinidad and Tobago", "Tunisia", "Turkey", "Turkmenistan", "Tuvalu", "Uganda", "Ukraine", "United Arab Emirates", "United Kingdom of Great Britain and Northern Ireland", "United Republic of Tanzania", "Uruguay", "Uzbekistan", "Vanuatu", "Bolivarian Republic of", "Viet Nam", "Yemen", "Zambia", "Zimbabwe")
levels(data$location_state.factor) <- c("Alabama", "Alaska", "Arizona", "Arkansas", "California", "Colorado", "Connecticut", "Delaware", "Florida", "Georgia", "Hawaii", "Idaho", "Illinois", "Indiana", "Iowa", "Kansas", "Kentucky", "Louisiana", "Maine", "Maryland", "Massachusetts", "Michigan", "Minnesota", "Mississippi", "Missouri", "Montana", "Nebraska", "Nevada", "New Hampshire", "New Jersey", "New Mexico", "New York", "North Carolina", "North Dakota", "Ohio", "Oklahoma", "Oregon", "Pennsylvania", "Rhode Island", "South Carolina", "South Dakota", "Tennessee", "Texas", "Utah", "Vermont", "Virginia", "Washington", "West Virginia", "Wisconsin", "Wyoming", "Washington, D.C.")
levels(data$patient_pop___1.factor) <- c("Unchecked", "Checked")
levels(data$patient_pop___2.factor) <- c("Unchecked", "Checked")
levels(data$patient_pop___3.factor) <- c("Unchecked", "Checked")
levels(data$patient_pop___4.factor) <- c("Unchecked", "Checked")
levels(data$patient_pop___5.factor) <- c("Unchecked", "Checked")
levels(data$patient_pop___6.factor) <- c("Unchecked", "Checked")
levels(data$patient_pop___7.factor) <- c("Unchecked", "Checked")
levels(data$patient_pop___8.factor) <- c("Unchecked", "Checked")
levels(data$patient_pop___9.factor) <- c("Unchecked", "Checked")
levels(data$patient_pop___10.factor) <- c("Unchecked", "Checked")
levels(data$patient_pop___11.factor) <- c("Unchecked", "Checked")
levels(data$patient_pop___12.factor) <- c("Unchecked", "Checked")
levels(data$patient_pop___13.factor) <- c("Unchecked", "Checked")
levels(data$patient_pop___15.factor) <- c("Unchecked", "Checked")
levels(data$patient_pop___14.factor) <- c("Unchecked", "Checked")
levels(data$patient_pop_cog___1.factor) <- c("Unchecked", "Checked")
levels(data$patient_pop_cog___2.factor) <- c("Unchecked", "Checked")
levels(data$patient_pop_cog___3.factor) <- c("Unchecked", "Checked")
levels(data$patient_pop_cog___4.factor) <- c("Unchecked", "Checked")
levels(data$patient_pop_motor___1.factor) <- c("Unchecked", "Checked")
levels(data$patient_pop_motor___2.factor) <- c("Unchecked", "Checked")
levels(data$patient_pop_motor___3.factor) <- c("Unchecked", "Checked")
levels(data$patient_pop_motor___4.factor) <- c("Unchecked", "Checked")
levels(data$patient_pop_age___1.factor) <- c("Unchecked", "Checked")
levels(data$patient_pop_age___2.factor) <- c("Unchecked", "Checked")
levels(data$patient_pop_age___3.factor) <- c("Unchecked", "Checked")
levels(data$patient_pop_age___4.factor) <- c("Unchecked", "Checked")
levels(data$patient_pop_age___5.factor) <- c("Unchecked", "Checked")
levels(data$patient_pop_age___6.factor) <- c("Unchecked", "Checked")
levels(data$patient_pop_age___7.factor) <- c("Unchecked", "Checked")
levels(data$patient_pop_age___8.factor) <- c("Unchecked", "Checked")
levels(data$patient_pop_age___9.factor) <- c("Unchecked", "Checked")
levels(data$patient_pop_phase___1.factor) <- c("Unchecked", "Checked")
levels(data$patient_pop_phase___2.factor) <- c("Unchecked", "Checked")
levels(data$patient_pop_phase___3.factor) <- c("Unchecked", "Checked")
levels(data$past_telepresence___1.factor) <- c("Unchecked", "Checked")
levels(data$past_telepresence___2.factor) <- c("Unchecked", "Checked")
levels(data$past_telepresence___3.factor) <- c("Unchecked", "Checked")
levels(data$past_telepresence___4.factor) <- c("Unchecked", "Checked")
levels(data$past_telepresence___5.factor) <- c("Unchecked", "Checked")
levels(data$past_telepresence___6.factor) <- c("Unchecked", "Checked")
levels(data$covid_inperson.factor) <- c("Yes", "No")
levels(data$covid_telepresence___1.factor) <- c("Unchecked", "Checked")
levels(data$covid_telepresence___2.factor) <- c("Unchecked", "Checked")
levels(data$covid_telepresence___3.factor) <- c("Unchecked", "Checked")
levels(data$covid_telepresence___4.factor) <- c("Unchecked", "Checked")
levels(data$covid_telepresence___5.factor) <- c("Unchecked", "Checked")
levels(data$covid_telepresence___6.factor) <- c("Unchecked", "Checked")
levels(data$video_plan.factor) <- c("Yes", "No")
levels(data$video_challenges___1.factor) <- c("Unchecked", "Checked")
levels(data$video_challenges___2.factor) <- c("Unchecked", "Checked")
levels(data$video_challenges___3.factor) <- c("Unchecked", "Checked")
levels(data$video_challenges___4.factor) <- c("Unchecked", "Checked")
levels(data$video_challenges___5.factor) <- c("Unchecked", "Checked")
levels(data$video_challenges___6.factor) <- c("Unchecked", "Checked")
levels(data$video_challenges___7.factor) <- c("Unchecked", "Checked")
levels(data$video_challenges___8.factor) <- c("Unchecked", "Checked")
levels(data$video_challenges___9.factor) <- c("Unchecked", "Checked")
levels(data$video_challenges___10.factor) <- c("Unchecked", "Checked")
levels(data$video_challenges___11.factor) <- c("Unchecked", "Checked")
levels(data$video_challenges___12.factor) <- c("Unchecked", "Checked")
levels(data$pvideo_challenges___1.factor) <- c("Unchecked", "Checked")
levels(data$pvideo_challenges___2.factor) <- c("Unchecked", "Checked")
levels(data$pvideo_challenges___3.factor) <- c("Unchecked", "Checked")
levels(data$pvideo_challenges___4.factor) <- c("Unchecked", "Checked")
levels(data$pvideo_challenges___5.factor) <- c("Unchecked", "Checked")
levels(data$pvideo_challenges___6.factor) <- c("Unchecked", "Checked")
levels(data$pvideo_challenges___7.factor) <- c("Unchecked", "Checked")
levels(data$pvideo_challenges___8.factor) <- c("Unchecked", "Checked")
levels(data$pvideo_challenges___9.factor) <- c("Unchecked", "Checked")
levels(data$pvideo_challenges___10.factor) <- c("Unchecked", "Checked")
levels(data$pvideo_challenges___11.factor) <- c("Unchecked", "Checked")
levels(data$pvideo_challenges___12.factor) <- c("Unchecked", "Checked")
levels(data$video_future.factor) <- c("Yes", "No", "Not Sure")
levels(data$flo_prior_experience___1.factor) <- c("Unchecked", "Checked")
levels(data$flo_prior_experience___2.factor) <- c("Unchecked", "Checked")
levels(data$flo_prior_experience___3.factor) <- c("Unchecked", "Checked")
levels(data$flo_prior_experience___4.factor) <- c("Unchecked", "Checked")
levels(data$flo_prior_experience___5.factor) <- c("Unchecked", "Checked")
levels(data$flo_location___1.factor) <- c("Unchecked", "Checked")
levels(data$flo_location___2.factor) <- c("Unchecked", "Checked")
levels(data$flo_location___3.factor) <- c("Unchecked", "Checked")
levels(data$flo_location___4.factor) <- c("Unchecked", "Checked")
levels(data$flo_location___5.factor) <- c("Unchecked", "Checked")
levels(data$flo_location___6.factor) <- c("Unchecked", "Checked")
levels(data$flo_location___7.factor) <- c("Unchecked", "Checked")
levels(data$flo_location___8.factor) <- c("Unchecked", "Checked")
levels(data$flo_location___9.factor) <- c("Unchecked", "Checked")
levels(data$flo_location___10.factor) <- c("Unchecked", "Checked")
levels(data$telemed_sys_feat_mobile.factor) <- c("Extremely Useless", "Somewhat Useless", "Neutral", "Somewhat Useful", "Extremely Useful")
levels(data$telemed_sys_feat_autodrive.factor) <- c("Extremely Useless", "Somewhat Useless", "Neutral", "Somewhat Useful", "Extremely Useful")
levels(data$telemed_sys_feat_arms.factor) <- c("Extremely Useless", "Somewhat Useless", "Neutral", "Somewhat Useful", "Extremely Useful")
levels(data$telemed_sys_feat_face.factor) <- c("Extremely Useless", "Somewhat Useless", "Neutral", "Somewhat Useful", "Extremely Useful")
levels(data$telemed_sys_feat_autointer.factor) <- c("Extremely Useless", "Somewhat Useless", "Neutral", "Somewhat Useful", "Extremely Useful")
levels(data$telemed_sys_feat_web.factor) <- c("Extremely Useless", "Somewhat Useless", "Neutral", "Somewhat Useful", "Extremely Useful")
levels(data$telemed_sys_feat_games.factor) <- c("Extremely Useless", "Somewhat Useless", "Neutral", "Somewhat Useful", "Extremely Useful")
levels(data$telemed_sys_feat_asses.factor) <- c("Extremely Useless", "Somewhat Useless", "Neutral", "Somewhat Useful", "Extremely Useful")
levels(data$telemed_sys_feat_screen.factor) <- c("Extremely Useless", "Somewhat Useless", "Neutral", "Somewhat Useful", "Extremely Useful")
levels(data$telemed_sys_feat_vid.factor) <- c("Extremely Useless", "Somewhat Useless", "Neutral", "Somewhat Useful", "Extremely Useful")
levels(data$activities_d_motor_ass___1.factor) <- c("Unchecked", "Checked")
levels(data$activities_d_motor_ass___3.factor) <- c("Unchecked", "Checked")
levels(data$activities_d_motor_ass___4.factor) <- c("Unchecked", "Checked")
levels(data$activities_d_motor_ass___5.factor) <- c("Unchecked", "Checked")
levels(data$activities_d_motor_ass___6.factor) <- c("Unchecked", "Checked")
levels(data$activities_d_stretching___1.factor) <- c("Unchecked", "Checked")
levels(data$activities_d_stretching___3.factor) <- c("Unchecked", "Checked")
levels(data$activities_d_stretching___4.factor) <- c("Unchecked", "Checked")
levels(data$activities_d_stretching___5.factor) <- c("Unchecked", "Checked")
levels(data$activities_d_stretching___6.factor) <- c("Unchecked", "Checked")
levels(data$activities_d_strength___1.factor) <- c("Unchecked", "Checked")
levels(data$activities_d_strength___3.factor) <- c("Unchecked", "Checked")
levels(data$activities_d_strength___4.factor) <- c("Unchecked", "Checked")
levels(data$activities_d_strength___5.factor) <- c("Unchecked", "Checked")
levels(data$activities_d_strength___6.factor) <- c("Unchecked", "Checked")
levels(data$activities_d_adl___1.factor) <- c("Unchecked", "Checked")
levels(data$activities_d_adl___3.factor) <- c("Unchecked", "Checked")
levels(data$activities_d_adl___4.factor) <- c("Unchecked", "Checked")
levels(data$activities_d_adl___5.factor) <- c("Unchecked", "Checked")
levels(data$activities_d_adl___6.factor) <- c("Unchecked", "Checked")
levels(data$activities_d_cog_ass___1.factor) <- c("Unchecked", "Checked")
levels(data$activities_d_cog_ass___3.factor) <- c("Unchecked", "Checked")
levels(data$activities_d_cog_ass___4.factor) <- c("Unchecked", "Checked")
levels(data$activities_d_cog_ass___5.factor) <- c("Unchecked", "Checked")
levels(data$activities_d_cog_ass___6.factor) <- c("Unchecked", "Checked")
levels(data$activites_d_cog_ex___1.factor) <- c("Unchecked", "Checked")
levels(data$activites_d_cog_ex___3.factor) <- c("Unchecked", "Checked")
levels(data$activites_d_cog_ex___4.factor) <- c("Unchecked", "Checked")
levels(data$activites_d_cog_ex___5.factor) <- c("Unchecked", "Checked")
levels(data$activites_d_cog_ex___6.factor) <- c("Unchecked", "Checked")
levels(data$activities_d_env_adap___1.factor) <- c("Unchecked", "Checked")
levels(data$activities_d_env_adap___3.factor) <- c("Unchecked", "Checked")
levels(data$activities_d_env_adap___4.factor) <- c("Unchecked", "Checked")
levels(data$activities_d_env_adap___5.factor) <- c("Unchecked", "Checked")
levels(data$activities_d_env_adap___6.factor) <- c("Unchecked", "Checked")
levels(data$activities_d_orthotics___1.factor) <- c("Unchecked", "Checked")
levels(data$activities_d_orthotics___3.factor) <- c("Unchecked", "Checked")
levels(data$activities_d_orthotics___4.factor) <- c("Unchecked", "Checked")
levels(data$activities_d_orthotics___5.factor) <- c("Unchecked", "Checked")
levels(data$activities_d_orthotics___6.factor) <- c("Unchecked", "Checked")
levels(data$activities_d_surg___1.factor) <- c("Unchecked", "Checked")
levels(data$activities_d_surg___3.factor) <- c("Unchecked", "Checked")
levels(data$activities_d_surg___4.factor) <- c("Unchecked", "Checked")
levels(data$activities_d_surg___5.factor) <- c("Unchecked", "Checked")
levels(data$activities_d_surg___6.factor) <- c("Unchecked", "Checked")
levels(data$activities_d_rads___1.factor) <- c("Unchecked", "Checked")
levels(data$activities_d_rads___3.factor) <- c("Unchecked", "Checked")
levels(data$activities_d_rads___4.factor) <- c("Unchecked", "Checked")
levels(data$activities_d_rads___5.factor) <- c("Unchecked", "Checked")
levels(data$activities_d_rads___6.factor) <- c("Unchecked", "Checked")
levels(data$activities_d_med___1.factor) <- c("Unchecked", "Checked")
levels(data$activities_d_med___3.factor) <- c("Unchecked", "Checked")
levels(data$activities_d_med___4.factor) <- c("Unchecked", "Checked")
levels(data$activities_d_med___5.factor) <- c("Unchecked", "Checked")
levels(data$activities_d_med___6.factor) <- c("Unchecked", "Checked")
levels(data$activities_b_motor_ass___1.factor) <- c("Unchecked", "Checked")
levels(data$activities_b_motor_ass___3.factor) <- c("Unchecked", "Checked")
levels(data$activities_b_motor_ass___4.factor) <- c("Unchecked", "Checked")
levels(data$activities_b_motor_ass___5.factor) <- c("Unchecked", "Checked")
levels(data$activities_b_stretching___1.factor) <- c("Unchecked", "Checked")
levels(data$activities_b_stretching___3.factor) <- c("Unchecked", "Checked")
levels(data$activities_b_stretching___4.factor) <- c("Unchecked", "Checked")
levels(data$activities_b_stretching___5.factor) <- c("Unchecked", "Checked")
levels(data$activities_b_strength___1.factor) <- c("Unchecked", "Checked")
levels(data$activities_b_strength___3.factor) <- c("Unchecked", "Checked")
levels(data$activities_b_strength___4.factor) <- c("Unchecked", "Checked")
levels(data$activities_b_strength___5.factor) <- c("Unchecked", "Checked")
levels(data$activities_b_adl___1.factor) <- c("Unchecked", "Checked")
levels(data$activities_b_adl___3.factor) <- c("Unchecked", "Checked")
levels(data$activities_b_adl___4.factor) <- c("Unchecked", "Checked")
levels(data$activities_b_adl___5.factor) <- c("Unchecked", "Checked")
levels(data$activities_b_cog_ass___1.factor) <- c("Unchecked", "Checked")
levels(data$activities_b_cog_ass___3.factor) <- c("Unchecked", "Checked")
levels(data$activities_b_cog_ass___4.factor) <- c("Unchecked", "Checked")
levels(data$activities_b_cog_ass___5.factor) <- c("Unchecked", "Checked")
levels(data$activites_b_cog_ex___1.factor) <- c("Unchecked", "Checked")
levels(data$activites_b_cog_ex___3.factor) <- c("Unchecked", "Checked")
levels(data$activites_b_cog_ex___4.factor) <- c("Unchecked", "Checked")
levels(data$activites_b_cog_ex___5.factor) <- c("Unchecked", "Checked")
levels(data$activities_b_env_adap___1.factor) <- c("Unchecked", "Checked")
levels(data$activities_b_env_adap___3.factor) <- c("Unchecked", "Checked")
levels(data$activities_b_env_adap___4.factor) <- c("Unchecked", "Checked")
levels(data$activities_b_env_adap___5.factor) <- c("Unchecked", "Checked")
levels(data$activities_b_orthotics___1.factor) <- c("Unchecked", "Checked")
levels(data$activities_b_orthotics___3.factor) <- c("Unchecked", "Checked")
levels(data$activities_b_orthotics___4.factor) <- c("Unchecked", "Checked")
levels(data$activities_b_orthotics___5.factor) <- c("Unchecked", "Checked")
levels(data$activities_b_surg___1.factor) <- c("Unchecked", "Checked")
levels(data$activities_b_surg___3.factor) <- c("Unchecked", "Checked")
levels(data$activities_b_surg___4.factor) <- c("Unchecked", "Checked")
levels(data$activities_b_surg___5.factor) <- c("Unchecked", "Checked")
levels(data$activities_b_rads___1.factor) <- c("Unchecked", "Checked")
levels(data$activities_b_rads___3.factor) <- c("Unchecked", "Checked")
levels(data$activities_b_rads___4.factor) <- c("Unchecked", "Checked")
levels(data$activities_b_rads___5.factor) <- c("Unchecked", "Checked")
levels(data$activities_b_med___1.factor) <- c("Unchecked", "Checked")
levels(data$activities_b_med___3.factor) <- c("Unchecked", "Checked")
levels(data$activities_b_med___4.factor) <- c("Unchecked", "Checked")
levels(data$activities_b_med___5.factor) <- c("Unchecked", "Checked")
levels(data$robot_feelings_positive.factor) <- c("Strongly Disagree", "Disagree", "Neutral", "Agree", "Strongly Agree")
levels(data$robot_feelings_same.factor) <- c("Strongly Disagree", "Disagree", "Neutral", "Agree", "Strongly Agree")
levels(data$robot_feelings_indep.factor) <- c("Strongly Disagree", "Disagree", "Neutral", "Agree", "Strongly Agree")
levels(data$robot_feelings_focus.factor) <- c("Strongly Disagree", "Disagree", "Neutral", "Agree", "Strongly Agree")
levels(data$robot_feelings_accurate.factor) <- c("Strongly Disagree", "Disagree", "Neutral", "Agree", "Strongly Agree")
levels(data$robot_feelings_expensive.factor) <- c("Strongly Disagree", "Disagree", "Neutral", "Agree", "Strongly Agree")
levels(data$robot_feelings_time.factor) <- c("Strongly Disagree", "Disagree", "Neutral", "Agree", "Strongly Agree")
levels(data$robot_feelings_unethical.factor) <- c("Strongly Disagree", "Disagree", "Neutral", "Agree", "Strongly Agree")
levels(data$robot_feelings_patwant.factor) <- c("Strongly Disagree", "Disagree", "Neutral", "Agree", "Strongly Agree")
levels(data$robot_feelings_control.factor) <- c("Strongly Disagree", "Disagree", "Neutral", "Agree", "Strongly Agree")
levels(data$robot_feelings_jobs.factor) <- c("Strongly Disagree", "Disagree", "Neutral", "Agree", "Strongly Agree")
levels(data$robot_feelings_fail.factor) <- c("Strongly Disagree", "Disagree", "Neutral", "Agree", "Strongly Agree")
levels(data$robot_feelings_clinfeel.factor) <- c("Strongly Disagree", "Disagree", "Neutral", "Agree", "Strongly Agree")
levels(data$robot_feelings_neg.factor) <- c("Strongly Disagree", "Disagree", "Neutral", "Agree", "Strongly Agree")
levels(data$robot_feelings_failethic.factor) <- c("Strongly Disagree", "Disagree", "Neutral", "Agree", "Strongly Agree")
levels(data$robot_feelings_interact.factor) <- c("Strongly Disagree", "Disagree", "Neutral", "Agree", "Strongly Agree")
levels(data$robot_feelings_laws.factor) <- c("Strongly Disagree", "Disagree", "Neutral", "Agree", "Strongly Agree")
levels(data$robot_feelings_study.factor) <- c("Strongly Disagree", "Disagree", "Neutral", "Agree", "Strongly Agree")
levels(data$robot_feelings_autonomy.factor) <- c("Strongly Disagree", "Disagree", "Neutral", "Agree", "Strongly Agree")
levels(data$robot_feelings_bad.factor) <- c("Strongly Disagree", "Disagree", "Neutral", "Agree", "Strongly Agree")
levels(data$giftcard.factor) <- c("Yes", "No")
levels(data$results.factor) <- c("Yes", "No")
levels(data$clinician_survey_complete.factor) <- c("Incomplete", "Unverified", "Complete")

## Fix mispelling in some activities:
colnames(data)<-sub('activites','activities',colnames(data))


### Clean Data ###

# Note: Some cleaning has been done manually in the data loading script
#       by changing values for the factors

## Remove all of the non-complete data entries
data <-
  data[which(data$clinician_survey_complete.factor == 'Complete'),]

## Set completion column as date, strips away time
data$clinician_survey_timestamp <-
  as.Date(data$clinician_survey_timestamp)

### Filter Data ###
## Functions ##
filter_clinician_type <- function(data, type) {
  # Type can be therapist, doctor, nurse, medical technologist
  type <-
    sprintf('%s%s', toupper(substring(type, 1, 1)), tolower(substring(type, 2)))
  start_len <- nrow(data)
  toReturn<-data[which(data$occupation.factor == type),]
  cat(sprintf('Narrowing by clinician type: %s, yeilding %i samples from %i samples\n', type,nrow(toReturn),start_len))
  
  
  return(toReturn)
}

filter_country <- function(data, country) {
  # Type can be therapist, doctor, nurse, medical technologist
  start_len <- nrow(data)
  toReturn<-data[which(grepl(country, data$location_country.factor, ignore.case = TRUE)), ]
  cat(sprintf('Narrowing by country: %s, yeilding %i samples from %i samples\n', country,nrow(toReturn),start_len))
  return(toReturn)
}

filter_record_number <- function(data, record_number) {
  start_len <- nrow(data)
  toReturn<-data[which(data$record_id <= record_number),]
  cat(sprintf('Narrowing by record number: %i, yeilding %i samples from %i samples\n', record_number,nrow(toReturn),start_len))
  
  return(toReturn)
}

filter_years_experience <- function(data, years) {
  start_len <- nrow(data)
  toReturn<-data[which(data$years_experience >= years),]
  cat(sprintf('Narrowing by years experience: %i, yeilding %i samples from %i samples\n', years,nrow(toReturn),start_len))
  
  return(toReturn)
}

#' Filter by Date Completed
#' 
#' Filters the survey by the date complete, accepting all dates
#' LESS THAN the provided date
#'
#' @param data The data frame on which to filter
#' @param date The date on the top end to filter by
#'
#' @return The filtered dataframe
#'
#' @examples
#' Filter out all data from 2020-09-15 and later:
#' df_out <- filter_date_completed(df_in, "2020-09-15")
filter_date_completed <- function(data, date) {
  start_len <- nrow(data)
  toReturn<-data[which(data$clinician_survey_timestamp < date), ]
  cat(sprintf('Narrowing by date completed: %s, yeilding %i samples from %i samples\n', date,nrow(toReturn),start_len))
  
  return(toReturn)
}

#' Filter by level of experience with Lil'Flo.
#'
#' Will select data which matches ANY of the criteria
#'
#' @param data The data to be filtering on
#' @param experience The levels of experience you want to end up with,
#'                   this should be a list of each level's ID. To find
#'                   out what each ID means, run:
#'                   `label(data$flo_prior_experience___<ID>`
#'
#' @return The filtered data
#' @export
#'
#' @examples data <- filter_flo_experience(data,c(1,3))
filter_flo_experience <- function(data, experience_yn) {
  start_len <- nrow(data)
  df <- data.frame(matrix(NA, ncol = 1, nrow = nrow(data)))[-1]
  for (idx in seq_along(experience_yn)) {
    df<-cbind(data[sprintf('flo_prior_experience___%i', experience_yn[idx])], df)
  }
  toReturn<-data[rowSums(df) > 0L, ]
  cat(sprintf('Narrowing by experience with flo: %s, yeilding %i samples from %i samples\n', experience_yn,nrow(toReturn),start_len))
  
  return(toReturn)
}

## Filter to relevant population ## 
# Only Physiatrists:
# A filter:
pmnr_filter<-stringr::str_detect(data$occupation_doctor_type,  stringr::regex('physiatrist|pmr|pmnr|pm&r|physical medicine and rehab|rehabilitation physician|rehabilitation medicine', ignore_case = T))

# Who gets left behind
lb<-data %>% filter(!pmnr_filter)%>% select(occupation_doctor_type)
print(lb[lb$occupation_doctor_type!="",])

# Who we keep
data<-data%>% filter(pmnr_filter) 
print(data%>%select(occupation_doctor_type))

### Save RDA file ###
data_out <-
  data %>% select(
    occupation_doctor_type, 
    age,
    years_experience,
    contains('gender'),
    matches("^location_type___"),
    matches("^location_country"),
    matches("^location___"),
    location_state.factor,
    contains('patient_pop'),
    contains('covid_telepresence___'),
    contains('past_telepresence___'),
    clinician_survey_timestamp,
    occupation.factor,
    matches("^video_future"),
    matches("^activities_d_"),
    matches("^activities_b_")
  )
saveRDS(
  list(
    filtered_data=data_out
  ),
  file='./paper_notebooks/physiatrists-covid.Rda'
)
write.csv(data_out,"./paper_notebooks/physiatrists-covid.csv", row.names = FALSE)
```
