## Supplementary material for "Insights on Telemedicine Use by Physiatrists Before, During, and Beyond the COVID-19 Pandemic": physiatrists-covid.html

- Libraries
- Useful Functions
  - Data Filtering
  - Plotting
  - “Relative Risk”
    - McNemar Exact Test
    - Confidence Intervals
- Load Data
- Demographics
  - Number of responses:
  - Types of Clinicians
  - Age
  - Years of Experience
  - Gender
  - Clinical Setting
  - Location
  - Location Type
  - Dates of Responses
- Patient Demographics
  - Patient Type
  - Cognitive Impairment
  - Motor Impairment
  - Age Group
  - Phase of Care
- H1: physiatrists use more telepresence during than before covid
  - Phone calls with patients
  - Video to another clinical setting
  - Video to home
  - Text based communication
  - Add significance to plot
- H2: COVID-19 changed whether clinicians will use video based telehealth going forward
- Q1: What have/could physiatrists used telehealth for?
  - Have Done
  - Believe Could Be Done
  - Compare

### Libraries

```
library(dplyr)
```

```
## 
## Attaching package: 'dplyr'
```

```
## The following objects are masked from 'package:stats':
## 
##     filter, lag
```

```
## The following objects are masked from 'package:base':
## 
##     intersect, setdiff, setequal, union
```

```
library(ggplot2)
library(ggpubr)
library(Hmisc)
```

```
## Loading required package: lattice
```

```
## Loading required package: survival
```

```
## Loading required package: Formula
```

```
## 
## Attaching package: 'Hmisc'
```

```
## The following objects are masked from 'package:dplyr':
## 
##     src, summarize
```

```
## The following objects are masked from 'package:base':
## 
##     format.pval, units
```

### Useful Functions

#### Data Filtering

```
#' Condense Multiple Choice Responses
#'
#' @param data The raw dataframe to work with
#' @param root The question sequence root id (this is the ID in REDCap)
#' @param range The range of responses, these are the IDs of the response options
#'
#' @return The data with a single column (field) containing the responses to the question
condense_multi_choice <- function(data, root, range) {
  sub_data_list <- list()
  sub_data_fields <- sprintf("%s___%i", root, range)

  factor_list <- list()

  for (field_idx in seq_along(sub_data_fields)) {
    sub_data <- data[, sub_data_fields[field_idx]]
    sub_data_list[[field_idx]] <-
      rep(Hmisc::label(sub_data), times = sum(sub_data))

    factor_list[[field_idx]] <-
      as.character(label(data[, sub_data_fields[field_idx]]))
  }

  df_sub_data <- as.data.frame(unlist(sub_data_list))
  colnames(df_sub_data) <- "field"
  df_sub_data$field <- factor(df_sub_data$field, levels = factor_list)
  return(df_sub_data)
}
```

#### Plotting

```
plot_simple_bar <- function(data, field, title, subtitle) {
  if (missing(subtitle)) {
    labels <- labs(x = "", y = "Frequency", title = title)
  } else {
    labels <-
      labs(
        x = "",
        y = "Frequency",
        title = title,
        subtitle = subtitle
      )
  }
  ggplot(data, aes(x = .data[[field]])) +
    geom_bar() +
    geom_text(stat = "count", aes(label = ..count..), hjust = -.1) +
    coord_flip() +
    labels +
    theme_classic2()
}

plot_multi_choice <- function(data, root, range, title) {
  df_sub_data <- condense_multi_choice(data, root, range)

  ggplot(df_sub_data, aes(x = field)) +
    geom_bar() +
    geom_text(stat = "count", aes(label = ..count..), hjust = -.1) +
    coord_flip() +
    labs(
      x = "",
      y = "Frequency",
      title = title,
      subtitle = "Multiple Choice"
    ) +
    theme_classic2()
}

plt_telepresence_options <- function(data, colnames, rng, nme) {
  # done_data<-data%>%select(matches('activities_d_'))%>%select(!matches('factor'))
  cond_data <- list()
  for (i in seq_along(colnames)) {
    col <- colnames[[i]]
    # plot <-
    #   plot_multi_choice(data, col, c(1, 3, 4, 5, 6), fields[substr(col,14,9999), ])
    # print(plot)
    gg <- fields[substr(col, 14, 9999), ]
    df_sub_data <- condense_multi_choice(data, col, rng)
    df_sub_data$act <- gg
    cond_data[[i]] <- df_sub_data
  }
  full_data <- do.call("rbind", cond_data)

  opt1 <- ggplot(full_data, aes(x = act, fill = field)) +
    geom_bar(position = position_dodge(.8), width = .6) +
    geom_text(
      stat = "count",
      aes(label = ..count..),
      hjust = -.5,
      position = position_dodge(width = .8)
    ) +
    coord_flip() +
    labs(
      x = "",
      y = "Frequency"
    ) +
    theme_classic2() +
    scale_x_discrete(
      labels = function(x) {
        stringr::str_wrap(x, width = 20)
      }
    ) +
    theme(
      legend.title = element_blank(),
      legend.position = "bottom"
    ) +
    guides(fill = guide_legend(reverse = TRUE, ncol = 2))

  opt2 <- ggplot(full_data, aes(x = field, fill = act)) +
    geom_bar(position = position_dodge(.9), width = .5) +
    geom_text(
      stat = "count",
      aes(label = ..count..),
      hjust = -.5,
      position = position_dodge(width = .9)
    ) +
    coord_flip() +
    labs(
      x = "",
      y = "Frequency"
    ) +
    theme_classic2() +
    scale_x_discrete(
      labels = function(x) {
        stringr::str_wrap(x, width = 20)
      }
    ) +
    theme(
      legend.title = element_blank(),
      legend.position = "bottom"
    ) +
    guides(fill = guide_legend(reverse = TRUE, ncol = 2))
  print(opt1)
  print(opt2)
  return(full_data)
}
```

#### “Relative Risk”

Looking at how much off axis interaction there is in a matched pairs table

##### McNemar Exact Test

We are going to use McNemar’s exact test. This is overly conservative (https://bmcmedresmethodol.biomedcentral.com/articles/10.1186/1471-2288-13-91) but it is safe to use.

```
mcnemar_exact <- function(tbl) {
  n <- tbl[1, 2] + tbl[2, 1]
  i <- seq(0, min(tbl[1, 2], tbl[2, 1]))
  2 * sum(choose(n, i) * 0.5^n)
}
```

##### Confidence Intervals

Confidence interval and test from Liddell: https://www.jstor.org/stable/25566382

```
liddell <- function(tbl, alpha = 0.05) {
  t <- tbl[1, 1]
  r <- tbl[1, 2]
  s <- tbl[2, 1]
  u <- tbl[2, 2]
  F_l <- qf(alpha / 2, 2 * (s + 1), 2 * r, lower.tail = FALSE)
  F_u <- qf(alpha / 2, 2 * (r + 1), 2 * s, lower.tail = FALSE)
  lower <- r / ((s + 1) * F_l)
  upper <- ((r + 1) * F_u) / s
  F <- r / (s + 1)
  p <- 2 * pf(r / ((s + 1)), 2 * (s + 1), 2 * r, lower.tail = FALSE)

  df <-
    data.frame(
      row.names = c("p", "lower confidence", "upper confidence", "alpha"),
      val = c(p, lower, upper, alpha)
    )
  return(df)
}
```

### Load Data

Place the RDA file in the R working directory when running this script or adjust the `data_file_location` variable to point to the location of the data file.

```
data_file_location <- "physiatrists-covid.Rda"
saved_data <- readRDS(data_file_location)
data <- saved_data$filtered_data
```

### Demographics

#### Number of responses:

```
nrow(data)
```

```
## [1] 56
```

#### Types of Clinicians

```
plot_simple_bar(data, "occupation_doctor_type", "Type of Physician")
```

```
data$occupation_doctor_type
```

```
## What type of doctor are you? 
##  [1] "Peds PM&R"                                  
##  [2] "Pediatric Physiatrist"                      
##  [3] "PM&R"                                       
##  [4] "PMR"                                        
##  [5] "Physical Medicine and Rehabilitation"       
##  [6] "PM&R"                                       
##  [7] "PM&R"                                       
##  [8] "PM&R resident"                              
##  [9] "Physiatrist"                                
## [10] "PM&R"                                       
## [11] "physiatrist"                                
## [12] "physiatrist"                                
## [13] "Physical Medicine and rehabilitation "      
## [14] "Physiatrist"                                
## [15] "Rehabilitation physician"                   
## [16] "PM&R resident"                              
## [17] "PM&R"                                       
## [18] "Physiatrist"                                
## [19] "Physiatrist"                                
## [20] "PM&R"                                       
## [21] "PM&R"                                       
## [22] "Physical Medicine and Rehabilitation"       
## [23] "pmr"                                        
## [24] "PM&R resident"                              
## [25] "PM&R"                                       
## [26] "Resident- PMR"                              
## [27] "PM&R"                                       
## [28] "Physiatrist "                               
## [29] "Physiatrist"                                
## [30] "PM&R Resident"                              
## [31] "PM&R"                                       
## [32] "PM&R Resident"                              
## [33] "Physiatrist"                                
## [34] "PM&R resident"                              
## [35] "Pmr"                                        
## [36] "Resident physician (physiatrist)"           
## [37] "PM&R "                                      
## [38] "PM&R resident"                              
## [39] "PM&R "                                      
## [40] "Physical Medicine and Rehabilitation "      
## [41] "Resident Physiatrist"                       
## [42] "Resident PM&R physician "                   
## [43] "Resident in PM&R"                           
## [44] "PM&R"                                       
## [45] "Pediatric Rehabilitation Medicine Physician"
## [46] "PM&R"                                       
## [47] "Physiatrist"                                
## [48] "Physiatrist"                                
## [49] "Physiatrist "                               
## [50] "Physiatrist"                                
## [51] "Physiatrist"                                
## [52] "Physiatrist"                                
## [53] "Physiatrist"                                
## [54] "Physiatrist"                                
## [55] "MD; Specialty: PMR"                         
## [56] "Physiatrist "
```

#### Age

```
summary(data$age)
```

```
##    Min. 1st Qu.  Median    Mean 3rd Qu.    Max. 
##   26.00   30.00   33.00   35.29   39.00   60.00
```

```
sd(data$age)
```

```
## [1] 7.898709
```

#### Years of Experience

```
summary(data$years_experience)
```

```
##    Min. 1st Qu.  Median    Mean 3rd Qu.    Max. 
##   1.000   2.000   3.000   6.196   8.250  35.000
```

```
sd(data$years_experience)
```

```
## [1] 6.97116
```

#### Gender

```
plot_simple_bar(data, "gender.factor", "Gender")
```

#### Clinical Setting

```
plot_multi_choice(data, "location", 1:12, "Locations")
```

#### Location

```
plot_simple_bar(data, "location_country.factor", "State")
```

```
plot_simple_bar(data, "location_state.factor", "State")
```

#### Location Type

```
plot_multi_choice(data, "location_type", 1:3, "Location Types")
```

```
data$locat_type_permute___1 <-
  data$location_type___1 == TRUE &
    data$location_type___2 == FALSE & data$location_type___3 == FALSE
data$locat_type_permute___2 <-
  data$location_type___1 == FALSE &
    data$location_type___2 == TRUE & data$location_type___3 == FALSE
data$locat_type_permute___3 <-
  data$location_type___1 == FALSE &
    data$location_type___2 == FALSE & data$location_type___3 == TRUE
data$locat_type_permute___4 <-
  data$location_type___1 == TRUE &
    data$location_type___2 == TRUE & data$location_type___3 == FALSE
data$locat_type_permute___5 <-
  data$location_type___1 == TRUE &
    data$location_type___2 == FALSE & data$location_type___3 == TRUE
data$locat_type_permute___6 <-
  data$location_type___1 == FALSE &
    data$location_type___2 == TRUE & data$location_type___3 == TRUE
data$locat_type_permute___7 <-
  data$location_type___1 == TRUE &
    data$location_type___2 == TRUE & data$location_type___3 == TRUE

label(data$locat_type_permute___1) <- label(data$location_type___1)
label(data$locat_type_permute___2) <- label(data$location_type___2)
label(data$locat_type_permute___3) <- label(data$location_type___3)
label(data$locat_type_permute___4) <- "Urban and Suburban"
label(data$locat_type_permute___5) <- "Urban and Rural"
label(data$locat_type_permute___6) <- "Suburban and Rural"
label(data$locat_type_permute___7) <- "Urban and Suburban and Rural"

plot_multi_choice(data, "locat_type_permute", 1:7, "Location Types")
```

#### Dates of Responses

```
data %>%
  select(clinician_survey_timestamp) %>%
  summary()
```

```
##  clinician_survey_timestamp
##  Min.   :2020-06-30        
##  1st Qu.:2020-08-15        
##  Median :2020-09-23        
##  Mean   :2020-09-18        
##  3rd Qu.:2020-10-05        
##  Max.   :2021-01-10
```

### Patient Demographics

#### Patient Type

Please select descriptors for the type of patients you treat. Patients with..

```
plot_multi_choice(data, "patient_pop", 1:15, "Patient Type")
```

#### Cognitive Impairment

What are the levels of COGNITIVE impairment that your patients have?

```
plot_multi_choice(data, "patient_pop_cog", 1:4, "Patient Cognitive Levels")
```

#### Motor Impairment

What are the levels of MOTOR impairment that your patients have?

```
plot_multi_choice(data, "patient_pop_motor", 1:4, "Patient Motor Levels")
```

#### Age Group

What age groups do you generally work with?

```
plot_multi_choice(data, "patient_pop_age", 1:9, "Patient Ages")
```

#### Phase of Care

What phase of care are your patients in?

```
plot_multi_choice(data, "patient_pop_phase", 1:3, "Patient Treatment Phase")
```

### H1: physiatrists use more telepresence during than before covid

To do this, we will use a paired proportions test. We will treat COVID as a treatment. We can test each question as a true/false independently of each other without a risk of increasing Type I error. However, doing it this way prevents us from making a general claim about “telepresence usage increasing”. We will instead be saying that “video calls to patients at home increased”.

Effect size ratings come from here: Cohen, J. 1988. Statistical Power Analysis for the Behavioral Sciences, 2nd Edition. Routledge.

I am following: - https://rcompanion.org/handbook/H\_05.html - https://www-jstor-org.proxy.library.upenn.edu/stable/25566382 - https://www.statsdirect.com/help/chi\_square\_tests/mcnemar.htm - https://bmcmedresmethodol.biomedcentral.com/articles/10.1186/1471-2288-13-91

**Question:** Do you use any of these tools to interact with patients during normal practice (prior to the COVID-19 pandemic)? AND Are you using any of these tools to connect with your patients during the COVID-19 pandemic?

```
tele_use_past <-
  condense_multi_choice(data, "past_telepresence", 1:6) %>%
  mutate(time = "Prior to COVID-19")
tele_use_covid <-
  condense_multi_choice(data, "covid_telepresence", 1:6) %>%
  mutate(time = "During COVID-19")

tele_use_agg <- rbind(tele_use_past, tele_use_covid)

plt <- ggplot(tele_use_agg, aes(x = field)) +
  geom_bar(aes(fill = time), position = "dodge", colour = "black") +
  geom_text(
    stat = "count",
    aes(fill = time, label = ..count..),
    hjust = -.5,
    position = position_dodge(width = 1),
  ) +
  coord_flip() +
  labs(
    x = "",
    y = "Frequency",
    color = "black"
  ) +
  theme_classic2() +
  scale_x_discrete(
    labels = function(x) {
      stringr::str_wrap(x, width = 28)
    }
  ) +
  theme(
    legend.title = element_blank(),
    legend.position = c(.85, .9),
    legend.background = element_rect(fill = alpha("blue", 0.0)),
    axis.text = element_text(color = "black"),
    plot.margin = unit(integer(4) + 1, "mm")
  ) +
  guides(fill = guide_legend(reverse = TRUE)) +
  scale_fill_grey(start = .3, end = .8)
```

```
## Warning: Ignoring unknown aesthetics: fill
```

```
print(plt)
```

```
field <- c()
p <- c()
y.position <- c()
```

#### Phone calls with patients

Make sure we have the right data:

```
label(data$past_telepresence___1)
```

```
## [1] "Phone calls with patients"
```

```
label(data$covid_telepresence___1)
```

```
## [1] "Phone calls with patients"
```

Build contingency table:

```
pc_tbl <-
  table(data$past_telepresence___1,
    data$covid_telepresence___1,
    dnn = c("prior", "covid")
  )
pc_tbl
```

```
##      covid
## prior  0  1
##     0  4 10
##     1  1 41
```

```
res <- liddell(pc_tbl)
res
```

**Significant change due to COVID**

We also need effect size:

```
rcompanion::cohenG(pc_tbl)
```

```
## $Global.statistics
##   Dimensions OR     P     g
## 1      2 x 2 10 0.909 0.409
```

By both Cohens g and odds ratio, this is a **large effect**

And some plotting values:

```
field <- append(field, label(data$covid_telepresence___1))
p <- append(p, res$val[[1]])
y.position <-
  append(y.position, max(
    sum(data$past_telepresence___1),
    sum(data$covid_telepresence___1)
  ))
```

#### Video to another clinical setting

Make sure we have the right data:

```
label(data$past_telepresence___4)
```

```
## [1] "Video calls to patients in another clinical setting"
```

```
label(data$covid_telepresence___4)
```

```
## [1] "Video calls to patients in another clinical setting"
```

Build contingency table:

```
vac_tbl <-
  table(data$past_telepresence___4,
    data$covid_telepresence___4,
    dnn = c("prior", "covid")
  )
vac_tbl
```

```
##      covid
## prior  0  1
##     0 45  7
##     1  0  4
```

```
res <- liddell(vac_tbl)
```

```
## Warning in qf(alpha/2, 2 * (r + 1), 2 * s, lower.tail = FALSE): NaNs produced
```

```
res
```

**Significant change due to COVID** Note, due to the zero on the prior true, covid false point, the upper confidence interval cannot be calculated.

We also need effect size:

```
rcompanion::cohenG(vac_tbl)
```

```
## $Global.statistics
##   Dimensions  OR P   g
## 1      2 x 2 Inf 1 0.5
```

Cohen’s g shows a **large effect**. Odds ratio again, can’t really be calculated

And some plotting values:

```
field <- append(field, label(data$covid_telepresence___4))
p <- append(p, res$val[[1]])
y.position <- append(y.position, max(sum(data$past_telepresence___4), sum(data$covid_telepresence___4)))
```

#### Video to home

Make sure we have the right data:

```
label(data$past_telepresence___3)
```

```
## [1] "Video calls to patients at home"
```

```
label(data$covid_telepresence___3)
```

```
## [1] "Video calls to patients at home"
```

Build contingency table:

```
vh_tbl <-
  table(data$past_telepresence___3,
    data$covid_telepresence___3,
    dnn = c("prior", "covid")
  )
vh_tbl
```

```
##      covid
## prior  0  1
##     0 11 24
##     1  2 19
```

```
res <- liddell(vh_tbl)
res
```

**Significant change due to COVID**

We also need effect size:

```
rcompanion::cohenG(vh_tbl)
```

```
## $Global.statistics
##   Dimensions OR     P     g
## 1      2 x 2 12 0.923 0.423
```

By both Cohens g and odds ratio, this is a **large effect**

And some plotting values:

```
field <- append(field, label(data$covid_telepresence___3))
p <- append(p, res$val[[1]])
y.position <-
  append(y.position, max(
    sum(data$past_telepresence___3),
    sum(data$covid_telepresence___3)
  ))
```

#### Text based communication

Make sure we have the right data:

```
label(data$past_telepresence___2)
```

```
## [1] "Text based communication"
```

```
label(data$covid_telepresence___2)
```

```
## [1] "Text based communication"
```

Build contingency table:

```
tc_tbl <-
  table(data$past_telepresence___2,
    data$covid_telepresence___2,
    dnn = c("prior", "covid")
  )
tc_tbl
```

```
##      covid
## prior  0  1
##     0 36  3
##     1  2 15
```

```
res <- liddell(tc_tbl)
res
```

**Not Significant**

And some plotting values:

```
field <- append(field, label(data$covid_telepresence___2))
p <- append(p, res$val[[1]])
y.position <-
  append(y.position, max(
    sum(data$past_telepresence___2),
    sum(data$covid_telepresence___2)
  ))
```

#### Add significance to plot

```
group1 <- rep("Prior to COVID-19", 4)
group2 <- rep("During COVID-19", 4)
sig <- ifelse(p < .05, "*", "ns")
plot_params <- ggplot_build(plt)
plt_levels <- levels(plot_params$plot$data$field)
x_pos <- sapply(field, function(x) {
  match(x, plt_levels)
})
stat.test <-
  data.frame(
    field,
    group1,
    group2,
    p,
    y.position = y.position + 5,
    sig,
    xmin = x_pos - .3,
    xmax = x_pos + .3
  )

plt <- plt + stat_pvalue_manual(
  stat.test,
  label = "sig",
  tip.length = 0.0,
  coord.flip = TRUE
)

print(plt)
```

Also create aggregate plot:

```
tele_use_compressed_past <-
  data %>%
  mutate(video = ifelse(
    past_telepresence___3 |
      past_telepresence___4,
    "Video based telepresence",
    0
  )) %>%
  mutate(
    other = ifelse(
      past_telepresence___1 |
        past_telepresence___2 |
        past_telepresence___5,
      "Non-Video based telepresence",
      0
    )
  ) %>%
  mutate(none = ifelse(past_telepresence___6, "No telepresence", 0)) %>%
  select(other, none, video) %>%
  tidyr::gather("key", "field") %>%
  filter(field != 0) %>%
  select(field) %>%
  mutate(time = "Prior to COVID-19")
```

```
## Warning: attributes are not identical across measure variables;
## they will be dropped
```

```
tele_use_compressed_covid <-
  data %>%
  mutate(
    video = ifelse(
      covid_telepresence___3 |
        covid_telepresence___4,
      "Video based telepresence",
      0
    )
  ) %>%
  mutate(
    other = ifelse(
      covid_telepresence___1 |
        covid_telepresence___2 |
        covid_telepresence___5,
      "Non-Video based telepresence",
      0
    )
  ) %>%
  mutate(none = ifelse(covid_telepresence___6, "No telepresence", 0)) %>%
  select(other, none, video) %>%
  tidyr::gather("key", "field") %>%
  filter(field != 0) %>%
  select(field) %>%
  mutate(time = "During COVID-19")
```

```
## Warning: attributes are not identical across measure variables;
## they will be dropped
```

```
tele_use_compressed <- rbind(tele_use_compressed_past, tele_use_compressed_covid)

plt_cond <-
  ggplot(tele_use_compressed, aes(x = forcats::fct_infreq(field))) +
  geom_bar(aes(fill = time), position = "dodge", colour = "black") +
  geom_text(
    stat = "count",
    aes(fill = time, label = ..count..),
    hjust = -.5,
    position = position_dodge(width = 1)
  ) +
  coord_flip() +
  labs(
    x = "",
    y = "Frequency",
    color = "black"
  ) +
  theme_classic2() +
  scale_x_discrete(
    labels = function(x) {
      stringr::str_wrap(x, width = 28)
    }
  ) +
  theme(
    legend.title = element_blank(),
    legend.position = c(.85, .9),
    legend.background = element_rect(fill = alpha("blue", 0.0)),
    axis.text = element_text(color = "black"),
    plot.margin = unit(integer(4) + 1, "mm")
  ) +
  guides(fill = guide_legend(reverse = TRUE)) +
  scale_fill_grey(start = .3, end = .8)
```

```
## Warning: Ignoring unknown aesthetics: fill
```

```
print(plt_cond)
```

```
plt_mod_all <- plt + labs(x = "", y = "")
plt_mod_cond <- plt_cond + theme(legend.position = "none")
x_max <- max(c(
  layer_scales(plt_mod_all)$y$range$range[[2]],
  layer_scales(plt_mod_cond)$y$range$range[[2]]
))
plt_mod_all <- plt_mod_all + scale_y_continuous(limits = c(0, x_max))
plt_mod_cond <- plt_mod_cond + scale_y_continuous(limits = c(0, x_max))
plt_grid <-
  cowplot::plot_grid(
    plt_mod_all,
    plt_mod_cond,
    align = "hv",
    ncol = 1,
    rel_heights = c(1, 0.6)
  )
print(plt_grid)
```

Useful to measure change in usage overall:

```
pre <- nrow(data) - (tele_use_compressed_past %>% filter(field == "No telepresence") %>% count())
post <- nrow(data) - (tele_use_compressed_covid %>% filter(field == "No telepresence") %>% count())

change <- (post - pre) / pre
change
```

and for video:

```
pre <- tele_use_compressed_past %>%
  filter(field == "Video based telepresence") %>%
  count()
post <- tele_use_compressed_covid %>%
  filter(field == "Video based telepresence") %>%
  count()

change <- (post - pre) / pre
change
```

### H2: COVID-19 changed whether clinicians will use video based telehealth going forward

Do you plan to use video calls with your patients for your practice after the COVID-19 pandemic has ended?

Group all of the ways that someone can do video pre-COVID

```
data <-
  data %>% mutate(pre_covid_video = past_telepresence___3 |
    past_telepresence___4)
data %>%
  select(pre_covid_video) %>%
  summary()
```

```
##  pre_covid_video
##  Mode :logical  
##  FALSE:35       
##  TRUE :21
```

Group all the ways someone can do video during COVID:

```
data <-
  data %>% mutate(covid_video = covid_telepresence___3 |
    covid_telepresence___4)
data %>%
  select(covid_video) %>%
  summary()
```

```
##  covid_video    
##  Mode :logical  
##  FALSE:13       
##  TRUE :43
```

Look at what changed before COVID to now

```
table(
  data$pre_covid_video,
  data$covid_video,
  dnn = c("Pre-COVID Usage", "During COVID Usage")
)
```

```
##                During COVID Usage
## Pre-COVID Usage FALSE TRUE
##           FALSE    11   24
##           TRUE      2   19
```

So a few people who used video before COVID are not using it during COVID (2). But many people who did not use it before are now (24).

What are people’s plans going forward:

```
data$video_future.factor %>% summary()
```

```
##      Yes       No Not Sure 
##       44        3        9
```

How does that look compared to people from pre-COVID?

```
pre_future_vid <-
  table(
    data$pre_covid_video,
    data$video_future.factor,
    dnn = c("Pre-COVID Usage", "Future Usage")
  )
pre_future_vid
```

```
##                Future Usage
## Pre-COVID Usage Yes No Not Sure
##           FALSE  28  1        6
##           TRUE   16  2        3
```

Wow, that is very few no’s for future usage.

Look at people who have been using during COVID compared to future usage:

```
table(
  data$covid_video,
  data$video_future.factor,
  dnn = c("Usage During COVID", "Future Usage")
)
```

```
##                   Future Usage
## Usage During COVID Yes No Not Sure
##              FALSE   8  1        4
##              TRUE   36  2        5
```

It is interesting that so many people who are not using video during COVID are planning to use it in the future.

Let’s try to visualize all 3 time periods at once

```
ftable(
  data$pre_covid_video,
  data$covid_video,
  data$video_future.factor,
  dnn = c("Usage Pre COVID", "Usage During COVID", "Future Usage")
)
```

```
##                                    Future Usage Yes No Not Sure
## Usage Pre COVID Usage During COVID                             
## FALSE           FALSE                             7  1        3
##                 TRUE                             21  0        3
## TRUE            FALSE                             1  0        1
##                 TRUE                             15  2        2
```

Ahh, that is nice. Ok, so interpretation:

- only 3 subjects are definitely not using video post-COVID, that compares to 35 who weren’t before
- 1 of the subjects who used video before but not during COVID are planning to use it in the future
- Strangely 2 who used it before COVID and during COVID don’t plan to use it going forward and 2 aren’t sure

So what can we test exactly? We could look at just people who did not use video before and see did COVID convince more people to use telepresence. But that seems a bit dishonest since some people won’t be using telepresence post-COVID. So I think the best thing is to look at pre-COVID to future plans and just use the table above in an explanatory fashion, not tested.

So then the question is how to get it to be 2x2 so that we can use McNemar’s or similar test. We need to condense Yes, No, and Not Sure down. There are 9 not sure responses. Options are to exclude those respondents or to roll them into one of the other categories. I think the best thing to do is to roll them into the no for future use, so that we are generating a very conservative estimate (some will end up no in the future and some yes).

```
compressed_future <-
  data %>%
  mutate(vid_future_comp = video_future.factor == "Yes") %>%
  select(vid_future_comp)
future_vid_use<-compressed_future %>% table()
future_vid_use
```

```
## .
## FALSE  TRUE 
##    12    44
```

```
future_vid_use[2]/sum(future_vid_use)
```

```
##      TRUE 
## 0.7857143
```

```
pre_future_vid_comp <-
  table(
    data$pre_covid_video,
    compressed_future$vid_future_comp,
    dnn = c("Pre-COVID Usage", "Future Usage")
  )
pre_future_vid_comp
```

```
##                Future Usage
## Pre-COVID Usage FALSE TRUE
##           FALSE     7   28
##           TRUE      5   16
```

```
liddell(pre_future_vid_comp)
```

**Significant change due to COVID**

We also need effect size:

```
rcompanion::cohenG(pre_future_vid_comp)
```

```
## $Global.statistics
##   Dimensions  OR     P     g
## 1      2 x 2 5.6 0.848 0.348
```

**Large Effect Size**

And let’s also get percent who will use going forward:

```
sum(pre_future_vid_comp[,2])/nrow(data)
```

```
## [1] 0.7857143
```

### Q1: What have/could physiatrists used telehealth for?

Some setup:

```
fields <-
  data.frame(
    row.names = c(
      "motor_ass",
      "stretching",
      "strength",
      "adl",
      "cog_ass",
      "cog_ex",
      "env_adap",
      "orthotics",
      "surg",
      "rads",
      "med"
    ),
    val = c(
      "Motor Assessments",
      "Stretching",
      "Strength Building",
      "ADL Practice",
      "Cognitive Assessments",
      "Cognitive Exercises",
      "Environmental Adaptation",
      "Orthotics Assessment/Prescription",
      "Discussions about Surgery",
      "Discussions about Radiology Results",
      "Medical Prescriptions"
    )
  )
```

#### Have Done

Which types of activities HAVE YOU DONE with each type of tool?

Non-video remote communication includes: phone calls, text messages, email, instant messages, and other types of communication which allow you to interact with patients from afar, without using video.

```
colnames_wnumbers <-
  names(data)[grepl("activities_d_.*1", names(data)) &
    !grepl("factor", names(data))]
colnames <-
  substr(colnames_wnumbers, 1, nchar(colnames_wnumbers) - 4)

hd_data <-
  plt_telepresence_options(data, colnames, c(1, 3, 5, 6), "have_done")
```

#### Believe Could Be Done

Which types of activities DO YOU BELIEVE YOU COULD DO with each type of tool?

Non-video remote communication includes: phone calls, text messages, email, instant messages, and other types of communication which allow you to interact with patients from afar, without using video.

```
colnames_wnumbers <-
  names(data)[grepl("activities_b_.*1", names(data)) &
    !grepl("factor", names(data))]
colnames <-
  substr(colnames_wnumbers, 1, nchar(colnames_wnumbers) - 4)

cd_data <-
  plt_telepresence_options(data, colnames, c(1, 3, 5), "could_do")
```

#### Compare

```
knitr::kable(table(hd_data))
```

|  | ADL Practice | Cognitive Assessments | Cognitive Exercises | Discussions about Radiology Results | Discussions about Surgery | Environmental Adaptation | Medical Prescriptions | Motor Assessments | Orthotics Assessment/Prescription | Strength Building | Stretching |
| --- | --- | --- | --- | --- | --- | --- | --- | --- | --- | --- | --- |
| Non-Video Remote Interaction | 3 | 10 | 3 | 30 | 21 | 4 | 38 | 3 | 5 | 4 | 4 |
| Video Call Based Remote Interaction | 8 | 31 | 9 | 35 | 30 | 11 | 39 | 36 | 16 | 13 | 20 |
| In Person | 21 | 41 | 16 | 42 | 39 | 16 | 45 | 49 | 33 | 32 | 37 |
| Have Not Done | 32 | 9 | 36 | 5 | 11 | 36 | 1 | 2 | 20 | 21 | 16 |

```
knitr::kable(table(cd_data))
```

|  | ADL Practice | Cognitive Assessments | Cognitive Exercises | Discussions about Radiology Results | Discussions about Surgery | Environmental Adaptation | Medical Prescriptions | Motor Assessments | Orthotics Assessment/Prescription | Strength Building | Stretching |
| --- | --- | --- | --- | --- | --- | --- | --- | --- | --- | --- | --- |
| Non-Video Remote Interaction | 3 | 24 | 19 | 37 | 37 | 6 | 40 | 4 | 8 | 9 | 7 |
| Video Call Based Remote Interaction | 46 | 53 | 52 | 52 | 52 | 41 | 51 | 41 | 36 | 44 | 41 |
| Could Not Be Done Remotely | 6 | 1 | 2 | 1 | 1 | 10 | 2 | 10 | 20 | 9 | 11 |

Create flat contingency table to directly compare. Note:

- In Person, Could Do was not asked, all zero
- Have Not Done, Have Done is just Have Not Done
- Have Not Done, Could Do was not asked, all zero
- Could Not Be Done Remotely, Could Do is Could not be done remotely
- Could Not Be Done Remotely, Have Done was not asked, all zero

```
cd_data$class <- "Could Do"
hd_data$class <- "Have Done"
all_data <- bind_rows(hd_data, cd_data)
knitr::kable(format(ftable(table(all_data), row.vars = "act")))
```

|  |  |  |  |  |  |  |  |  |  |  |  |
| --- | --- | --- | --- | --- | --- | --- | --- | --- | --- | --- | --- |
|  | “field” | “Non-Video Remote Interaction” |  | “Video Call Based Remote Interaction” |  | “In Person” |  | “Have Not Done” |  | “Could Not Be Done Remotely” |  |
|  | “class” | “Could Do” | “Have Done” | “Could Do” | “Have Done” | “Could Do” | “Have Done” | “Could Do” | “Have Done” | “Could Do” | “Have Done” |
| “act” |  |  |  |  |  |  |  |  |  |  |  |
| “ADL Practice” |  | 3 | 3 | 46 | 8 | 0 | 21 | 0 | 32 | 6 | 0 |
| “Cognitive Assessments” |  | 24 | 10 | 53 | 31 | 0 | 41 | 0 | 9 | 1 | 0 |
| “Cognitive Exercises” |  | 19 | 3 | 52 | 9 | 0 | 16 | 0 | 36 | 2 | 0 |
| “Discussions about Radiology Results” |  | 37 | 30 | 52 | 35 | 0 | 42 | 0 | 5 | 1 | 0 |
| “Discussions about Surgery” |  | 37 | 21 | 52 | 30 | 0 | 39 | 0 | 11 | 1 | 0 |
| “Environmental Adaptation” |  | 6 | 4 | 41 | 11 | 0 | 16 | 0 | 36 | 10 | 0 |
| “Medical Prescriptions” |  | 40 | 38 | 51 | 39 | 0 | 45 | 0 | 1 | 2 | 0 |
| “Motor Assessments” |  | 4 | 3 | 41 | 36 | 0 | 49 | 0 | 2 | 10 | 0 |
| “Orthotics Assessment/Prescription” |  | 8 | 5 | 36 | 16 | 0 | 33 | 0 | 20 | 20 | 0 |
| “Strength Building” |  | 9 | 4 | 44 | 13 | 0 | 32 | 0 | 21 | 9 | 0 |
| “Stretching” |  | 7 | 4 | 41 | 20 | 0 | 37 | 0 | 16 | 11 | 0 |
