## Supplementary material for "Insights on Telemedicine Use by Physiatrists Before, During, and Beyond the COVID-19 Pandemic": README.md

# Supplemental Files for Insights on Telecommunication Use by Physiatrists before, during, and beyond COVID-19

## Survey Files

### `RehabilitationAndTelemedicineT.pdf`

A print out of the survey administered
Note: this is not identical to the web interface
used but shows all of the questions and options fully

### `RehabilitationAndTelemedicineT-design.pdf`

This is the designer view of the survey from REDCap. This
view shows the form types used, branching, etc.

### `RehabilitationAndTelemedicineT-dictionary`

Dictionary of questions in survey.

## Data Analysis Files

These files are meant to be used with R. It is easiest
to do this with R Studio. They can however be opened 
with any standard text editor. 

### `physiatrists-covid-data_export.R`

This is the file used to generate the data file attached in
the supplementary data. You cannot run this file, it is
provided for reference only. To generate the data file
which you are provided, the authors pass the raw data
exported directly from REDCap to this file.

### `physiatrists-covid-data_export.R-output.txt`

The output from running `physiatrists-covid-data_export.R`
provided for reference.

### `physiatrists-covid.Rda`
The data file for this work. This contains all of the data
needed for this project. [For Preprints: contact authors to
access this file]

### `physiatrists-covid.csv`
The same data as `physiatrists-covid.Rda` but in csv format
[For Preprints: contact the authors to access this file]

### `physiatrists-covid.Rmd`
The actual data analysis notebook. It is best run from within
R studio.

### `physiatrists-covid.html`
A knited html version of `physiatrists-covid.Rmd` containing 
the source document with output in a html document. Can be
opened by any modern web browser to view data analysis.
