## Supplementary material for "Insights on Telemedicine Use by Physiatrists Before, During, and Beyond the COVID-19 Pandemic": RehabilitationAndTelemedicineT-design.pdf

Penn Medicine

Penn Medicine
Institute for Translational Medicine and Therapeutics (ITMAT)

### Flo Clinician Surveys PID 22692

Online Designer

**Since this project is currently in PRODUCTION, changes will not be made in real time.** Tell me more

Submit Changes for Review

Fields to be added: **0** / Total resulting field count: **118**
Fields to be deleted: **0** / Existing field count: **118**

Remove all drafted changes

View detailed summary of all drafted changes

Create snapshot of instruments

VIDEO: How to use this page

Last snapshot: 06/03/2020 6:28pm ?

This page allows you to build and customize your data collection instruments one field at a time. You may add new fields or edit existing ones. New fields may be added by clicking the Add Field buttons. You can begin editing an existing field by clicking on the **Edit** icon. If you decide that you do not want to keep a field, you can simply delete it by clicking on the **Delete** icon. To reorder the fields, simply **drag and drop** a field to a different position within the form below.

Learn how to use [⚡] Smart Variables | Piping | @ Action Tags | Field embedding

Return to list of instruments | Survey settings

Current instrument: **Clinician Survey**

Preview instrument

*Variable:* record_id

* This field will NOT be displayed on the survey page.
* You should NOT use identifiers (e.g., MRN, SSN) for the record ID field.

**Record ID**

NOTE: The field above is the record ID field and thus cannot be deleted or moved. It can only be edited.

Add Field | Add Matrix of Fields

**Progress:**

,

**This section of the survey will ask about you and your practice as it runs normally (prior to the COVID-19 pandemic).**

Add Field | Add Matrix of Fields

*Variable:* occupation

**Are you:**

* must provide value

- A Therapist
- A Nurse
- A Doctor
- A Medical Technologist
- Other type of clinician providing rehabilitation services

reset

Add Field | Add Matrix of Fields

*Variable:* occupation_therapist_type *Branching logic:* [occupation] = '1'

**What type of therapist are you?**

* must provide value

Add Field | Add Matrix of Fields

*Variable:* occupation_nurse_type *Branching logic:* [occupation] = '2'

**What type of nurse are you?**

* must provide value

Add Field | Add Matrix of Fields

*Variable:* occupation_doctor_type *Branching logic:* [occupation] = '3'

**What type of doctor are you?**

* must provide value

Add Field | Add Matrix of Fields

*Variable:* occupation_medtech_type *Branching logic:* [occupation] = '4'

**What type of medical technologist are you?**

* must provide value

Add Field | Add Matrix of Fields

*Variable:* occupation_other_type *Branching logic:* [occupation] = '5'

**What type of clinician are you?**

* must provide value

Add Field | Add Matrix of Fields

*Variable:* age

**What is your age (years)**

* must provide value

Add Field | Add Matrix of Fields

*Variable:* gender

**What is your gender?**

* must provide value

- ○ Male
- ○ Female
- ○ Other

reset

Add Field | Add Matrix of Fields

*Variable:* location

**What type of center(s) do you work in?**
Select all that apply

* must provide value

- ☐ School
- ☐ Hospital for Children
- ☐ General Hospital
- ☐ Elder Care Hospital
- ☐ Rehab Center
- ☐ Elder Care Home
- ☐ Community Center
- ☐ Private Practice
- ☐ Patient Home
- ☐ Inpatient Facility
- ☐ Outpatient Facility
- ☐ Other

Add Field | Add Matrix of Fields

*Variable:* location_other *Branching logic:* [location(12)] = '1'

**Please specify your work location:**

* must provide value

Add Field | Add Matrix of Fields

*Variable:* location_type

**Do you work in a suburban, urban, and/or rural setting?**
Select all that apply

* must provide value

- ☐ Urban
- ☐ Suburban
- ☐ Rural

Add Field | Add Matrix of Fields

*Variable:* location_country

**What country do you primarily work in?**

* must provide value

Add Field | Add Matrix of Fields

*Variable:* location_state *Branching logic:* [location_country] = '1'

**What state do you primarily work in?**

* must provide value

Add Field | Add Matrix of Fields

*Variable:* years_experience

**How many years have you been practicing?**

* must provide value

Add Field | Add Matrix of Fields

*Variable:* patient_pop

**Please select descriptors for the type of patients you treat. Patients with..**
Select all that apply

* must provide value

- ☐ CP
- ☐ Stroke
- ☐ Brachial Plexus Injuries
- ☐ Autism
- ☐ TBI
- ☐ Cardiovascular Disease
- ☐ Speech Disorders
- ☐ Sports Injuries
- ☐ Amputations
- ☐ Arthritis
- ☐ Dementia
- ☐ Burns
- ☐ General Traumatic Injuries
- ☐ Mental Health Issues
- ☐ Other

Add Field | Add Matrix of Fields

*Variable:* patient_pop_other *Branching logic:* [patient_pop(14)] = '1'

**Please describe the other patient types who you treat:**

Expand

Add Field | Add Matrix of Fields

*Variable:* patient_pop_cog

**What are the levels of COGNITIVE impairment that your patients have?**
Select all that apply

* must provide value

- ☐ Severely Impaired
- ☐ Moderately Impaired
- ☐ Mildly Impaired
- ☐ Not Impaired

Add Field | Add Matrix of Fields

*Variable:* patient_pop_motor

**What are the levels of MOTOR impairment that your patients have?**
Select all that apply

* must provide value

- ☐ Severely Impaired
- ☐ Moderately Impaired
- ☐ Mildly Impaired
- ☐ Not Impaired

Add Field | Add Matrix of Fields

*Variable:* patient_pop_age

**What age groups do you generally work with?**
Select all that apply
* must provide value

- ☐ Infants (0-1)
- ☐ Toddlers (1-3)
- ☐ Preschoolers (3-5)
- ☐ Gradeschoolers (5-12)
- ☐ Teens (12-18)
- ☐ Young Adults (18-21)
- ☐ Adults (21-40)
- ☐ Middle Aged Adults (40-65)
- ☐ Older Adults (65+)

Add Field | Add Matrix of Fields

*Variable:* patient_pop_phase

**What phase of care are your patients in?**
Select all that apply
* must provide value

- ☐ Acute
- ☐ Sub-Acute
- ☐ Chronic

Add Field | Add Matrix of Fields

*Variable:* past_telepresence

**Do you use any of these tools to interact with patients during normal practice (prior to the COVID-19 pandemic)?**
Select all that apply
* must provide value

- ☐ Phone calls with patients
- ☐ Text based communication (email, text message, text app, etc.)
- ☐ Video calls to patients at home
- ☐ Video calls to patients in another clinical setting
- ☐ Other method of remotely connecting with patients
- ☐ None of these

Add Field | Add Matrix of Fields

*Variable:* past_telepresence_other *Branching logic:* [past_telepresence(5)] = '1'

**What is the other method you have used?**

Expand

Add Field | Add Matrix of Fields

**Progress:**
**20%**

**In this section we will ask you about your experience delivering care to your patients as a result of the COVID-19 pandemic.**

Add Field | Add Matrix of Fields

*Variable:* covid_inperson

**Have you been able to see your patients in-person during the pandemic?**
* must provide value

◯ Yes ◯ No

reset

Add Field | Add Matrix of Fields

*Variable:* covid_treatment_difficulty

**How has the pandemic changed your level of interaction with your patients?**

* must provide value

I cannot interact with my patients | I am interacting with my patients as normal | I am interacting with my patients much more than normal

Change the slider above to set a response

reset

Add Field | Add Matrix of Fields

*Variable:* covid_outcomes

**How do you expect the pandemic to affect your patients' long term health outcomes?**

* must provide value

Much worse outcomes | Unchanged outcomes | Much better outcomes

Change the slider above to set a response

reset

Add Field | Add Matrix of Fields

*Variable:* covid_patient_selfcare

**How do you believe the pandemic is affecting your patients' ability to care for themselves and meet their rehabilitation goals?**

* must provide value

Much worse | Unchanged | Much better

Change the slider above to set a response

reset

Add Field | Add Matrix of Fields

*Variable:* covid_telepresence

**Are you using any of these tools to connect with your patients during the COVID-19 pandemic?**
Select all that apply

* must provide value

- ☐ Phone calls with patients
- ☐ Text based communication (email, text message, text app, etc.)
- ☐ Video calls to patients at home
- ☐ Video calls to patients in another clinical setting
- ☐ Other method of remotely connecting with patients
- ☐ None of these

Add Field | Add Matrix of Fields

*Variable:* covid_telepresence_other *Branching logic:* [covid_telepresence(5)] = '1'

**What is the other method you are using?**

Expand

Add Field | Add Matrix of Fields

*Variable:* video_plan *Branching logic:* [covid_telepresence(3)] = '0' and [covid_telepresence(4)] = '0'

**Do you plan to use video calls to talk to your patients going forward during the COVID-19 pandemic?**

* must provide value

- ◯ Yes
- ◯ No

reset

Add Field | Add Matrix of Fields

**Progress:**

,

**In this section we will ask you about your experience and thoughts about using telemedicine, specifically video calls, both before and during the COVID-19 pandemic.**

Add Field | Add Matrix of Fields

*Variable:* video_satisfaction *Branching logic:* [past_telepresence(3)] = '1' or [past_telepresence(4)] = '1' or...

**Please rate your overall SATISFACTION using video calls for rehab**

* must provide value

Very Dissatisfied | Neutral | Very Satisfied

Change the slider above to set a response

reset

Add Field | Add Matrix of Fields

*Variable:* video_communication *Branching logic:* [past_telepresence(3)] = '1' or [past_telepresence(4)] = '1' or...

**How well are you able to COMMUNICATE with your patients over video calls compared to in-person?**

* must provide value

In-person much better communication | No difference | Video call much better communication

Change the slider above to set a response

reset

Add Field | Add Matrix of Fields

*Variable:* pvideo_communication *Branching logic:* [past_telepresence(3)] = '0' and [past_telepresence(4)] = '0' a...

**How well do you believe you could COMMUNICATE with your patients over video calls compared to in-person?**

* must provide value

In-person much better communication | No difference | Video call much better communication

Change the slider above to set a response

reset

Add Field | Add Matrix of Fields

*Variable:* video_assessment *Branching logic:* [past_telepresence(3)] = '1' or [past_telepresence(4)] = '1' or...

**How well are you able to ASSESS your patients' level of function over video calls compared to in-person?**

* must provide value

In-person much better assessment | No difference | Video call much better assessment

Change the slider above to set a response

reset

Add Field | Add Matrix of Fields

*Variable:* pvideo_assessment *Branching logic:* [past_telepresence(3)] = '0' and [past_telepresence(4)] = '0' a...

**How well do you believe you could ASSESS your patients' level of function over video calls compared to in-person?**

* must provide value

In-person much better assessment | No difference | Video call much better assessment

Change the slider above to set a response

reset

Add Field | Add Matrix of Fields

*Variable:* video_motivation *Branching logic:* [past_telepresence(3)] = '1' or [past_telepresence(4)] = '1' or...

**How MOTIVATED are your patients DURING a video call compared to in-person?**

* must provide value

In-person much more motivated | No difference | Video call much more motivated

Change the slider above to set a response

reset

Add Field | Add Matrix of Fields

*Variable:* pvideo_motivation *Branching logic:* [past_telepresence(3)] = '0' and [past_telepresence(4)] = '0' a...

**How MOTIVATED do you believe your patients would be DURING a video call compared to in-person?**

* must provide value

In-person much more motivated | No difference | Video call much more motivated

Change the slider above to set a response

reset

Add Field | Add Matrix of Fields

*Variable:* video_compliance_during *Branching logic:* [past_telepresence(3)] = '1' or [past_telepresence(4)] = '1' or...

**How well do your patients COMPLY with instructions DURING a video call vs in-person visit?**

* must provide value

In-person much higher compliance | The same | Video call much higher compliance

Change the slider above to set a response

reset

Add Field | Add Matrix of Fields

*Variable:* pvideo_compliance_during *Branching logic:* [past_telepresence(3)] = '0' and [past_telepresence(4)] = '0' a...

**How well do you believe your patients would COMPLY with instructions DURING a video call vs in-person visit?**

* must provide value

In-person much higher compliance | The same | Video call much higher compliance

Change the slider above to set a response

reset

Add Field | Add Matrix of Fields

*Variable:* video_adherence_after *Branching logic:* [past_telepresence(3)] = '1' or [past_telepresence(4)] = '1' or...

**How well do your patients ADHERE to the treatment plan AFTER a video call vs in-person visit?**

* must provide value

In-person much higher adherence | The same | Video call much higher adherence

Change the slider above to set a response

reset

Add Field | Add Matrix of Fields

*Variable:* pvideo_adherence_after *Branching logic:* [past_telepresence(3)] = '0' and [past_telepresence(4)] = '0' a...

**How well do you believe your patients would ADHERE to the treatment plan AFTER a video call vs in-person visit?**

* must provide value

In-person much higher compliance | The same | Video call much higher compliance

Change the slider above to set a response

reset

Add Field | Add Matrix of Fields

*Variable:* video_challenges *Branching logic:* [past_telepresence(3)] = '1' or [past_telepresence(4)] = '1' or...

**Do you face any challenges using video calls that affect quality of care?**
Select all that apply

- ☐ No challenges
- ☐ connection failures
- ☐ poor audio quality
- ☐ poor video quality
- ☐ Patients do not have a device for video
- ☐ Patients do not have internet
- ☐ Patients cannot understand what I want
- ☐ I cannot understand what patients want
- ☐ Patients cannot complete activities without my physical intervention
- ☐ Assessments require the use of a tool which the patient does not have
- ☐ It is not safe for patients to do the desired activities without a professional present
- ☐ Other challenges

Add Field | Add Matrix of Fields

*Variable:* pvideo_challenges *Branching logic:* [past_telepresence(3)] = '0' and [past_telepresence(4)] = '0' a...

**Do you believe you would face any challenges using video calls that would affect quality of care?**
Select all that apply

- ☐ No challenges
- ☐ Connection failures
- ☐ Poor audio quality
- ☐ Poor video quality
- ☐ Patients would not have a device for video
- ☐ Patients would not have internet
- ☐ Patients would not understand what I want
- ☐ I would not understand what patients want
- ☐ Patients could not complete activities without my physical intervention
- ☐ Assessments require the use of a tool which the patient would not have
- ☐ It would not be safe for patients to do the desired activities without a professional present
- ☐ Other challenges

Add Field | Add Matrix of Fields

*Variable:* video_challenges_other *Branching logic:* [video_challenges(12)] = '1' or [pvideo_challenges(12)] = '1'

**What other challenges?**

Expand

Add Field | Add Matrix of Fields

*Variable:* video_future

**Do you plan to use video calls with your patients for your practice after the COVID-19 pandemic has ended?**
* must provide value

- ○ Yes
- ○ No
- ○ Not Sure

reset

Add Field | Add Matrix of Fields

**Progress:**

,

**In this section we present a new robotic platform for telemedicine, specifically telerehabilitation. We then ask some questions about the system and how it could fit into your practice.**

Add Field | Add Matrix of Fields

*Variable:* flo_description

**Please watch this video which describes the Lil'Flo Robot, a socially assitive telerehabilitation robot.**

**The system consists of a social robot (a robot which interacts with people socially), which can play games, demonstrate motions, and express emotion, mounted on a mobile telepresence system which has a screen, cameras, and a microphone.**

**Then continue to answer questions about the system.**

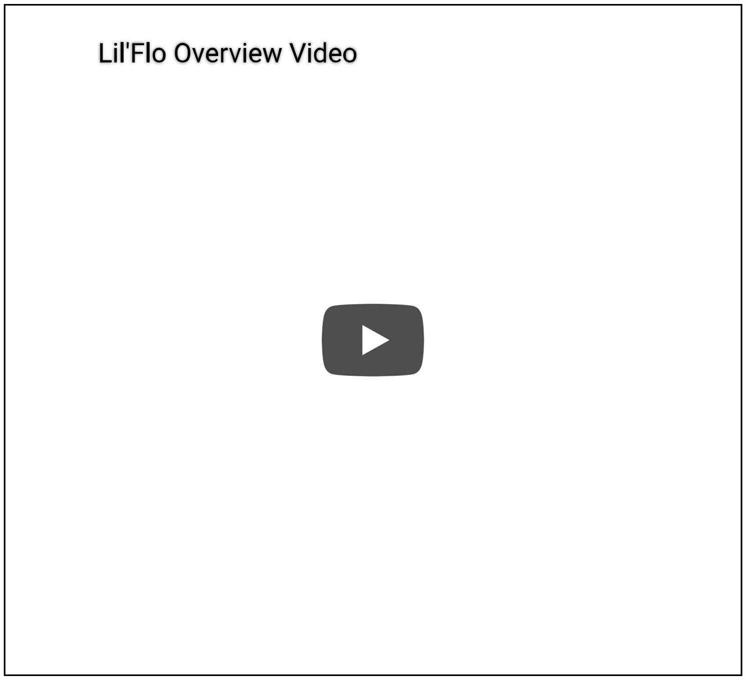


Add Field | Add Matrix of Fields

*Variable:* flo_prior_experience

**Do you have any prior knowledge of the Lil'Flo system?**
Select all that apply
* must provide value

- ☐ No prior knowledge
- ☐ I have read a paper on the system
- ☐ I have seen the system in person
- ☐ I have used the system
- ☐ I have some other experience with system

Add Field | Add Matrix of Fields

*Variable:* flo_interest

**How interested would you be in using the Lil'Flo system?**
* must provide value

Not At All Interested — Very Interested

Change the slider above to set a response

reset

Add Field | Add Matrix of Fields

*Variable:* flo_location

**What locations do you think Lil'Flo could be deployed in?**
Select all that apply

- ☐ Rural outpatient clinics
- ☐ Rural inpatient clinics
- ☐ Elder care facilities
- ☐ Schools
- ☐ Patient homes
- ☐ Community centers
- ☐ Urban inpatient clinics
- ☐ Urban outpatient clinics
- ☐ None
- ☐ Other

Add Field | Add Matrix of Fields

*Variable:* flo_location_other *Branching logic:* [flo_location(10)] = '1'

**What other locations?**

Expand

Add Field | Add Matrix of Fields

*Variable:* flo_desc

**How do you believe that adding a social robot as a companion for your patients during video+audio telepresence interactions (such as the LII'Flo system) would change the following when compared with traditional video+audio telepresence based rehab?**

Add Field | Add Matrix of Fields

*Variable:* flo_communication

**COMMUNICATION during the interaction**

* must provide value

Decrease communication | No change | Help communication

Change the slider above to set a response

reset

Add Field | Add Matrix of Fields

*Variable:* flo_motivation

**Patient MOTIVATION during the interaction**

* must provide value

Decrease motivation | No change | Increase motivation

Change the slider above to set a response

reset

Add Field | Add Matrix of Fields

*Variable:* flo_assessment

**Your ability to ASSESS your patients from telepresence interactions**

* must provide value

Impair assessment | Same | Improve assessment

Change the slider above to set a response

reset

Add Field | Add Matrix of Fields

*Variable:* flo_compliance

**How well your patients COMPLY with instructions DURING the telepresence interaction**

* must provide value

Reduce compliance | Same | Improve compliance

Change the slider above to set a response

reset

Add Field | Add Matrix of Fields

*Variable:* flo_adherence

**How well your patients ADHERE to the treatment plan AFTER a telepresence interaction**

* must provide value

Reduce adherence | Same | Improve adherence

Change the slider above to set a response

reset

Add Field | Add Matrix of Fields

*Variable:* telemed_sys_feat_desc

**What features do you believe are useful in a system to make tele-rehabilitation work well?**

Add Field | Add Matrix of Fields

*Matrix group:* telemed_sys_feat

| | Extremely Useless | Somewhat Useless | Neutral | Somewhat Useful | Extremely Useful |
|---|---|---|---|---|---|
| *Variable:* telemed_sys_feat_mobile<br>**A mobile system which can be driven remotely**<br>* must provide value | ○ | ○ | ○ | ○ | ○ reset |
| *Variable:* telemed_sys_feat_autodrive<br>**A mobile system which can drive on its own**<br>* must provide value | ○ | ○ | ○ | ○ | ○ reset |
| *Variable:* telemed_sys_feat_arms<br>**A social robot with arms to augment standard video calls**<br>* must provide value | ○ | ○ | ○ | ○ | ○ reset |
| *Variable:* telemed_sys_feat_face<br>**A social robot with an expressive face to augment standard video calls**<br>* must provide value | ○ | ○ | ○ | ○ | ○ reset |
| *Variable:* telemed_sys_feat_autointer<br>**A social robot that can interact with patients without needing operator input**<br>* must provide value | ○ | ○ | ○ | ○ | ○ reset |
| *Variable:* telemed_sys_feat_web<br>**A web interface for controlling remote telerehabilitation systems**<br>* must provide value | ○ | ○ | ○ | ○ | ○ reset |
| *Variable:* telemed_sys_feat_games<br>**A social robot which can play games with subjects during telerehabilitation calls to augment standard video**<br>* must provide value | ○ | ○ | ○ | ○ | ○ reset |
| *Variable:* telemed_sys_feat_asses<br>**A system which collects data and performs automated assessments of patient function**<br>* must provide value | ○ | ○ | ○ | ○ | ○ reset |
| *Variable:* telemed_sys_feat_screen<br>**A clear screen to see the clinician**<br>* must provide value | ○ | ○ | ○ | ○ | ○ reset |
| *Variable:* telemed_sys_feat_vid<br>**High quality video to see the patient**<br>* must provide value | ○ | ○ | ○ | ○ | ○ reset |

Add Field | Add Matrix of Fields

*Variable:* flo_features_other

**Please list any other features which you believe would be useful.**

Expand

Add Field | Add Matrix of Fields

**Progress:**

,

**We would like to know which tools you think are appropriate for different tasks.**

Add Field | Add Matrix of Fields

*Variable:* activities_d_desc

**Which types of activities HAVE YOU DONE with each type of tool?**

**Non-video remote communication includes: phone calls, text messages, email, instant messages, and other types of communcation which allow you to interact with patients from afar, without using video.**

Select all that apply

Add Field | Add Matrix of Fields

*Matrix group:* activities_d

| | Non-Video Remote Interaction | Video Call Based Remote Interaction | Lil'Flo Based Remote Interaction | In Person | Have Not Done |
|---|---|---|---|---|---|
| *Variable:* activities_d_motor_ass<br>**Motor Assessments**<br>* must provide value | ☐ | ☐ | ☐ | ☐ | ☐ |
| *Variable:* activities_d_stretching<br>**Stretching**<br>* must provide value | ☐ | ☐ | ☐ | ☐ | ☐ |
| *Variable:* activities_d_strength<br>**Strength Building**<br>* must provide value | ☐ | ☐ | ☐ | ☐ | ☐ |
| *Variable:* activities_d_adl<br>**ADL Practice**<br>* must provide value | ☐ | ☐ | ☐ | ☐ | ☐ |
| *Variable:* activities_d_cog_ass<br>**Cognitive Assessments**<br>* must provide value | ☐ | ☐ | ☐ | ☐ | ☐ |
| *Variable:* activites_d_cog_ex<br>**Cognitive Exercises**<br>* must provide value | ☐ | ☐ | ☐ | ☐ | ☐ |
| *Variable:* activities_d_env_adap<br>**Environmental Adaptation**<br>* must provide value | ☐ | ☐ | ☐ | ☐ | ☐ |
| *Variable:* activities_d_orthotics<br>**Orthotics Assessment/Prescription**<br>* must provide value | ☐ | ☐ | ☐ | ☐ | ☐ |
| *Variable:* activities_d_surg<br>**Discussions about Surgery**<br>* must provide value | ☐ | ☐ | ☐ | ☐ | ☐ |

*Variable:* activities_d_rads

**Discussions about Radiology Results** ☐ ☐ ☐ ☐ ☐
* must provide value

*Variable:* activities_d_med

**Medical Prescriptions** ☐ ☐ ☐ ☐ ☐
* must provide value

Add Field | Add Matrix of Fields

*Variable:* activities_b_desc

**Which types of activities DO YOU BELIEVE YOU COULD DO with each type of tool?**

**Non-video remote communication includes: phone calls, text messages, email, instant messages, and other types of communcation which allow you to interact with patients from afar, without using video.**

Select all that apply

Add Field | Add Matrix of Fields

*Matrix group:* activities_b

*Variable:* activities_b_motor_ass

| | Non-Video Remote Interaction | Video Call Based Remote Interaction | Lil'Flo Based Remote Interaction | Could Not Be Done Remotely |
|---|---|---|---|---|
| **Motor Assessments**<br>* must provide value | ☐ | ☐ | ☐ | ☐ |

*Variable:* activities_b_stretching

**Stretching** ☐ ☐ ☐ ☐
* must provide value

*Variable:* activities_b_strength

**Strength Building** ☐ ☐ ☐ ☐
* must provide value

*Variable:* activities_b_adl

**ADL Practice** ☐ ☐ ☐ ☐
* must provide value

*Variable:* activities_b_cog_ass

**Cognitive Assessments** ☐ ☐ ☐ ☐
* must provide value

*Variable:* activites_b_cog_ex

**Cognitive Exercises** ☐ ☐ ☐ ☐
* must provide value

*Variable:* activities_b_env_adap

**Environmental Adaptation** ☐ ☐ ☐ ☐
* must provide value

*Variable:* activities_b_orthotics

**Orthotics Assessment/Prescription** ☐ ☐ ☐ ☐
* must provide value

*Variable:* activities_b_surg

**Discussions about Surgery** ☐ ☐ ☐ ☐
* must provide value

*Variable:* activities_b_rads

**Discussions about Radiology Results** ☐ ☐ ☐ ☐
* must provide value

*Variable:* activities_b_med

**Medical Prescriptions** ☐ ☐ ☐ ☐
* must provide value

Add Field | Add Matrix of Fields

*Matrix group:* robot_feelings_matrix

**Progress:**

,

**In this section we ask for some final thoughts on robotics in healthcare.**

| | Strongly Disagree | Disagree | Neutral | Agree | Strongly Agree | |
|---|---|---|---|---|---|---|
| *Variable:* robot_feelings_positive<br>**I have general positive feelings towards robots**<br>* must provide value | ○ | ○ | ○ | ○ | ○ | reset |
| *Variable:* robot_feelings_same<br>**Robots are the same as any other medical/assistive devices**<br>* must provide value | ○ | ○ | ○ | ○ | ○ | reset |
| *Variable:* robot_feelings_indep<br>**Robots will improve patient independence**<br>* must provide value | ○ | ○ | ○ | ○ | ○ | reset |
| *Variable:* robot_feelings_focus<br>**Robots allow other caretakers to focus on higher level work**<br>* must provide value | ○ | ○ | ○ | ○ | ○ | reset |
| *Variable:* robot_feelings_accurate<br>**Robots are consistent and accurate**<br>* must provide value | ○ | ○ | ○ | ○ | ○ | reset |
| *Variable:* robot_feelings_expensive<br>**Robots are too expensive**<br>* must provide value | ○ | ○ | ○ | ○ | ○ | reset |
| *Variable:* robot_feelings_time<br>**Robots take too much caretaker time to set up**<br>* must provide value | ○ | ○ | ○ | ○ | ○ | reset |
| *Variable:* robot_feelings_unethical<br>**The use of robots for care and treatment is unethical**<br>* must provide value | ○ | ○ | ○ | ○ | ○ | reset |
| *Variable:* robot_feelings_patwant<br>**Patients don't want robots to take care of them**<br>* must provide value | ○ | ○ | ○ | ○ | ○ | reset |
| *Variable:* robot_feelings_control<br>**I want robots that I can control**<br>* must provide value | ○ | ○ | ○ | ○ | ○ | reset |
| *Variable:* robot_feelings_jobs<br>**Robots are going to take jobs**<br>* must provide value | ○ | ○ | ○ | ○ | ○ | reset |

*Variable:* robot_feelings_fail

**Robots are too likely to break or fail**
* must provide value

reset

*Variable:* robot_feelings_clinfeel

**Robots will improve clinician well-being**
* must provide value

reset

*Variable:* robot_feelings_neg

**I have general negative feelings towards robots**
* must provide value

reset

*Variable:* robot_feelings_failethic

**Failing to use robots for care and treatment is unethical**
* must provide value

reset

*Variable:* robot_feelings_interact

**Using robots will damage people's ability to interact with other people**
* must provide value

reset

*Variable:* robot_feelings_laws

**We need more laws and regulations about robots**
* must provide value

reset

*Variable:* robot_feelings_study

**We need to study more about how robots affect function**
* must provide value

reset

*Variable:* robot_feelings_autonomy

**I want robots that act on their own**
* must provide value

reset

*Variable:* robot_feelings_bad

**Robots are bad for society**
* must provide value

reset

Add Field | Add Matrix of Fields

*Variable:* robot_feelings_other

**What other feelings do you have towards robots?**

Expand

Add Field | Add Matrix of Fields

*Variable:* final_other

**If you have any other general thoughts you would like to share with the community about COVID-19's impacts on your practice and patients, telemedicine/telepresence, Lil'Flo, etc. Please share them here. This is the final question of the survey.**

Expand

Add Field | Add Matrix of Fields

**Progress:**

,

Add Field | Add Matrix of Fields

*Variable:* giftcard

**Would you like to be entered into the drawing for a $20 Amazon gift card?**

◯ Yes ◯ No

reset

* must provide value

Add Field | Add Matrix of Fields

*Variable:* results

**Would you like to be made aware of the results of this survey when they are published?**

◯ Yes ◯ No

reset

* must provide value

Add Field | Add Matrix of Fields

*Variable:* email *Branching logic:* [results] = '1' or [giftcard] = '1'

**Please provide your email**

* must provide value

Add Field | Add Matrix of Fields
