## Supplementary material for "Insights on Telemedicine Use by Physiatrists Before, During, and Beyond the COVID-19 Pandemic": RehabilitationAndTelemedicineT-dictionary.pdf

### Data Dictionary Codebook

02/17/2022 2:33pm

| # | Variable / Field Name | Field Label<br>*Field Note* | Field Attributes (Field Type, Validation, Choices, Calculations, etc.) |
|---|---|---|---|
| Instrument: **Clinician Survey** (clinician_survey) Enabled as survey | | | |
| 1 | [record_id] | Record ID | text |
| 2 | [occupation] | Section Header: *Progress: , This section of the survey will ask about you and your practice as it runs normally (prior to the COVID-19 pandemic).*<br>Are you: | radio, Required<br>1 A Therapist<br>2 A Nurse<br>3 A Doctor<br>4 A Medical Technologist<br>5 Other type of clinician providing rehabilitation services |
| 3 | [occupation_therapist_type]<br>Show the field ONLY if:<br>[occupation] = '1' | What type of therapist are you? | text, Required |
| 4 | [occupation_nurse_type]<br>Show the field ONLY if:<br>[occupation] = '2' | What type of nurse are you? | text, Required |
| 5 | [occupation_doctor_type]<br>Show the field ONLY if:<br>[occupation] = '3' | What type of doctor are you? | text, Required |
| 6 | [occupation_medtech_type]<br>Show the field ONLY if:<br>[occupation] = '4' | What type of medical technologist are you? | text, Required |
| 7 | [occupation_other_type]<br>Show the field ONLY if:<br>[occupation] = '5' | What type of clinician are you? | text, Required |
| 8 | [age] | What is your age (years) | text (integer), Required |
| 9 | [gender] | What is your gender? | radio, Required<br>1 Male<br>2 Female<br>3 Other<br>Field Annotation: @RANDOMORDER |
| 10 | [location] | What type of center(s) do you work in?Select all that apply | checkbox, Required<br>1 location___1 School<br>2 location___2 Hospital for Children<br>3 location___3 General Hospital<br>4 location___4 Elder Care Hospital<br>5 location___5 Rehab Center<br>6 location___6 Elder Care Home<br>7 location___7 Community Center<br>8 location___8 Private Practice<br>9 location___9 Patient Home<br>10 location___10 Inpatient Facility<br>11 location___11 Outpatient Facility<br>12 location___12 Other |
| 11 | [location_other]<br>Show the field ONLY if:<br>[location(12)] = '1' | Please specify your work location: | text, Required |

| | | | |
|---|---|---|---|
| 12 | [location_type] | Do you work in a suburban, urban, and/or rural setting?Select all that apply | checkbox, Required<br>1 location_type___1 Urban<br>2 location_type___2 Suburban<br>3 location_type___3 Rural |
| 13 | [location_country] | What country do you primarily work in? | dropdown, Required |

| | |
|---|---|
| 1 | United States of America |
| 2 | Canada |
| 3 | Mexico |
| 4 | Afghanistan |
| 5 | Albania |
| 6 | Algeria |
| 7 | Andorra |
| 8 | Angola |
| 9 | Antigua and Barbuda |
| 10 | Argentina |
| 11 | Armenia |
| 12 | Australia |
| 13 | Austria |
| 14 | Azerbaijan |
| 15 | Bahamas |
| 16 | Bahrain |
| 17 | Bangladesh |
| 18 | Barbados |
| 19 | Belarus |
| 20 | Belgium |
| 21 | Belize |
| 22 | Benin |
| 23 | Bhutan |
| 24 | Bolivia (Plurinational State of) |
| 25 | Bosnia and Herzegovina |
| 26 | Botswana |
| 27 | Brazil |
| 28 | Brunei Darussalam |
| 29 | Bulgaria |
| 30 | Burkina Faso |
| 31 | Burundi |
| 32 | Cabo Verde |
| 33 | Cambodia |
| 34 | Cameroon |
| 35 | Central African Republic |
| 36 | Chad |
| 37 | Chile |
| 38 | China |
| 39 | Colombia |
| 40 | Comoros |
| 41 | Congo |
| 42 | Costa Rica |
| 43 | Côte D'Ivoire |

| | |
|---|---|
| 44 | Croatia |
| 45 | Cuba |
| 46 | Cyprus |
| 47 | Czech Republic |
| 48 | Democratic People's Republic of Korea |
| 49 | Democratic Republic of the Congo |
| 50 | Denmark |
| 51 | Djibouti |
| 52 | Dominica |
| 53 | Dominican Republic |
| 54 | Ecuador |
| 55 | Egypt |
| 56 | El Salvador |
| 57 | Equatorial Guinea |
| 58 | Eritrea |
| 59 | Estonia |
| 60 | Eswatini (the Kingdom of) |
| 61 | Ethiopia |
| 62 | Fiji |
| 63 | Finland |
| 64 | France |
| 65 | Gabon |
| 66 | Gambia (Republic of The) |
| 67 | Georgia |
| 68 | Germany |
| 69 | Ghana |
| 70 | Greece |
| 71 | Grenada |
| 72 | Guatemala |
| 73 | Guinea |
| 74 | Guinea Bissau |
| 75 | Guyana |
| 76 | Haiti |
| 77 | Honduras |
| 78 | Hungary |
| 79 | Iceland |
| 80 | India |
| 81 | Indonesia |
| 82 | Iran (Islamic Republic of) |
| 83 | Iraq |
| 84 | Ireland |
| 85 | Israel |
| 86 | Italy |
| 87 | Jamaica |
| 88 | Japan |
| 89 | Jordan |
| 90 | Kazakhstan |
| 91 | Kenya |

| | |
|---|---|
| 92 | Kiribati |
| 93 | Kuwait |
| 94 | Kyrgyzstan |
| 95 | Lao People's Democratic Republic |
| 96 | Latvia |
| 97 | Lebanon |
| 98 | Lesotho |
| 99 | Liberia |
| 100 | Libya |
| 101 | Liechtenstein |
| 102 | Lithuania |
| 103 | Luxembourg |
| 104 | Madagascar |
| 105 | Malawi |
| 106 | Malaysia |
| 107 | Maldives |
| 108 | Mali |
| 109 | Malta |
| 110 | Marshall Islands |
| 111 | Mauritania |
| 112 | Mauritius |
| 113 | Micronesia (Federated States of) |
| 114 | Monaco |
| 115 | Mongolia |
| 116 | Montenegro |
| 117 | Morocco |
| 118 | Mozambique |
| 119 | Myanmar |
| 120 | Namibia |
| 121 | Nauru |
| 122 | Nepal |
| 123 | Netherlands |
| 124 | New Zealand |
| 125 | Nicaragua |
| 126 | Niger |
| 127 | Nigeria |
| 128 | Norway |
| 129 | Oman |
| 130 | Pakistan |
| 131 | Palau |
| 132 | Panama |
| 133 | Papua New Guinea |
| 134 | Paraguay |
| 135 | Peru |
| 136 | Philippines |
| 137 | Poland |
| 138 | Portugal |
| 139 | Qatar |

| | |
|---|---|
| 140 | Republic of Korea |
| 141 | Republic of Moldova |
| 142 | Romania |
| 143 | Russian Federation |
| 144 | Rwanda |
| 145 | Saint Kitts and Nevis |
| 146 | Saint Lucia |
| 147 | Saint Vincent and the Grenadines |
| 148 | Samoa |
| 149 | San Marino |
| 150 | Sao Tome and Principe |
| 151 | Saudi Arabia |
| 152 | Senegal |
| 153 | Serbia |
| 154 | Seychelles |
| 155 | Sierra Leone |
| 156 | Singapore |
| 157 | Slovakia |
| 158 | Slovenia |
| 159 | Solomon Islands |
| 160 | Somalia |
| 161 | South Africa |
| 162 | South Sudan |
| 163 | Spain |
| 164 | Sri Lanka |
| 165 | Sudan |
| 166 | Suriname |
| 167 | Sweden |
| 168 | Switzerland |
| 169 | Syrian Arab Republic |
| 170 | Tajikistan |
| 171 | Thailand |
| 172 | The former Yugoslav Republic of Macedonia |
| 173 | Timor-Leste |
| 174 | Togo |
| 175 | Tonga |
| 176 | Trinidad and Tobago |
| 177 | Tunisia |
| 178 | Turkey |
| 179 | Turkmenistan |
| 180 | Tuvalu |
| 181 | Uganda |
| 182 | Ukraine |
| 183 | United Arab Emirates |
| 184 | United Kingdom of Great Britain and Northern Ireland |
| 185 | United Republic of Tanzania |
| 186 | Uruguay |

| | | | |
|---|---|---|---|
| | | | 187 Uzbekistan<br>188 Vanuatu<br>Venezuela Bolivarian Republic of<br>189 Viet Nam<br>190 Yemen<br>191 Zambia<br>192 Zimbabwe |
| 14 | [ location_state ]<br>Show the field ONLY if:<br>[location_country] = '1' | What state do you primarily work in? | dropdown, Required<br>1 Alabama<br>2 Alaska<br>3 Arizona<br>4 Arkansas<br>5 California<br>6 Colorado<br>7 Connecticut<br>8 Delaware<br>9 Florida<br>10 Georgia<br>11 Hawaii<br>12 Idaho<br>13 Illinois<br>14 Indiana<br>15 Iowa<br>16 Kansas<br>17 Kentucky<br>18 Louisiana<br>19 Maine<br>20 Maryland<br>21 Massachusetts<br>22 Michigan<br>23 Minnesota<br>24 Mississippi<br>25 Missouri<br>26 Montana<br>27 Nebraska<br>28 Nevada<br>29 New Hampshire<br>30 New Jersey<br>31 New Mexico<br>32 New York<br>33 North Carolina<br>34 North Dakota<br>35 Ohio<br>36 Oklahoma<br>37 Oregon<br>38 Pennsylvania<br>39 Rhode Island<br>40 South Carolina |

| | | | |
|---|---|---|---|
| | | | 41 South Dakota<br>42 Tennessee<br>43 Texas<br>44 Utah<br>45 Vermont<br>46 Virginia<br>47 Washington<br>48 West Virginia<br>49 Wisconsin<br>50 Wyoming<br>51 Washington, D.C. |
| 15 | [years_experience] | How many years have you been practicing? | text (integer), Required |
| 16 | [patient_pop] | Please select descriptors for the type of patients you treat. Patients with..Select all that apply | checkbox, Required<br>1 patient_pop___1 CP<br>2 patient_pop___2 Stroke<br>3 patient_pop___3 Brachial Plexus Injuries<br>4 patient_pop___4 Autism<br>5 patient_pop___5 TBI<br>6 patient_pop___6 Cardiovascular Disease<br>7 patient_pop___7 Speech Disorders<br>8 patient_pop___8 Sports Injuries<br>9 patient_pop___9 Amputations<br>10 patient_pop___10 Arthritis<br>11 patient_pop___11 Dementia<br>12 patient_pop___12 Burns<br>13 patient_pop___13 General Traumatic Injuries<br>15 patient_pop___15 Mental Health Issues<br>14 patient_pop___14 Other |
| 17 | [patient_pop_other]<br>Show the field ONLY if: [patient_pop(14)] = '1' | Please describe the other patient types who you treat: | notes |
| 18 | [patient_pop_cog] | What are the levels of COGNITIVE impairment that your patients have?Select all that apply | checkbox, Required<br>1 patient_pop_cog___1 Severely Impaired<br>2 patient_pop_cog___2 Moderately Impaired<br>3 patient_pop_cog___3 Mildly Impaired<br>4 patient_pop_cog___4 Not Impaired |
| 19 | [patient_pop_motor] | What are the levels of MOTOR impairment that your patients have?Select all that apply | checkbox, Required<br>1 patient_pop_motor___1 Severely Impaired<br>2 patient_pop_motor___2 Moderately Impaired<br>3 patient_pop_motor___3 Mildly Impaired<br>4 patient_pop_motor___4 Not Impaired |

| | | | |
|---|---|---|---|
| 20 | [ patient_pop_age ] | What age groups do you generally work with?Select all that apply | checkbox, Required<br>1 \| patient_pop_age___1 \| Infants (0-1)<br>2 \| patient_pop_age___2 \| Toddlers (1-3)<br>3 \| patient_pop_age___3 \| Preschoolers (3-5)<br>4 \| patient_pop_age___4 \| Gradeschoolers (5-12)<br>5 \| patient_pop_age___5 \| Teens (12-18)<br>6 \| patient_pop_age___6 \| Young Adults (18-21)<br>7 \| patient_pop_age___7 \| Adults (21-40)<br>8 \| patient_pop_age___8 \| Middle Aged Adults (40-65)<br>9 \| patient_pop_age___9 \| Older Adults (65+) |
| 21 | [ patient_pop_phase ] | What phase of care are your patients in?Select all that apply | checkbox, Required<br>1 \| patient_pop_phase___1 \| Acute<br>2 \| patient_pop_phase___2 \| Sub-Acute<br>3 \| patient_pop_phase___3 \| Chronic |
| 22 | [ past_telepresence ] | Do you use any of these tools to interact with patients during normal practice (prior to the COVID-19 pandemic)? Select all that apply | checkbox, Required<br>1 \| past_telepresence___1 \| Phone calls with patients<br>2 \| past_telepresence___2 \| Text based communication (email, text message, text app, etc.)<br>3 \| past_telepresence___3 \| Video calls to patients at home<br>4 \| past_telepresence___4 \| Video calls to patients in another clinical setting<br>5 \| past_telepresence___5 \| Other method of remotely connecting with patients<br>6 \| past_telepresence___6 \| None of these |
| 23 | [ past_telepresence_other ]<br>Show the field ONLY if:<br>[past_telepresence(5)] = '1' | What is the other method you have used? | notes<br>Custom alignment: RH |
| 24 | [ covid_inperson ] | Section Header: *Progress: 20% In this section we will ask you about your experience delivering care to your patients as a result of the COVID-19 pandemic.*<br>Have you been able to see your patients in-person during the pandemic? | yesno, Required<br>1 \| Yes<br>0 \| No<br>Custom alignment: RH |
| 25 | [ covid_treatment_difficulty ] | How has the pandemic changed your level of interaction with your patients? | slider, Required<br>Slider labels: I cannot interact with my patients, I am interacting with my patients as normal, I am interacting with my patients much more than normal<br>Custom alignment: RH |
| 26 | [ covid_outcomes ] | How do you expect the pandemic to affect your patients' long term health outcomes? | slider, Required<br>Slider labels: Much worse outcomes, Unchanged outcomes, Much better outcomes<br>Custom alignment: RH |
| 27 | [ covid_patient_selfcare ] | How do you believe the pandemic is affecting your patients' ability to care for themselves and meet their rehabilitation goals? | slider, Required<br>Slider labels: Much worse, Unchanged, Much better<br>Custom alignment: RH |

| | | | |
|---|---|---|---|
| 28 | [ covid_telepresence ] | Are you using any of these tools to connect with your patients during the COVID-19 pandemic?Select all that apply | checkbox, Required<br>1 \| covid_telepresence___1 \| Phone calls with patients<br>2 \| covid_telepresence___2 \| Text based communication (email, text message, text app, etc.)<br>3 \| covid_telepresence___3 \| Video calls to patients at home<br>4 \| covid_telepresence___4 \| Video calls to patients in another clinical setting<br>5 \| covid_telepresence___5 \| Other method of remotely connecting with patients<br>6 \| covid_telepresence___6 \| None of these |
| 29 | [ covid_telepresence_other ]<br>Show the field ONLY if:<br>[covid_telepresence(5)] = '1' | What is the other method you are using? | notes |
| 30 | [ video_plan ]<br>Show the field ONLY if:<br>[covid_telepresence(3)] = '0' and [covid_telepresence(4)] = '0' | Do you plan to use video calls to talk to your patients going forward during the COVID-19 pandemic? | yesno, Required<br>1 \| Yes<br>0 \| No |
| 31 | [ video_satisfaction ]<br>Show the field ONLY if:<br>[past_telepresence(3)] = '1' or [past_telepresence(4)] = '1' or [covid_telepresence(3)] = '1' or [covid_telepresence(4)] = '1' | Section Header: *Progress: , In this section we will ask you about your experience and thoughts about using telemedicine, specifically video calls, both before and during the COVID-19 pandemic.*<br>Please rate your overall SATISFACTION using video calls for rehab | slider, Required<br>Slider labels: Very Dissatisfied, Neutral, Very Satisfied<br>Custom alignment: RH |
| 32 | [ video_communication ]<br>Show the field ONLY if:<br>[past_telepresence(3)] = '1' or [past_telepresence(4)] = '1' or [covid_telepresence(3)] = '1' or [covid_telepresence(4)] = '1' | How well are you able to COMMUNICATE with your patients over video calls compared to in-person? | slider, Required<br>Slider labels: In-person much better communication, No difference, Video call much better communication<br>Custom alignment: RH |
| 33 | [ pvideo_communication ]<br>Show the field ONLY if:<br>[past_telepresence(3)] = '0' and [past_telepresence(4)] = '0' and [covid_telepresence(3)] = '0' and [covid_telepresence(4)] = '0' | How well do you believe you could COMMUNICATE with your patients over video calls compared to in-person? | slider, Required<br>Slider labels: In-person much better communication, No difference, Video call much better communication<br>Custom alignment: RH |
| 34 | [ video_assessment ]<br>Show the field ONLY if:<br>[past_telepresence(3)] = '1' or [past_telepresence(4)] = '1' or [covid_telepresence(3)] = '1' or [covid_telepresence(4)] = '1' | How well are you able to ASSESS your patients' level of function over video calls compared to in-person? | slider, Required<br>Slider labels: In-person much better assessment, No difference, Video call much better assessment<br>Custom alignment: RH |
| 35 | [ pvideo_assessment ]<br>Show the field ONLY if:<br>[past_telepresence(3)] = '0' and [past_telepresence(4)] = '0' and [covid_telepresence(3)] = '0' and [covid_telepresence(4)] = '0' | How well do you believe you could ASSESS your patients' level of function over video calls compared to in-person? | slider, Required<br>Slider labels: In-person much better assessment, No difference, Video call much better assessment<br>Custom alignment: RH |

| | | | |
|---|---|---|---|
| 36 | [video_motivation]<br>Show the field ONLY if:<br>[past_telepresence(3)] = '1' or [past_telepresence(4)] = '1' or [covid_telepresence(3)] = '1' or [covid_telepresence(4)] = '1' | How MOTIVATED are your patients DURING a video call compared to in-person? | slider, Required<br>Slider labels: In-person much more motivated, No difference, Video call much more motivated<br>Custom alignment: RH |
| 37 | [pvideo_motivation]<br>Show the field ONLY if:<br>[past_telepresence(3)] = '0' and [past_telepresence(4)] = '0' and [covid_telepresence(3)] = '0' and [covid_telepresence(4)] = '0' | How MOTIVATED do you believe your patients would be DURING a video call compared to in-person? | slider, Required<br>Slider labels: In-person much more motivated, No difference, Video call much more motivated<br>Custom alignment: RH |
| 38 | [video_compliance_during]<br>Show the field ONLY if:<br>[past_telepresence(3)] = '1' or [past_telepresence(4)] = '1' or [covid_telepresence(3)] = '1' or [covid_telepresence(4)] = '1' | How well do your patients COMPLY with instructions DURING a video call vs in-person visit? | slider, Required<br>Slider labels: In-person much higher compliance, The same, Video call much higher compliance<br>Custom alignment: RH |
| 39 | [pvideo_compliance_during]<br>Show the field ONLY if:<br>[past_telepresence(3)] = '0' and [past_telepresence(4)] = '0' and [covid_telepresence(3)] = '0' and [covid_telepresence(4)] = '0' | How well do you believe your patients would COMPLY with instructions DURING a video call vs in-person visit? | slider, Required<br>Slider labels: In-person much higher compliance, The same, Video call much higher compliance<br>Custom alignment: RH |
| 40 | [video_adherence_after]<br>Show the field ONLY if:<br>[past_telepresence(3)] = '1' or [past_telepresence(4)] = '1' or [covid_telepresence(3)] = '1' or [covid_telepresence(4)] = '1' | How well do your patients ADHERE to the treatment plan AFTER a video call vs in-person visit? | slider, Required<br>Slider labels: In-person much higher adherence, The same, Video call much higher adherence<br>Custom alignment: RH |
| 41 | [pvideo_adherence_after]<br>Show the field ONLY if:<br>[past_telepresence(3)] = '0' and [past_telepresence(4)] = '0' and [covid_telepresence(3)] = '0' and [covid_telepresence(4)] = '0' | How well do you believe your patients would ADHERE to the treatment plan AFTER a video call vs in-person visit? | slider, Required<br>Slider labels: In-person much higher compliance, The same, Video call much higher compliance<br>Custom alignment: RH |

| | | | |
|---|---|---|---|
| 42 | [ video_challenges ]<br>Show the field ONLY if:<br>[past_telepresence(3)] = '1' or [past_telepresence(4)] = '1' or [covid_telepresence(3)] = '1' or [covid_telepresence(4)] = '1' | Do you face any challenges using video calls that affect quality of care?Select all that apply | checkbox |

| | | |
|---|---|---|
| 1 | video_challenges___1 | No challenges |
| 2 | video_challenges___2 | connection failures |
| 3 | video_challenges___3 | poor audio quality |
| 4 | video_challenges___4 | poor video quality |
| 5 | video_challenges___5 | Patients do not have a device for video |
| 6 | video_challenges___6 | Patients do not have internet |
| 7 | video_challenges___7 | Patients cannot understand what I want |
| 8 | video_challenges___8 | I cannot understand what patients want |
| 9 | video_challenges___9 | Patients cannot complete activities without my physical intervention |
| 10 | video_challenges___10 | Assessments require the use of a tool which the patient does not have |
| 11 | video_challenges___11 | It is not safe for patients to do the desired activities without a professional present |
| 12 | video_challenges___12 | Other challenges |

| | | | |
|---|---|---|---|
| 43 | [ pvideo_challenges ]<br>Show the field ONLY if:<br>[past_telepresence(3)] = '0' and [past_telepresence(4)] = '0' and [covid_telepresence(3)] = '0' and [covid_telepresence(4)] = '0' | Do you believe you would face any challenges using video calls that would affect quality of care?Select all that apply | checkbox |

| | | |
|---|---|---|
| 1 | pvideo_challenges___1 | No challenges |
| 2 | pvideo_challenges___2 | Connection failures |
| 3 | pvideo_challenges___3 | Poor audio quality |
| 4 | pvideo_challenges___4 | Poor video quality |
| 5 | pvideo_challenges___5 | Patients would not have a device for video |
| 6 | pvideo_challenges___6 | Patients would not have internet |
| 7 | pvideo_challenges___7 | Patients would not understand what I want |
| 8 | pvideo_challenges___8 | I would not understand what patients want |
| 9 | pvideo_challenges___9 | Patients could not complete activities without my physical intervention |
| 10 | pvideo_challenges___10 | Assessments require the use of a tool which the patient would not have |
| 11 | pvideo_challenges___11 | It would not be safe for patients to do the desired activities without a professional present |
| 12 | pvideo_challenges___12 | Other challenges |

| | | | |
|---|---|---|---|
| 44 | [video_challenges_other]<br>Show the field ONLY if:<br>[video_challenges(12)] = '1' or [pvideo_challenges(12)] = '1' | What other challenges? | notes<br>Custom alignment: RH |
| 45 | [video_future] | Do you plan to use video calls with your patients for your practice after the COVID-19 pandemic has ended? | radio, Required<br>1 Yes<br>2 No<br>3 Not Sure |
| 46 | [flo_description] | Section Header: *Progress: , In this section we present a new robotic platform for telemedicine, specifically telerehabilitation. We then ask some questions about the system and how it could fit into your practice.*<br>Please watch this video which describes the Lil'Flo Robot, a socially assitive telerehabilitation robot.<br><br>The system consists of a social robot (a robot which interacts with people socially), which can play games, demonstrate motions, and express emotion, mounted on a mobile telepresence system which has a screen, cameras, and a microphone.<br><br>Then continue to answer questions about the system. | descriptive |
| 47 | [flo_prior_experience] | Do you have any prior knowledge of the Lil'Flo system?Select all that apply | checkbox, Required<br>1 flo_prior_experience___1 No prior knowledge<br>2 flo_prior_experience___2 I have read a paper on the system<br>3 flo_prior_experience___3 I have seen the system in person<br>4 flo_prior_experience___4 I have used the system<br>5 flo_prior_experience___5 I have some other experience with system |
| 48 | [flo_interest] | How interested would you be in using the Lil'Flo system? | slider, Required<br>Slider labels: Not At All Interested, , Very Interested<br>Custom alignment: RH |
| 49 | [flo_location] | What locations do you think Lil'Flo could be deployed in?Select all that apply | checkbox<br>1 flo_location___1 Rural outpatient clinics<br>2 flo_location___2 Rural inpatient clinics<br>3 flo_location___3 Elder care facilities<br>4 flo_location___4 Schools<br>5 flo_location___5 Patient homes<br>6 flo_location___6 Community centers<br>7 flo_location___7 Urban inpatient clinics<br>8 flo_location___8 Urban outpatient clinics<br>9 flo_location___9 None<br>10 flo_location___10 Other |
| 50 | [flo_location_other]<br>Show the field ONLY if:<br>[flo_location(10)] = '1' | What other locations? | notes |
| 51 | [flo_desc] | How do you believe that adding a social robot as a companion for your patients during video+audio telepresence interactions (such as the LIl'Flo system) would change the following when compared with traditional video+audio telepresence based rehab? | descriptive |
| 52 | [flo_communication] | COMMUNICATION during the interaction | slider, Required<br>Slider labels: Decrease communication, No change, Help communication<br>Custom alignment: RH |

| | | | |
|---|---|---|---|
| 53 | [flo_motivation] | Patient MOTIVATION during the interaction | slider, Required<br>Slider labels: Decrease motivation, No change, Increase motivation<br>Custom alignment: RH |
| 54 | [flo_assessment] | Your ability to ASSESS your patients from telepresence interactions | slider, Required<br>Slider labels: Impair assessment, Same, Improve assessment<br>Custom alignment: RH |
| 55 | [flo_compliance] | How well your patients COMPLY with instructions DURING the telepresence interaction | slider, Required<br>Slider labels: Reduce compliance, Same, Improve compliance<br>Custom alignment: RH |
| 56 | [flo_adherence] | How well your patients ADHERE to the treatment plan AFTER a telepresence interaction | slider, Required<br>Slider labels: Reduce adherence, Same, Improve adherence<br>Custom alignment: RH |
| 57 | [telemed_sys_feat_desc] | What features do you believe are useful in a system to make tele-rehabilitation work well? | descriptive |
| 58 | [telemed_sys_feat_mobile] | A mobile system which can be driven remotely | radio (Matrix), Required<br>1 Extremely Useless<br>2 Somewhat Useless<br>3 Neutral<br>4 Somewhat Useful<br>5 Extremely Useful |
| 59 | [telemed_sys_feat_autodrive] | A mobile system which can drive on its own | radio (Matrix), Required<br>1 Extremely Useless<br>2 Somewhat Useless<br>3 Neutral<br>4 Somewhat Useful<br>5 Extremely Useful |
| 60 | [telemed_sys_feat_arms] | A social robot with arms to augment standard video calls | radio (Matrix), Required<br>1 Extremely Useless<br>2 Somewhat Useless<br>3 Neutral<br>4 Somewhat Useful<br>5 Extremely Useful |
| 61 | [telemed_sys_feat_face] | A social robot with an expressive face to augment standard video calls | radio (Matrix), Required<br>1 Extremely Useless<br>2 Somewhat Useless<br>3 Neutral<br>4 Somewhat Useful<br>5 Extremely Useful |
| 62 | [telemed_sys_feat_autointer] | A social robot that can interact with patients without needing operator input | radio (Matrix), Required<br>1 Extremely Useless<br>2 Somewhat Useless<br>3 Neutral<br>4 Somewhat Useful<br>5 Extremely Useful |

| | | | |
|---|---|---|---|
| 63 | [telemed_sys_feat_web] | A web interface for controlling remote telerehabilitation systems | radio (Matrix), Required<br>1 Extremely Useless<br>2 Somewhat Useless<br>3 Neutral<br>4 Somewhat Useful<br>5 Extremely Useful |
| 64 | [telemed_sys_feat_games] | A social robot which can play games with subjects during telerehabilitation calls to augment standard video | radio (Matrix), Required<br>1 Extremely Useless<br>2 Somewhat Useless<br>3 Neutral<br>4 Somewhat Useful<br>5 Extremely Useful |
| 65 | [telemed_sys_feat_asses] | A system which collects data and performs automated assessments of patient function | radio (Matrix), Required<br>1 Extremely Useless<br>2 Somewhat Useless<br>3 Neutral<br>4 Somewhat Useful<br>5 Extremely Useful |
| 66 | [telemed_sys_feat_screen] | A clear screen to see the clinician | radio (Matrix), Required<br>1 Extremely Useless<br>2 Somewhat Useless<br>3 Neutral<br>4 Somewhat Useful<br>5 Extremely Useful |
| 67 | [telemed_sys_feat_vid] | High quality video to see the patient | radio (Matrix), Required<br>1 Extremely Useless<br>2 Somewhat Useless<br>3 Neutral<br>4 Somewhat Useful<br>5 Extremely Useful |
| 68 | [flo_features_other] | Please list any other features which you believe would be useful. | notes |
| 69 | [activities_d_desc] | Section Header: *Progress: , We would like to know which tools you think are appropriate for different tasks.*<br>Which types of activities HAVE YOU DONE with each type of tool? Non-video remote communication includes: phone calls, text messages, email, instant messages, and other types of communcation which allow you to interact with patients from afar, without using video. Select all that apply | descriptive |
| 70 | [activities_d_motor_ass] | Motor Assessments | checkbox, Required<br>1 activities_d_motor_ass___1 Non-Video Remote Interaction<br>3 activities_d_motor_ass___3 Video Call Based Remote Interaction<br>4 activities_d_motor_ass___4 Lil'Flo Based Remote Interaction<br>5 activities_d_motor_ass___5 In Person<br>6 activities_d_motor_ass___6 Have Not Done |

| | | | |
|---|---|---|---|
| 71 | [activities_d_stretching] | Stretching | checkbox, Required<br>1 activities_d_stretching___1 Non-Video Remote Interaction<br>3 activities_d_stretching___3 Video Call Based Remote Interaction<br>4 activities_d_stretching___4 Lil'Flo Based Remote Interaction<br>5 activities_d_stretching___5 In Person<br>6 activities_d_stretching___6 Have Not Done |
| 72 | [activities_d_strength] | Strength Building | checkbox, Required<br>1 activities_d_strength___1 Non-Video Remote Interaction<br>3 activities_d_strength___3 Video Call Based Remote Interaction<br>4 activities_d_strength___4 Lil'Flo Based Remote Interaction<br>5 activities_d_strength___5 In Person<br>6 activities_d_strength___6 Have Not Done |
| 73 | [activities_d_adl] | ADL Practice | checkbox, Required<br>1 activities_d_adl___1 Non-Video Remote Interaction<br>3 activities_d_adl___3 Video Call Based Remote Interaction<br>4 activities_d_adl___4 Lil'Flo Based Remote Interaction<br>5 activities_d_adl___5 In Person<br>6 activities_d_adl___6 Have Not Done |
| 74 | [activities_d_cog_ass] | Cognitive Assessments | checkbox, Required<br>1 activities_d_cog_ass___1 Non-Video Remote Interaction<br>3 activities_d_cog_ass___3 Video Call Based Remote Interaction<br>4 activities_d_cog_ass___4 Lil'Flo Based Remote Interaction<br>5 activities_d_cog_ass___5 In Person<br>6 activities_d_cog_ass___6 Have Not Done |
| 75 | [activites_d_cog_ex] | Cognitive Exercises | checkbox, Required<br>1 activites_d_cog_ex___1 Non-Video Remote Interaction<br>3 activites_d_cog_ex___3 Video Call Based Remote Interaction<br>4 activites_d_cog_ex___4 Lil'Flo Based Remote Interaction<br>5 activites_d_cog_ex___5 In Person<br>6 activites_d_cog_ex___6 Have Not Done |
| 76 | [activities_d_env_adap] | Environmental Adaptation | checkbox, Required<br>1 activities_d_env_adap___1 Non-Video Remote Interaction<br>3 activities_d_env_adap___3 Video Call Based Remote Interaction<br>4 activities_d_env_adap___4 Lil'Flo Based Remote Interaction<br>5 activities_d_env_adap___5 In Person<br>6 activities_d_env_adap___6 Have Not Done |

| | | | |
|---|---|---|---|
| 77 | [activities_d_orthotics] | Orthotics Assessment/Prescription | checkbox, Required<br>1 activities_d_orthotics___1 Non-Video Remote Interaction<br>3 activities_d_orthotics___3 Video Call Based Remote Interaction<br>4 activities_d_orthotics___4 Lil'Flo Based Remote Interaction<br>5 activities_d_orthotics___5 In Person<br>6 activities_d_orthotics___6 Have Not Done |
| 78 | [activities_d_surg] | Discussions about Surgery | checkbox, Required<br>1 activities_d_surg___1 Non-Video Remote Interaction<br>3 activities_d_surg___3 Video Call Based Remote Interaction<br>4 activities_d_surg___4 Lil'Flo Based Remote Interaction<br>5 activities_d_surg___5 In Person<br>6 activities_d_surg___6 Have Not Done |
| 79 | [activities_d_rads] | Discussions about Radiology Results | checkbox, Required<br>1 activities_d_rads___1 Non-Video Remote Interaction<br>3 activities_d_rads___3 Video Call Based Remote Interaction<br>4 activities_d_rads___4 Lil'Flo Based Remote Interaction<br>5 activities_d_rads___5 In Person<br>6 activities_d_rads___6 Have Not Done |
| 80 | [activities_d_med] | Medical Prescriptions | checkbox, Required<br>1 activities_d_med___1 Non-Video Remote Interaction<br>3 activities_d_med___3 Video Call Based Remote Interaction<br>4 activities_d_med___4 Lil'Flo Based Remote Interaction<br>5 activities_d_med___5 In Person<br>6 activities_d_med___6 Have Not Done |
| 81 | [activities_b_desc] | Which types of activities DO YOU BELIEVE YOU COULD DO with each type of tool?Non-video remote communication includes: phone calls, text messages, email, instant messages, and other types of communcation which allow you to interact with patients from afar, without using video.Select all that apply | descriptive |
| 82 | [activities_b_motor_ass] | Motor Assessments | checkbox, Required<br>1 activities_b_motor_ass___1 Non-Video Remote Interaction<br>3 activities_b_motor_ass___3 Video Call Based Remote Interaction<br>4 activities_b_motor_ass___4 Lil'Flo Based Remote Interaction<br>5 activities_b_motor_ass___5 Could Not Be Done Remotely |

| | | | |
|---|---|---|---|
| 83 | [activities_b_stretching] | Stretching | checkbox, Required<br>1 activities_b_stretching___1 Non-Video Remote Interaction<br>3 activities_b_stretching___3 Video Call Based Remote Interaction<br>4 activities_b_stretching___4 Lil'Flo Based Remote Interaction<br>5 activities_b_stretching___5 Could Not Be Done Remotely |
| 84 | [activities_b_strength] | Strength Building | checkbox, Required<br>1 activities_b_strength___1 Non-Video Remote Interaction<br>3 activities_b_strength___3 Video Call Based Remote Interaction<br>4 activities_b_strength___4 Lil'Flo Based Remote Interaction<br>5 activities_b_strength___5 Could Not Be Done Remotely |
| 85 | [activities_b_adl] | ADL Practice | checkbox, Required<br>1 activities_b_adl___1 Non-Video Remote Interaction<br>3 activities_b_adl___3 Video Call Based Remote Interaction<br>4 activities_b_adl___4 Lil'Flo Based Remote Interaction<br>5 activities_b_adl___5 Could Not Be Done Remotely |
| 86 | [activities_b_cog_ass] | Cognitive Assessments | checkbox, Required<br>1 activities_b_cog_ass___1 Non-Video Remote Interaction<br>3 activities_b_cog_ass___3 Video Call Based Remote Interaction<br>4 activities_b_cog_ass___4 Lil'Flo Based Remote Interaction<br>5 activities_b_cog_ass___5 Could Not Be Done Remotely |
| 87 | [activites_b_cog_ex] | Cognitive Exercises | checkbox, Required<br>1 activites_b_cog_ex___1 Non-Video Remote Interaction<br>3 activites_b_cog_ex___3 Video Call Based Remote Interaction<br>4 activites_b_cog_ex___4 Lil'Flo Based Remote Interaction<br>5 activites_b_cog_ex___5 Could Not Be Done Remotely |
| 88 | [activities_b_env_adap] | Environmental Adaptation | checkbox, Required<br>1 activities_b_env_adap___1 Non-Video Remote Interaction<br>3 activities_b_env_adap___3 Video Call Based Remote Interaction<br>4 activities_b_env_adap___4 Lil'Flo Based Remote Interaction<br>5 activities_b_env_adap___5 Could Not Be Done Remotely |

| | | | |
|---|---|---|---|
| 89 | [activities_b_orthotics] | Orthotics Assessment/Prescription | checkbox, Required<br>1 activities_b_orthotics___1 Non-Video Remote Interaction<br>3 activities_b_orthotics___3 Video Call Based Remote Interaction<br>4 activities_b_orthotics___4 Lil'Flo Based Remote Interaction<br>5 activities_b_orthotics___5 Could Not Be Done Remotely |
| 90 | [activities_b_surg] | Discussions about Surgery | checkbox, Required<br>1 activities_b_surg___1 Non-Video Remote Interaction<br>3 activities_b_surg___3 Video Call Based Remote Interaction<br>4 activities_b_surg___4 Lil'Flo Based Remote Interaction<br>5 activities_b_surg___5 Could Not Be Done Remotely |
| 91 | [activities_b_rads] | Discussions about Radiology Results | checkbox, Required<br>1 activities_b_rads___1 Non-Video Remote Interaction<br>3 activities_b_rads___3 Video Call Based Remote Interaction<br>4 activities_b_rads___4 Lil'Flo Based Remote Interaction<br>5 activities_b_rads___5 Could Not Be Done Remotely |
| 92 | [activities_b_med] | Medical Prescriptions | checkbox, Required<br>1 activities_b_med___1 Non-Video Remote Interaction<br>3 activities_b_med___3 Video Call Based Remote Interaction<br>4 activities_b_med___4 Lil'Flo Based Remote Interaction<br>5 activities_b_med___5 Could Not Be Done Remotely |
| 93 | [robot_feelings_positive] | Section Header: *Progress: , In this section we ask for some final thoughts on robotics in healthcare.*<br>I have general positive feelings towards robots | radio (Matrix), Required<br>1 Strongly Disagree<br>2 Disagree<br>3 Neutral<br>4 Agree<br>5 Strongly Agree |
| 94 | [robot_feelings_same] | Robots are the same as any other medical/assistive devices | radio (Matrix), Required<br>1 Strongly Disagree<br>2 Disagree<br>3 Neutral<br>4 Agree<br>5 Strongly Agree |

| | | | |
|---|---|---|---|
| 95 | [robot_feelings_indep] | Robots will improve patient independence | radio (Matrix), Required<br>1 Strongly Disagree<br>2 Disagree<br>3 Neutral<br>4 Agree<br>5 Strongly Agree |
| 96 | [robot_feelings_focus] | Robots allow other caretakers to focus on higher level work | radio (Matrix), Required<br>1 Strongly Disagree<br>2 Disagree<br>3 Neutral<br>4 Agree<br>5 Strongly Agree |
| 97 | [robot_feelings_accurate] | Robots are consistent and accurate | radio (Matrix), Required<br>1 Strongly Disagree<br>2 Disagree<br>3 Neutral<br>4 Agree<br>5 Strongly Agree |
| 98 | [robot_feelings_expensive] | Robots are too expensive | radio (Matrix), Required<br>1 Strongly Disagree<br>2 Disagree<br>3 Neutral<br>4 Agree<br>5 Strongly Agree |
| 99 | [robot_feelings_time] | Robots take too much caretaker time to set up | radio (Matrix), Required<br>1 Strongly Disagree<br>2 Disagree<br>3 Neutral<br>4 Agree<br>5 Strongly Agree |
| 100 | [robot_feelings_unethical] | The use of robots for care and treatment is unethical | radio (Matrix), Required<br>1 Strongly Disagree<br>2 Disagree<br>3 Neutral<br>4 Agree<br>5 Strongly Agree |
| 101 | [robot_feelings_patwant] | Patients don't want robots to take care of them | radio (Matrix), Required<br>1 Strongly Disagree<br>2 Disagree<br>3 Neutral<br>4 Agree<br>5 Strongly Agree |
| 102 | [robot_feelings_control] | I want robots that I can control | radio (Matrix), Required<br>1 Strongly Disagree<br>2 Disagree<br>3 Neutral<br>4 Agree<br>5 Strongly Agree |

| | | | |
|---|---|---|---|
| 103 | [ robot_feelings_jobs ] | Robots are going to take jobs | radio (Matrix), Required<br>1 Strongly Disagree<br>2 Disagree<br>3 Neutral<br>4 Agree<br>5 Strongly Agree |
| 104 | [ robot_feelings_fail ] | Robots are too likely to break or fail | radio (Matrix), Required<br>1 Strongly Disagree<br>2 Disagree<br>3 Neutral<br>4 Agree<br>5 Strongly Agree |
| 105 | [ robot_feelings_clinfeel ] | Robots will improve clinician well-being | radio (Matrix), Required<br>1 Strongly Disagree<br>2 Disagree<br>3 Neutral<br>4 Agree<br>5 Strongly Agree |
| 106 | [ robot_feelings_neg ] | I have general negative feelings towards robots | radio (Matrix), Required<br>1 Strongly Disagree<br>2 Disagree<br>3 Neutral<br>4 Agree<br>5 Strongly Agree |
| 107 | [ robot_feelings_failethic ] | Failing to use robots for care and treatment is unethical | radio (Matrix), Required<br>1 Strongly Disagree<br>2 Disagree<br>3 Neutral<br>4 Agree<br>5 Strongly Agree |
| 108 | [ robot_feelings_interact ] | Using robots will damage people's ability to interact with other people | radio (Matrix), Required<br>1 Strongly Disagree<br>2 Disagree<br>3 Neutral<br>4 Agree<br>5 Strongly Agree |
| 109 | [ robot_feelings_laws ] | We need more laws and regulations about robots | radio (Matrix), Required<br>1 Strongly Disagree<br>2 Disagree<br>3 Neutral<br>4 Agree<br>5 Strongly Agree |
| 110 | [ robot_feelings_study ] | We need to study more about how robots affect function | radio (Matrix), Required<br>1 Strongly Disagree<br>2 Disagree<br>3 Neutral<br>4 Agree<br>5 Strongly Agree |

| | | | |
|---|---|---|---|
| 111 | [robot_feelings_autonomy] | I want robots that act on their own | radio (Matrix), Required<br>1 Strongly Disagree<br>2 Disagree<br>3 Neutral<br>4 Agree<br>5 Strongly Agree |
| 112 | [robot_feelings_bad] | Robots are bad for society | radio (Matrix), Required<br>1 Strongly Disagree<br>2 Disagree<br>3 Neutral<br>4 Agree<br>5 Strongly Agree |
| 113 | [robot_feelings_other] | What other feelings do you have towards robots? | notes |
| 114 | [final_other] | If you have any other general thoughts you would like to share with the community about COVID-19's impacts on your practice and patients, telemedicine/telepresence, Lil'Flo, etc. Please share them here. This is the final question of the survey. | notes |
| 115 | [giftcard] | Section Header: *Progress: ,*<br>Would you like to be entered into the drawing for a $20 Amazon gift card? | yesno, Required<br>1 Yes<br>0 No<br>Custom alignment: RH |
| 116 | [results] | Would you like to be made aware of the results of this survey when they are published? | yesno, Required<br>1 Yes<br>0 No<br>Custom alignment: RH |
| 117 | [email]<br>Show the field ONLY if:<br>[results] = '1' or [giftcard] = '1' | Please provide your email | text (email), Required, Identifier<br>Custom alignment: RH |
| 118 | [clinician_survey_complete] | Section Header: *Form Status*<br>Complete? | dropdown<br>0 Incomplete<br>1 Unverified<br>2 Complete |
