## Supplementary material for "Insights on Telemedicine Use by Physiatrists Before, During, and Beyond the COVID-19 Pandemic": RehabilitationAndTelemedicineT.pdf

### Rehabilitation and Telemedicine/Telehealth During COVID-19 Pandemic

Hello, we (The Rehabilitation Robotics Lab at the University of Pennsylvania) are trying to understand the implications of COVID-19 on rehabilitation care, opportunities for telemedicine/telehealth, and feelings towards robotics. We believe that at this time, it is critical to understand and share the needs of rehab providers and their patients.

You are being invited to complete a survey on these topics as a clinician who works in rehabilitation. The entire survey should take you no more than 15 minutes.

We are attempting to collect 500 responses from physiatrists, occupational therapists, physical therapists, speech and language pathologists, medical and rehab technologists, and other rehab professionals, so please share the survey with your co-workers and network.

We will be giving a $20 Amazon gift card randomly to one in every 20 respondents on a rolling basis. To be eligible you need to complete the survey and opt-in to the drawing with your e-mail at the end. You can also opt in to be notified of publication of the data.

You are under no obligation to start or complete this survey. If you have any questions or concerns about the administration of this survey, please contact the Institutional Review Board (IRB) at (215) 898-2614.

Note: Options with a "+" in a circle in the choices allow multiple selections. Please select all relevant options.

**Progress:**

,

**This section of the survey will ask about you and your practice as it runs normally (prior to the COVID-19 pandemic).**

| | |
|---|---|
| Are you: | ◯ A Therapist<br>◯ A Nurse<br>◯ A Doctor<br>◯ A Medical Technologist<br>◯ Other type of clinician providing rehabilitation services |
| What type of therapist are you? | __________________________________ |
| What type of nurse are you? | __________________________________ |
| What type of doctor are you? | __________________________________ |
| What type of medical technologist are you? | __________________________________ |
| What type of clinician are you? | __________________________________ |
| What is your age (years) | __________________________________ |

| | |
|---|---|
| What is your gender? | ◯ Male<br>◯ Female<br>◯ Other |
| What type of center(s) do you work in?<br>Select all that apply | ☐ School<br>☐ Hospital for Children<br>☐ General Hospital<br>☐ Elder Care Hospital<br>☐ Rehab Center<br>☐ Elder Care Home<br>☐ Community Center<br>☐ Private Practice<br>☐ Patient Home<br>☐ Inpatient Facility<br>☐ Outpatient Facility<br>☐ Other |
| Please specify your work location: | ______________________________ |
| Do you work in a suburban, urban, and/or rural setting?<br>Select all that apply | ☐ Urban<br>☐ Suburban<br>☐ Rural |

What country do you primarily work in?

- ◯ United States of America
- ◯ Canada
- ◯ Mexico
- ◯ Afghanistan
- ◯ Albania
- ◯ Algeria
- ◯ Andorra
- ◯ Angola
- ◯ Antigua and Barbuda
- ◯ Argentina
- ◯ Armenia
- ◯ Australia
- ◯ Austria
- ◯ Azerbaijan
- ◯ Bahamas
- ◯ Bahrain
- ◯ Bangladesh
- ◯ Barbados
- ◯ Belarus
- ◯ Belgium
- ◯ Belize
- ◯ Benin
- ◯ Bhutan
- ◯ Bolivia (Plurinational State of)
- ◯ Bosnia and Herzegovina
- ◯ Botswana
- ◯ Brazil
- ◯ Brunei Darussalam
- ◯ Bulgaria
- ◯ Burkina Faso
- ◯ Burundi
- ◯ Cabo Verde
- ◯ Cambodia
- ◯ Cameroon
- ◯ Central African Republic
- ◯ Chad
- ◯ Chile
- ◯ China
- ◯ Colombia
- ◯ Comoros
- ◯ Congo
- ◯ Costa Rica
- ◯ Côte D'Ivoire
- ◯ Croatia
- ◯ Cuba
- ◯ Cyprus
- ◯ Czech Republic
- ◯ Democratic People's Republic of Korea
- ◯ Democratic Republic of the Congo
- ◯ Denmark
- ◯ Djibouti
- ◯ Dominica
- ◯ Dominican Republic
- ◯ Ecuador
- ◯ Egypt
- ◯ El Salvador
- ◯ Equatorial Guinea
- ◯ Eritrea
- ◯ Estonia
- ◯ Eswatini (the Kingdom of)
- ◯ Ethiopia
- ◯ Fiji
- ◯ Finland
- ◯ France
- ◯ Gabon
- ◯ Gambia (Republic of The)
- ◯ Georgia
- ◯ Germany
- ◯ Ghana

◯ Greece
◯ Grenada
◯ Guatemala
◯ Guinea
◯ Guinea Bissau
◯ Guyana
◯ Haiti
◯ Honduras
◯ Hungary
◯ Iceland
◯ India
◯ Indonesia
◯ Iran (Islamic Republic of)
◯ Iraq
◯ Ireland
◯ Israel
◯ Italy
◯ Jamaica
◯ Japan
◯ Jordan
◯ Kazakhstan
◯ Kenya
◯ Kiribati
◯ Kuwait
◯ Kyrgyzstan
◯ Lao People's Democratic Republic
◯ Latvia
◯ Lebanon
◯ Lesotho
◯ Liberia
◯ Libya
◯ Liechtenstein
◯ Lithuania
◯ Luxembourg
◯ Madagascar
◯ Malawi
◯ Malaysia
◯ Maldives
◯ Mali
◯ Malta
◯ Marshall Islands
◯ Mauritania
◯ Mauritius
◯ Micronesia (Federated States of)
◯ Monaco
◯ Mongolia
◯ Montenegro
◯ Morocco
◯ Mozambique
◯ Myanmar
◯ Namibia
◯ Nauru
◯ Nepal
◯ Netherlands
◯ New Zealand
◯ Nicaragua
◯ Niger
◯ Nigeria
◯ Norway
◯ Oman
◯ Pakistan
◯ Palau
◯ Panama
◯ Papua New Guinea
◯ Paraguay
◯ Peru
◯ Philippines
◯ Poland
◯ Portugal
◯ Qatar
◯ Republic of Korea

◯ Republic of Moldova
◯ Romania
◯ Russian Federation
◯ Rwanda
◯ Saint Kitts and Nevis
◯ Saint Lucia
◯ Saint Vincent and the Grenadines
◯ Samoa
◯ San Marino
◯ Sao Tome and Principe
◯ Saudi Arabia
◯ Senegal
◯ Serbia
◯ Seychelles
◯ Sierra Leone
◯ Singapore
◯ Slovakia
◯ Slovenia
◯ Solomon Islands
◯ Somalia
◯ South Africa
◯ South Sudan
◯ Spain
◯ Sri Lanka
◯ Sudan
◯ Suriname
◯ Sweden
◯ Switzerland
◯ Syrian Arab Republic
◯ Tajikistan
◯ Thailand
◯ The former Yugoslav Republic of Macedonia
◯ Timor-Leste
◯ Togo
◯ Tonga
◯ Trinidad and Tobago
◯ Tunisia
◯ Turkey
◯ Turkmenistan
◯ Tuvalu
◯ Uganda
◯ Ukraine
◯ United Arab Emirates
◯ United Kingdom of Great Britain and Northern Ireland
◯ United Republic of Tanzania
◯ Uruguay
◯ Uzbekistan
◯ Vanuatu
◯ Bolivarian Republic of
◯ Viet Nam
◯ Yemen
◯ Zambia
◯ Zimbabwe

What state do you primarily work in?

- ◯ Alabama
- ◯ Alaska
- ◯ Arizona
- ◯ Arkansas
- ◯ California
- ◯ Colorado
- ◯ Connecticut
- ◯ Delaware
- ◯ Florida
- ◯ Georgia
- ◯ Hawaii
- ◯ Idaho
- ◯ Illinois
- ◯ Indiana
- ◯ Iowa
- ◯ Kansas
- ◯ Kentucky
- ◯ Louisiana
- ◯ Maine
- ◯ Maryland
- ◯ Massachusetts
- ◯ Michigan
- ◯ Minnesota
- ◯ Mississippi
- ◯ Missouri
- ◯ Montana
- ◯ Nebraska
- ◯ Nevada
- ◯ New Hampshire
- ◯ New Jersey
- ◯ New Mexico
- ◯ New York
- ◯ North Carolina
- ◯ North Dakota
- ◯ Ohio
- ◯ Oklahoma
- ◯ Oregon
- ◯ Pennsylvania
- ◯ Rhode Island
- ◯ South Carolina
- ◯ South Dakota
- ◯ Tennessee
- ◯ Texas
- ◯ Utah
- ◯ Vermont
- ◯ Virginia
- ◯ Washington
- ◯ West Virginia
- ◯ Wisconsin
- ◯ Wyoming
- ◯ Washington, D.C.

How many years have you been practicing?

__________________________________

| | |
|---|---|
| Please select descriptors for the type of patients you treat. Patients with..<br>Select all that apply | ☐ CP<br>☐ Stroke<br>☐ Brachial Plexus Injuries<br>☐ Autism<br>☐ TBI<br>☐ Cardiovascular Disease<br>☐ Speech Disorders<br>☐ Sports Injuries<br>☐ Amputations<br>☐ Arthritis<br>☐ Dementia<br>☐ Burns<br>☐ General Traumatic Injuries<br>☐ Mental Health Issues<br>☐ Other |
| Please describe the other patient types who you treat: | ______________________________ |
| What are the levels of COGNITIVE impairment that your patients have?<br>Select all that apply | ☐ Severely Impaired<br>☐ Moderately Impaired<br>☐ Mildly Impaired<br>☐ Not Impaired |
| What are the levels of MOTOR impairment that your patients have?<br>Select all that apply | ☐ Severely Impaired<br>☐ Moderately Impaired<br>☐ Mildly Impaired<br>☐ Not Impaired |
| What age groups do you generally work with?<br>Select all that apply | ☐ Infants (0-1)<br>☐ Toddlers (1-3)<br>☐ Preschoolers (3-5)<br>☐ Gradeschoolers (5-12)<br>☐ Teens (12-18)<br>☐ Young Adults (18-21)<br>☐ Adults (21-40)<br>☐ Middle Aged Adults (40-65)<br>☐ Older Adults (65+) |
| What phase of care are your patients in?<br>Select all that apply | ☐ Acute<br>☐ Sub-Acute<br>☐ Chronic |
| Do you use any of these tools to interact with patients during normal practice (prior to the COVID-19 pandemic)?<br>Select all that apply | ☐ Phone calls with patients<br>☐ Text based communication (email, text message, text app, etc.)<br>☐ Video calls to patients at home<br>☐ Video calls to patients in another clinical setting<br>☐ Other method of remotely connecting with patients<br>☐ None of these |
| What is the other method you have used? | ______________________________ |

**Progress:**
**20%**

**In this section we will ask you about your experience delivering care to your patients as a result of the COVID-19 pandemic.**

Have you been able to see your patients in-person during the pandemic?
○ Yes ○ No

How has the pandemic changed your level of interaction with your patients?
I cannot interact with my patients — I am interacting with my patients as normal — I am interacting with my patients much more than normal
*(Place a mark on the scale above)*

How do you expect the pandemic to affect your patients' long term health outcomes?
Much worse outcomes — Unchanged outcomes — Much better outcomes
*(Place a mark on the scale above)*

How do you believe the pandemic is affecting your patients' ability to care for themselves and meet their rehabilitation goals?
Much worse — Unchanged — Much better
*(Place a mark on the scale above)*

Are you using any of these tools to connect with your patients during the COVID-19 pandemic?
Select all that apply
- ☐ Phone calls with patients
- ☐ Text based communication (email, text message, text app, etc.)
- ☐ Video calls to patients at home
- ☐ Video calls to patients in another clinical setting
- ☐ Other method of remotely connecting with patients
- ☐ None of these

What is the other method you are using?
__________________________________

Do you plan to use video calls to talk to your patients going forward during the COVID-19 pandemic?
○ Yes
○ No

**Progress:**

**,**

**In this section we will ask you about your experience and thoughts about using telemedicine, specifically video calls, both before and during the COVID-19 pandemic.**

Please rate your overall SATISFACTION using video calls for rehab

Very Dissatisfied | Neutral | Very Satisfied

*(Place a mark on the scale above)*

How well are you able to COMMUNICATE with your patients over video calls compared to in-person?

In-person much better communication | No difference | Video call much better communication

*(Place a mark on the scale above)*

How well do you believe you could COMMUNICATE with your patients over video calls compared to in-person?

In-person much better communication | No difference | Video call much better communication

*(Place a mark on the scale above)*

How well are you able to ASSESS your patients' level of function over video calls compared to in-person?

In-person much better assessment | No difference | Video call much better assessment

*(Place a mark on the scale above)*

How well do you believe you could ASSESS your patients' level of function over video calls compared to in-person?

In-person much better assessment | No difference | Video call much better assessment

*(Place a mark on the scale above)*

How MOTIVATED are your patients DURING a video call compared to in-person?

In-person much more motivated | No difference | Video call much more motivated

*(Place a mark on the scale above)*

How MOTIVATED do you believe your patients would be DURING a video call compared to in-person?

In-person much more motivated | No difference | Video call much more motivated

*(Place a mark on the scale above)*

How well do your patients COMPLY with instructions DURING a video call vs in-person visit?

In-person much higher compliance | The same | Video call much higher compliance

*(Place a mark on the scale above)*

How well do you believe your patients would COMPLY with instructions DURING a video call vs in-person visit?

In-person much higher compliance | The same | Video call much higher compliance

*(Place a mark on the scale above)*

How well do your patients ADHERE to the treatment plan AFTER a video call vs in-person visit?

In-person much higher adherence | The same | Video call much higher adherence

*(Place a mark on the scale above)*

How well do you believe your patients would ADHERE to the treatment plan AFTER a video call vs in-person visit?

In-person much higher compliance — The same — Video call much higher compliance

(Place a mark on the scale above)

---

Do you face any challenges using video calls that affect quality of care?
Select all that apply

- ☐ No challenges
- ☐ connection failures
- ☐ poor audio quality
- ☐ poor video quality
- ☐ Patients do not have a device for video
- ☐ Patients do not have internet
- ☐ Patients cannot understand what I want
- ☐ I cannot understand what patients want
- ☐ Patients cannot complete activities without my physical intervention
- ☐ Assessments require the use of a tool which the patient does not have
- ☐ It is not safe for patients to do the desired activities without a professional present
- ☐ Other challenges

---

Do you believe you would face any challenges using video calls that would affect quality of care?
Select all that apply

- ☐ No challenges
- ☐ Connection failures
- ☐ Poor audio quality
- ☐ Poor video quality
- ☐ Patients would not have a device for video
- ☐ Patients would not have internet
- ☐ Patients would not understand what I want
- ☐ I would not understand what patients want
- ☐ Patients could not complete activities without my physical intervention
- ☐ Assessments require the use of a tool which the patient would not have
- ☐ It would not be safe for patients to do the desired activities without a professional present
- ☐ Other challenges

---

What other challenges?

__________________________________________

---

Do you plan to use video calls with your patients for your practice after the COVID-19 pandemic has ended?

- ◯ Yes
- ◯ No
- ◯ Not Sure

**Progress:**

,

**In this section we present a new robotic platform for telemedicine, specifically telerehabilitation. We then ask some questions about the system and how it could fit into your practice.**

Please watch this video which describes the Lil'Flo Robot, a socially assitive telerehabilitation robot.

The system consists of a social robot (a robot which interacts with people socially), which can play games, demonstrate motions, and express emotion, mounted on a mobile telepresence system which has a screen, cameras, and a microphone.

Then continue to answer questions about the system.

---

Do you have any prior knowledge of the Lil'Flo system?
Select all that apply

- ☐ No prior knowledge
- ☐ I have read a paper on the system
- ☐ I have seen the system in person
- ☐ I have used the system
- ☐ I have some other experience with system

---

How interested would you be in using the Lil'Flo system?

Not At All Interested — Very Interested

*(Place a mark on the scale above)*

---

What locations do you think Lil'Flo could be deployed in?
Select all that apply

- ☐ Rural outpatient clinics
- ☐ Rural inpatient clinics
- ☐ Elder care facilities
- ☐ Schools
- ☐ Patient homes
- ☐ Community centers
- ☐ Urban inpatient clinics
- ☐ Urban outpatient clinics
- ☐ None
- ☐ Other

---

What other locations?

__________________________________

---

How do you believe that adding a social robot as a companion for your patients during video+audio telepresence interactions (such as the LIl'Flo system) would change the following when compared with traditional video+audio telepresence based rehab?

---

COMMUNICATION during the interaction

Decrease communication — No change — Help communication

*(Place a mark on the scale above)*

---

Patient MOTIVATION during the interaction

Decrease motivation — No change — Increase motivation

*(Place a mark on the scale above)*

Your ability to ASSESS your patients from telepresence interactions

Impair assessment — Same — Improve assessment

(Place a mark on the scale above)

How well your patients COMPLY with instructions DURING the telepresence interaction

Reduce compliance — Same — Improve compliance

(Place a mark on the scale above)

How well your patients ADHERE to the treatment plan AFTER a telepresence interaction

Reduce adherence — Same — Improve adherence

(Place a mark on the scale above)

What features do you believe are useful in a system to make tele-rehabilitation work well?

| | Extremely Useless | Somewhat Useless | Neutral | Somewhat Useful | Extremely Useful |
|---|---|---|---|---|---|
| A mobile system which can be driven remotely | ○ | ○ | ○ | ○ | ○ |
| A mobile system which can drive on its own | ○ | ○ | ○ | ○ | ○ |
| A social robot with arms to augment standard video calls | ○ | ○ | ○ | ○ | ○ |
| A social robot with an expressive face to augment standard video calls | ○ | ○ | ○ | ○ | ○ |
| A social robot that can interact with patients without needing operator input | ○ | ○ | ○ | ○ | ○ |
| A web interface for controlling remote telerehabilitation systems | ○ | ○ | ○ | ○ | ○ |
| A social robot which can play games with subjects during telerehabilitation calls to augment standard video | ○ | ○ | ○ | ○ | ○ |
| A system which collects data and performs automated assessments of patient function | ○ | ○ | ○ | ○ | ○ |
| A clear screen to see the clinician | ○ | ○ | ○ | ○ | ○ |
| High quality video to see the patient | ○ | ○ | ○ | ○ | ○ |

Please list any other features which you believe would be useful.

__________________________________

**Progress:**

,

**We would like to know which tools you think are appropriate for different tasks.**

Which types of activities HAVE YOU DONE with each type of tool?

Non-video remote communication includes: phone calls, text messages, email, instant messages, and other types of communcation which allow you to interact with patients from afar, without using video.

Select all that apply

| | Non-Video Remote Interaction | Video Call Based Remote Interaction | Lil'Flo Based Remote Interaction | In Person | Have Not Done |
|---|---|---|---|---|---|
| Motor Assessments | ☐ | ☐ | ☐ | ☐ | ☐ |
| Stretching | ☐ | ☐ | ☐ | ☐ | ☐ |
| Strength Building | ☐ | ☐ | ☐ | ☐ | ☐ |
| ADL Practice | ☐ | ☐ | ☐ | ☐ | ☐ |
| Cognitive Assessments | ☐ | ☐ | ☐ | ☐ | ☐ |
| Cognitive Exercises | ☐ | ☐ | ☐ | ☐ | ☐ |
| Environmental Adaptation | ☐ | ☐ | ☐ | ☐ | ☐ |
| Orthotics Assessment/Prescription | ☐ | ☐ | ☐ | ☐ | ☐ |
| Discussions about Surgery | ☐ | ☐ | ☐ | ☐ | ☐ |
| Discussions about Radiology Results | ☐ | ☐ | ☐ | ☐ | ☐ |
| Medical Prescriptions | ☐ | ☐ | ☐ | ☐ | ☐ |

Which types of activities DO YOU BELIEVE YOU COULD DO with each type of tool?

Non-video remote communication includes: phone calls, text messages, email, instant messages, and other types of communcation which allow you to interact with patients from afar, without using video.

Select all that apply

| | Non-Video Remote Interaction | Video Call Based Remote Interaction | Lil'Flo Based Remote Interaction | Could Not Be Done Remotely |
|---|---|---|---|---|
| Motor Assessments | ☐ | ☐ | ☐ | ☐ |
| Stretching | ☐ | ☐ | ☐ | ☐ |
| Strength Building | ☐ | ☐ | ☐ | ☐ |
| ADL Practice | ☐ | ☐ | ☐ | ☐ |
| Cognitive Assessments | ☐ | ☐ | ☐ | ☐ |
| Cognitive Exercises | ☐ | ☐ | ☐ | ☐ |
| Environmental Adaptation | ☐ | ☐ | ☐ | ☐ |
| Orthotics Assessment/Prescription | ☐ | ☐ | ☐ | ☐ |
| Discussions about Surgery | ☐ | ☐ | ☐ | ☐ |

| | | | | |
|---|---|---|---|---|
| Discussions about Radiology Results | ☐ | ☐ | ☐ | ☐ |
| Medical Prescriptions | ☐ | ☐ | ☐ | ☐ |

**Progress:**

**,**

**In this section we ask for some final thoughts on robotics in healthcare.**

| | Strongly Disagree | Disagree | Neutral | Agree | Strongly Agree |
|---|---|---|---|---|---|
| I have general positive feelings towards robots | ○ | ○ | ○ | ○ | ○ |
| Robots are the same as any other medical/assistive devices | ○ | ○ | ○ | ○ | ○ |
| Robots will improve patient independence | ○ | ○ | ○ | ○ | ○ |
| Robots allow other caretakers to focus on higher level work | ○ | ○ | ○ | ○ | ○ |
| Robots are consistent and accurate | ○ | ○ | ○ | ○ | ○ |
| Robots are too expensive | ○ | ○ | ○ | ○ | ○ |
| Robots take too much caretaker time to set up | ○ | ○ | ○ | ○ | ○ |
| The use of robots for care and treatment is unethical | ○ | ○ | ○ | ○ | ○ |
| Patients don't want robots to take care of them | ○ | ○ | ○ | ○ | ○ |
| I want robots that I can control | ○ | ○ | ○ | ○ | ○ |
| Robots are going to take jobs | ○ | ○ | ○ | ○ | ○ |
| Robots are too likely to break or fail | ○ | ○ | ○ | ○ | ○ |
| Robots will improve clinician well-being | ○ | ○ | ○ | ○ | ○ |
| I have general negative feelings towards robots | ○ | ○ | ○ | ○ | ○ |
| Failing to use robots for care and treatment is unethical | ○ | ○ | ○ | ○ | ○ |
| Using robots will damage people's ability to interact with other people | ○ | ○ | ○ | ○ | ○ |
| We need more laws and regulations about robots | ○ | ○ | ○ | ○ | ○ |
| We need to study more about how robots affect function | ○ | ○ | ○ | ○ | ○ |
| I want robots that act on their own | ○ | ○ | ○ | ○ | ○ |

| | | | | | |
|---|---|---|---|---|---|
| Robots are bad for society | ○ | ○ | ○ | ○ | ○ |

What other feelings do you have towards robots?

__________________________________

If you have any other general thoughts you would like to share with the community about COVID-19's impacts on your practice and patients, telemedicine/telepresence, Lil'Flo, etc. Please share them here. This is the final question of the survey.

__________________________________

**Progress:**

**,**

| | |
|---|---|
| Would you like to be entered into the drawing for a $20 Amazon gift card? | ◯ Yes ◯ No |
| Would you like to be made aware of the results of this survey when they are published? | ◯ Yes ◯ No |
| Please provide your email | __________________________ |
